# Parallel Analysis of Pharmaceuticals and Viruses in Wastewater: An Untargeted Approach to Tracking Population-Level Respiratory Illness Burden

**DOI:** 10.1101/2024.10.16.24315620

**Authors:** Stephan Baumgartner, Michelle Salvisberg, Patrick Schmidhalter, Timothy R Julian, Christoph Ort, Heinz Singer

## Abstract

Wastewater-based surveillance is widely used for the detection and tracking of priority respiratory pathogens. In this study, pharmaceuticals are tracked in wastewater as an untargeted indicator of symptoms related to acute respiratory infections and influenza-like illnesses, such as coughing, fever, and pain, coincident to respiratory viral loads.

From January 2021 to June 2024, wastewater samples from ten wastewater treatment plants across Switzerland serving 23% of the population were analyzed. The analysis encompassed 15 pharmaceuticals and priority respiratory viruses SARS-CoV-2, respiratory syncytial virus, influenza A, and influenza B. Pharmaceutical data were further benchmarked against national clinical surveillance data on a broader range of respiratory pathogens.

The pharmaceutical compounds dextrorphan (indicative of dextromethorphan), pheniramine, clarithromycin, acetaminophen, and codeine showed a strong correlation with respiratory viral loads. This enabled the estimation of pathogen-specific and cumulative symptom burdens in the population. Additionally, notable increases in pharmaceutical loads without corresponding increases in viral loads in 2021 and 2024 signaled high community symptom burdens linked to unsurveilled pathogens.

The study demonstrates that pharmaceutical surveillance can inform respiratory disease burden and highlights the broader potential of integrated surveillance to assess population-scale symptoms, indicating emerging public health threats beyond those routinely monitored.

## 2 Main

Wastewater contains a diverse array of human excreta, offering a valuable source of epidemiological information at the population level. Historically, wastewater-based surveillance (WBS) has been used for analysis of small molecules, such as illicit drugs, pharmaceuticals, and lifestyle chemicals, to estimate population drug consumption [1]. The detection of severe acute respiratory syndrome coronavirus-2 (SARS-CoV-2) RNA in wastewater – which showed strong correlations with clinical cases – marked a rapid uptake in WBS for pathogen surveillance globally [2]. The rigorous, simultaneous monitoring of SARS-CoV-2 in both clinical settings and wastewater provided a unique opportunity to validate WBS as an objective, inclusive, and cost-effective tool for public health monitoring. As WBS has gained recognition and institutionalization, its scope has expanded to cover a wide range of pathogens beyond SARS-CoV-2, including additional respiratory viruses, gastrointestinal pathogens, hepatitis, and monkeypox virus [3–9].

In addition to tracking pathogens, WBS was applied to chemicals during the pandemic to examine shifts in the consumption of illicit drugs and pharmaceuticals, illustrating the widespread impacts of the pandemic on lifestyle and emotional health [10–13].

Although chemicals and pathogens were investigated extensively, combined analyses are rare and focused exclusively on COVID-19 [14,15]. However, chemical surveillance, including pharmaceutical analysis, offers a promising avenue for the assessment of the level of symptom treatment in a population, as a proxy for the disease burden in an untargeted approach. Pharmaceutical loads in wastewater thereby provide rapid feedback on the population’s response during infectious disease outbreaks, complementing the highly targeted nucleic acid quantification assays of priority pathogens. This enables the estimation of symptom burdens for monitored pathogens and the detection of emerging, unsurveilled threats, such as “Pathogen X” [16–18].

In this study, chemical surveillance focused on pharmaceuticals used to alleviate symptoms of acute respiratory infections and influenza-like illnesses, specifically coughing, fever, and pain. We analyzed wastewater samples from ten wastewater treatment plants (WWTPs) serving approximately 23% of the Swiss population, collected from 2021 to mid-2024. Pharmaceutical surveillance was complemented by wastewater measurements for priority respiratory viruses: SARS-CoV-2, human respiratory syncytial virus (RSV), influenza A virus (IAV), and influenza B virus (IBV). These viruses are recognized as leading causes of respiratory illnesses and impose significant health and economic burdens on communities [19–21]. Furthermore, the data were compared to national-level sentinel surveillance to explore additional respiratory viruses as explanatory variables for pharmaceutical consumption.

With this analysis we aimed to:

i. Quantify the dynamics of pharmaceutical consumption in wastewater as an indicator of symptomatic surveillance.
ii. Examine the relationship between symptomatic treatment and phases of respiratory viral exposure to better understand population-level symptom burdens.
iii. Identify periods of high levels of treatment without corresponding etiologies to highlight potential contributions from unsurveilled pathogens.

### Spatiotemporal Trends of Symptom Treatment Pharmaceuticals and Respiratory Viruses in Wastewater

Wastewater samples were collected from ten WWTPs throughout Switzerland, representing diverse geographic and socioeconomic contexts, including Zurich, Basel, Bern, Solothurn, Chur, Schwyz (German-speaking), Geneva, Lausanne, Neuchâtel (French-speaking), and Lugano (Italian-speaking). These WWTPs cover the five largest cities as well as rural areas, with catchment populations ranging from 31,000 to 471,000 individuals, collectively serving around 2.02 million people (Figure SI1). Chemical analysis was performed on samples collected every 13th day to ensure each weekday was represented once per quarter between January 2021 and June 2024.

We monitored fifteen chemicals in wastewater, including the opioid analog dextrorphan (the main urine metabolite of the cough suppressant dextromethorphan), the opioids codeine, morphine, and tramadol, and non-opioid analgesics acetaminophen (paracetamol), diclofenac, and naproxen. Additionally, pheniramine and its metabolite N-desmethylpheniramine were assessed due to its use in Switzerland, where it is found exclusively in over-the-counter combination products for treating flu and cold symptoms (Table SI12). Furthermore, the antibiotics clarithromycin (macrolide), sulfamethoxazole (sulfonamide), trimethoprim (antifolate), and metronidazole (nitroimidazole) were quantified.

Candesartan and paraxanthine were included as control compounds, with their loads presumed to be stable over time. This allowed for a direct control over random or systematic deviations in the observed temporal patterns, which could arise from factors such as unknown changes in population dynamics or unreported variations in sampling techniques. We also conducted in-sample stability experiments, demonstrating that transformation of compounds is negligible, under the study’s specified conditions of 4°C for short-term handling and -20°C for long-term storage (SI 3).

Figure 1 displays the population-normalized loads of individual samples for the pharmaceuticals dextrorphan, pheniramine, clarithromycin, acetaminophen, and codeine across the ten locations.

**Figure 1:**
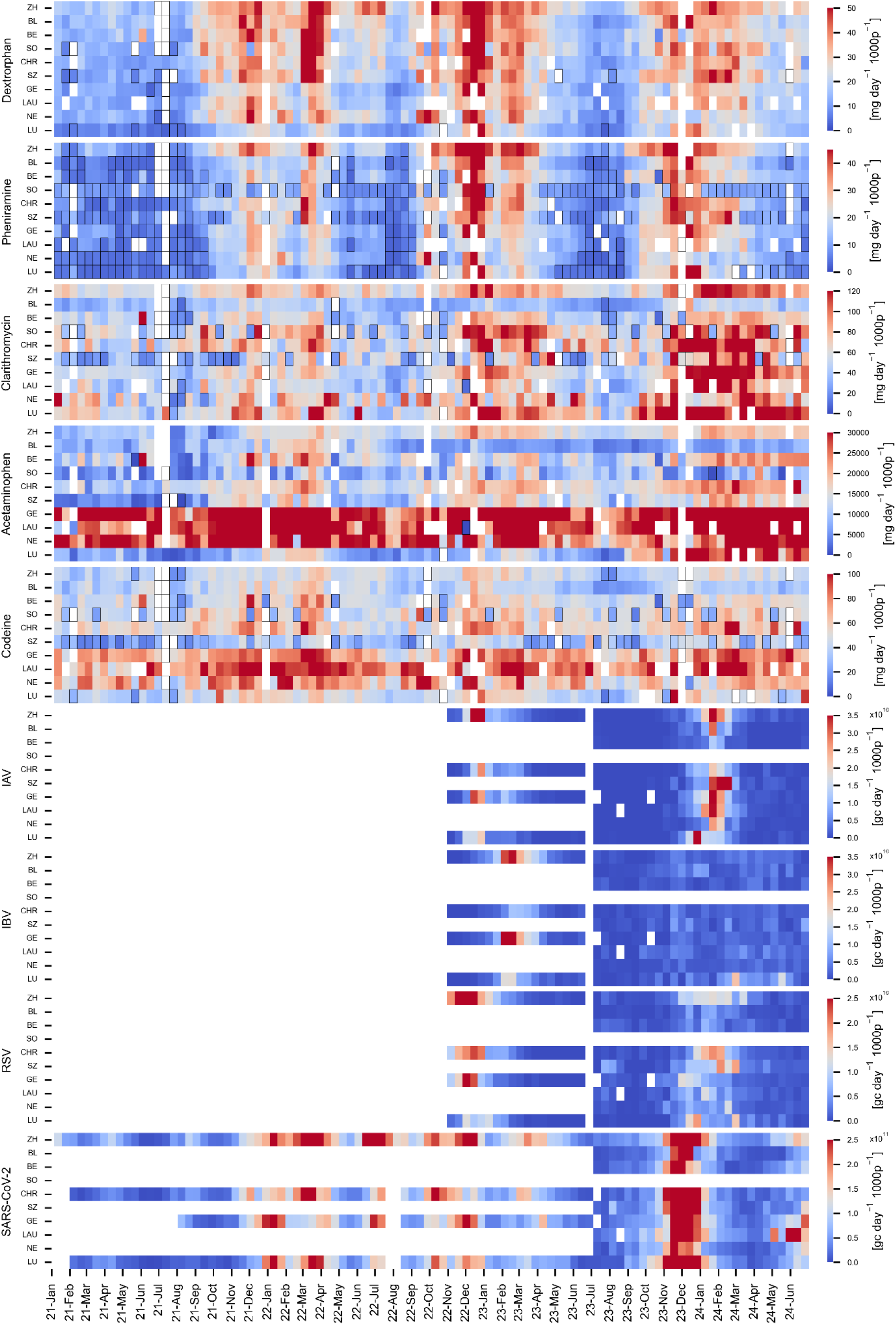
Spatiotemporal Load Patterns of Pharmaceuticals and Viral Gene Copies in Wastewater. Wastewater loads from every 13th day between January 2021 and June 2024 are displayed. Data is shown for individual treatment plants according to the abbreviations on the Y-axis: ZH (Zurich), BL (Basel), BE (Bern), SO (Solothurn), CHR (Chur), SZ (Schwyz), GE (Geneva), LA (Lausanne), NE (Neuchâtel), and LU (Lugano). These are arranged first by language region, then by catchment population size. The coloring represents the load values for pharmaceuticals [mg d− ^1^ 1000p− ^1^d− ^1^] and viruses [gc d−^1^ 1000p−1] with linear color scaling. For pharmaceuticals, each space refers to an individual sample of a particular day, and for the viral loads, each space reflects values from the centered 7-day median. White spaces indicate data exclusions due to rain-related increases in wastewater volumes exceeding maximum threshold values or due to absence of sampling. Black-outlined spaces represent pharmaceutical loads from samples with concentrations below the limit of quantification (LOQ), which are estimated based on half the LOQ value.

Pronounced seasonal trends were observed in the wastewater for several pharmaceuticals monitored. Higher loads of dextrorphan and pheniramine were measured typically during the winter months with occasional increases in summer months, such as June and July 2022. Clarithromycin followed a similar seasonal pattern from 2021 to 2023; however, its levels remained consistently elevated in the first half of 2024.

Other pharmaceuticals did not show clear seasonality with trends obscured by pronounced spatial variation in loads. Codeine had notably higher levels observed in the French-speaking regions of Switzerland at mean 75 mg/d/1000p as compared to non-French-speaking regions of 45 mg/d/1000p. Similarly, acetaminophen was higher in French-speaking regions with mean 33‘531 mg/d/1000p as compared to 12‘441 mg/d/1000p in non-French-speaking regions. This disparity suggests that regional usage practices, potentially shaped by Switzerland’s decentralized healthcare system and fragmented decision-making [22], exert a substantial influence on pharmaceutical consumption patterns.

From 2021 through late 2022, viral analysis was conducted daily for SARS-CoV-2 and limited to four WWTPs. The scope expanded in late October 2022 to include RSV, IAV, and IBV. Since July 2023, these four pathogens were measured in all ten WWTPs, with the frequency reduced to 5 days per week (Figure1).

The missing data for pharmaceuticals, indicated by white spaces, are due the exclusion of values on rainy days when the inflow volume at WWTPs exceeded an empirically determined location-specific maximum threshold (Table SI1, Figure SI2 and Figure SI3). Beyond this threshold, estimated loads are consistently underestimated or overestimated across the substance spectrum. This discrepancy is linked to the potential discharge of untreated wastewater through combined sewer overflows and to inaccuracies in sampling or flow measurements at the WWTPs during high flow conditions. These events usually span over several locations. In contrast, viral gene copy load data from these dates were still considered reliable, as they represent 7-day centered median values, which reduce the impact of outliers.

The compounds dextrorphan, pheniramine, clarithromycin, acetaminophen, and codeine were selected for detailed analysis based on their significant positive Pearson correlation with viral loads of SARS-CoV-2, RSV, and IAV, performed both location-dependent and independent (Figure SI13 – Figure SI19). While the manuscript focuses on these pharmaceuticals, analogous analyses for the additional chemicals are provided in the SI.

### Interpreting National-Level Symptom-Relief Pharmaceutical Usage via Wastewater and Clinical Virus Data

For dextrorphan, pheniramine, and to a lesser extent clarithromycin, temporal fluctuations were relatively consistent across locations, as evidenced by the narrow 10% to 90% interpercentile range (Figures SI5–Figure SI7). This pattern was also observed in viral loads (Figure SI8), indicating similar trends across the sampled locations. To directly compare wastewater patterns with national-level sentinel clinical testing data for respiratory viruses (Figure 2c), we estimated the median load values from the ten locations as indicators of pharmaceutical consumption (Figure 2a) and viral exposure (Figure 2b) at the national level.

**Figure 2:**
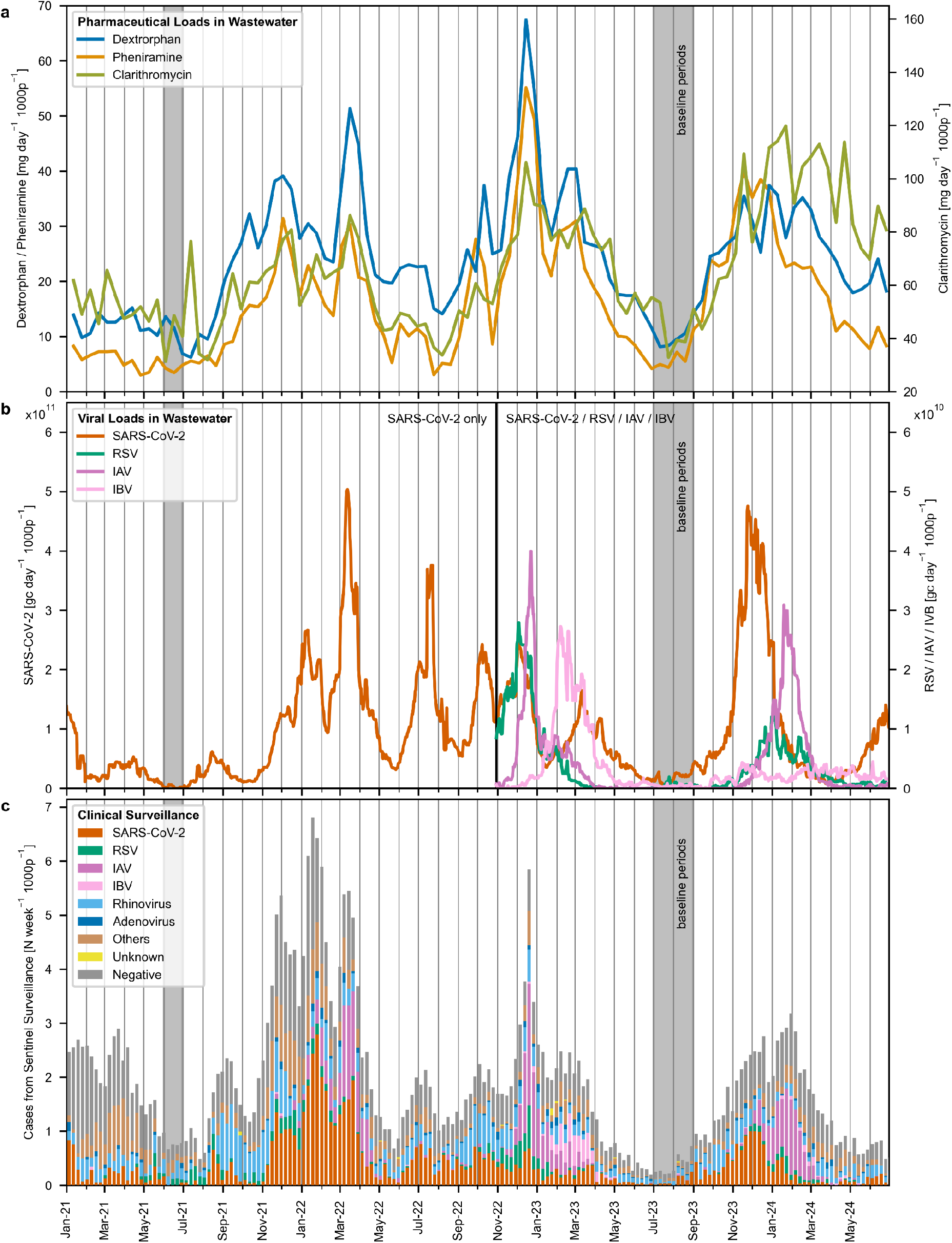
National Trends of Pharmaceutical and Viral Data in Wastewater alongside Clinical Sentinel Testing Data (January 2021 – June 2024). (a) Median pharmaceutical loads shown for every 13th day. (b) Median viral gene copy loads, based on daily 7-day centered median values. (c) Stacked testing cases from general practitioner-based sentinel surveillance system. The “Others” category (shown in grey) encompasses tests for bocavirus, human coronaviruses (229E, HKU1, NL63, OC43), metapneumovirus, and parainfluenza viruses (types 1-4). The “Negative” category includes cases where individuals sought consultation for symptoms of acute respiratory infections or influenza-like illnesses but tested negative for the respiratory viruses under surveillance. The “Unknown” category includes samples with incomplete data on detected respiratory viruses.

The seasonal patterns for dextrorphan, pheniramine and clarithromycine align with seasonality of virus loads. In December 2022, the peak loads of dextrorphan and pheniramine coincided with a SARS-CoV-2 wave and the highest recorded loads of RSV and IAV, highlighting a substantial cumulative viral burden on the population. Subsequently, pharmaceutical loads surged again in February 2023 during an Influenza B wave, presumably of the Victoria subtype (Figure SI10).

In the following winter (2023/2024), the major SARS-CoV-2 wave in November 2023 preceded the RSV and IAV waves, mirroring concurrent peaks in pharmaceutical consumption. Notably, IBV was almost absent in winter of 2023/2024. Between 2021 and October 2022, when only SARS-CoV-2 was monitored, pharmaceutical loads generally rose in tandem with SARS-CoV-2 waves, including during the summer surge in June 2022.

From September to November 2021, an intriguing increase in pharmaceutical loads, particularly dextrorphan, occurred despite relatively low SARS-CoV-2 levels in wastewater. To expand the analysis beyond respiratory pathogens included in wastewater surveillance, we also investigated trends in the publicly available Swiss Sentinel System (Sentinella), a voluntary reporting system with contributions from 160 to 180 family physicians and pediatricians [23]. The sentinel surveillance includes monitoring of additional respiratory viruses such as rhinoviruses and adenoviruses, alongside seasonal human coronaviruses, bocavirus, human metapneumovirus, and parainfluenza viruses (types 1-4), which are grouped under “Others” in Figure 2c. In addition to pathogens, Sentinella tracks the number of individuals who sought medical consultation due to symptoms but tested negative for the monitored viruses, suggesting the presence of alternative etiological factors (the “negative” category”). The summed values from sentinel testing reflects the number of primary consultations for acute respiratory infection or influenza-like illness symptoms (Figure SI11). These consultation counts are multiplied by testing positivity rates (Figure SI12) to calculate weekly virus-specific cases displayed in Figure 2c. During the period of October to November 2021, an increased number of consultations were noted, predominantly testing positive for rhinovirus and “Others.” This period was closely followed by an observed increase in SARS-CoV-2 cases.

Periods marked by low viral exposure for SARS-CoV-2, RSV, IAV, and IBV, as indicated in wastewater and sentinel data from early 2021 and mid-2023, are highlighted with grey bars (Figure 2). To accommodate spatial variation in pharmaceutical loads, particularly for acetaminophen and codeine, location-specific mean loads from these low-activity periods were utilized to establish baselines for subsequent modeling of the influence of respiratory viruses on pharmaceutical consumption.

### Estimating the Contribution of SARS-CoV-2, RSV, Influenza A and B to Acute Respiratory Infection and Influenza-like Symptom Treatment

We employed a linear model to estimate the contributions of SARS-CoV-2, RSV, IAV, and IBV to pharmaceutical consumption, accounting for location-specific baselines at each WWTP. Using Ordinary Least Squares (OLS) regression, we assessed the linear relationship between viral gene copy loads and baseline-subtracted pharmaceutical loads, under the assumption that this relationship remains consistent across locations and time. An empirical baseline was selected instead of estimating a baseline term within the OLS model to prevent the potential obscuring of additional, unsurveilled symptom triggers by artificially reducing the sum of the squared errors. Diagnostics of the OLS modeling are available in SI (Figure SI20 – SI24, Table SI5 – SI9).

The OLS-estimated coefficients, applied to median viral gene copy loads, are compared with the observed median pharmaceutical loads adjusted for baselines in the left panel of Figure 3. This comparison highlights periods where pharmaceutical loads correspond closely to respiratory viral loads and identifies discrepancies. Colored stacked areas in the figure represent the estimated contributions of each virus from late October 2022 onward, when monitoring included all four viruses. The coefficient determined for SARS-CoV-2 in this model was also applied to data from early 2021 through late October 2022, the period during which only this virus was monitored in wastewater.

**Figure 3:**
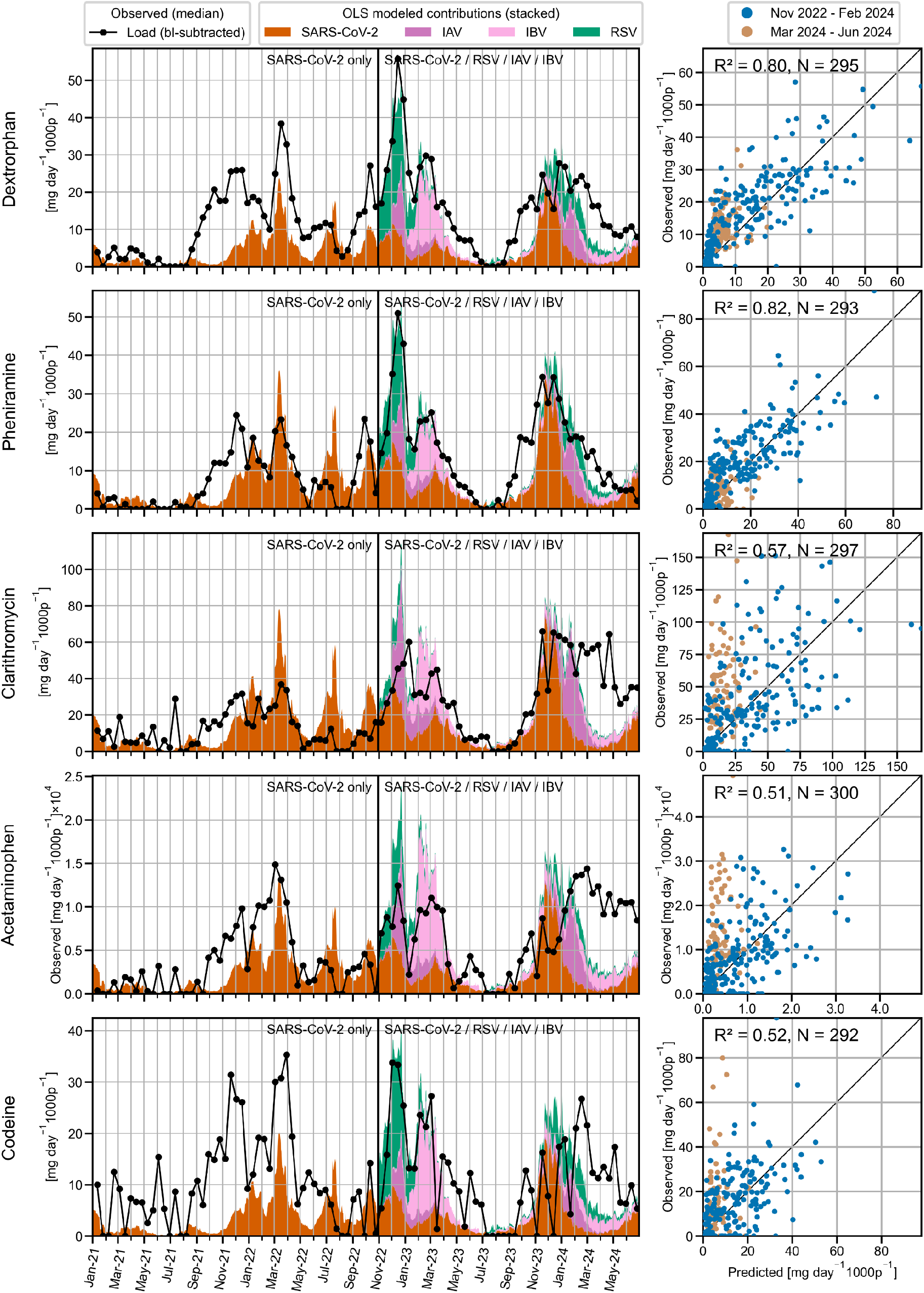
Modeled Contributions of Respiratory Viruses to Pharmaceutical Loads. The left panel presents time series plots of observed baseline-subtracted median pharmaceutical loads (black line) and the modeled contributions, derived from ordinary least squares (OLS) regression coefficients applied to the median viral gene copy loads of SARS-CoV-2, RSV, IAV, and IBV. The colored areas represent the stacked contributions of each virus to the overall baseline-subtracted pharmaceutical load. Baseline loads were determined from location-specific mean values during periods of low respiratory virus exposure (June 2021 and July to August 2023). The OLS coefficient determined for SARS-CoV-2 was applied to data from early 2021 through late October 2022, the period during which only this virus was monitored in wastewater. The right panel displays scatter plots that compare observed versus predicted baseline-subtracted pharmaceutical loads from the OLS analyses. The data points are color-coded by time periods: November 2022 to February 2024 (blue), and March 2024 to June 2024 (orange, indicating poor model fit with viral etiologies). Each scatter plot includes the uncentered coefficient of determination (R^2^) and the number of observations (N). The diagonal black line represents the 1:1 line, indicating perfect agreement between observed and predicted values.

Our analysis indicates that peaks in dextrorphan, pheniramine, and clarithromycin from November 2022 to February 2024, align well with loads of SARS-CoV-2, RSV, IAV, and IBV. However, patterns for acetaminophen and codeine, while elevated during these periods, exhibited spikes that did not consistently align with viral trends. Both codeine and acetaminophen are frequently recommended or used for conditions beyond infectious diseases, including managing chronic pain, osteoarthritis, menstrual pain, and injuries [24,25].

From March to June 2024, a notable discrepancy was observed: high wastewater loads of clarithromycin, acetaminophen, codeine, and, to a lesser extent, dextrorphan persisted even after a decline in respiratory viruses. This inconsistency is further emphasized in the right panel of Figure 3, where the observed versus modeled baseline-subtracted pharmaceutical loads are plotted and color-coded by date periods. Data highlighted in orange indicate deviations from previous patterns, especially for clarithromycin, acetaminophen, and codeine.

Pharmaceutical loads modeled from the beginning of 2021 to October 2022 were consistently underestimated when only SARS-CoV-2 was considered, a trend also evident in OLS models using SARS-CoV-2 as the sole predictive variable (Figure SI27). The analyses reinforce the earlier speculation that SARS-CoV-2 alone does not adequately explain the observed pharmaceutical consumption during the period from September to November 2021.To address such gaps in pharmaceutical consumption explanation, we performed analogous OLS modeling, utilizing case data from the sentinel surveillance system. In an initial analysis we used only SARS-CoV-2, RSV, IAV, and IBV cases as predictors for median baseline-subtracted loads. This provided insights similar to those using wastewater viral loads but improved descriptions for 2021 and 2022 by considering additional viral waves of RSV and IAV (Figure SI28).

Expanding these analyses to case data for rhinovirus and the ‘Others’ category (comprising seasonal human coronaviruses, bocavirus, human metapneumovirus, and parainfluenza viruses), we found that including rhinovirus substantially enhanced the model’s ability to describe pharmaceutical loads, particularly for dextrorphan and pheniramine (Figure SI29 –SI31). This was due to rhinovirus waves preceding other respiratory viruses each year around September, which correlated with early increases in symptom treatment.

From March to June 2024, the OLS analyses showed that none of the respiratory viruses monitored by the sentinel system could account for the increased usage of clarithromycin, acetaminophen, and codeine, suggesting the presence of other contributing factors beyond the surveilled respiratory virus infections.

The observed correlation between clarithromycin and viral loads may suggest its use in treating pharyngitis and pneumonia [26,27], which can arise as complications from viral infections, including viral-bacterial co-infections or as a result of potential misdiagnoses [28–30]. However, clarithromycin is also prescribed for other medical conditions, such as skin infections and Helicobacter pylori infections [31].

Notably, this period in 2024 coincides with a substantial increase in pertussis cases across Europe, including Switzerland, where pertussis had been nearly absent since the start of the COVID-19 pandemic in 2020 [32,33]. This surge may explain the increase in clarithromycin use, as it is one of the first line macrolide antibiotics for treating pertussis [34].

While clinical data from sentinel surveillance yield valuable insights into circulating pathogens, the quantitative estimation of infection rates is susceptible to biases introduced by underreporting and changes in testing regimes. This issue is particularly evident in the Swiss mandatory reporting data for SARS-CoV-2 during the transition to a post-pandemic scenario (Figure SI11). The sentinel data may experience similar biases, and further may not be nationally representative given the relatively low number of participating physicians and potential subjectivity in reporting. These biases can influence the accurate attribution of symptoms to specific viruses. In contrast, wastewater data is considered more stable over time, offering a more consistent basis for analyzing trends in pathogen prevalence and corresponding pharmaceutical usage.

Overall, the OLS analyses demonstrate that the respiratory viruses SARS-CoV-2, RSV, IAV, and IBV can explain most of the observed peaks in pharmaceutical usage throughout the study period, highlighting their impact on symptom burden. Additionally, the observations highlight the reliance on pharmaceuticals from infections with respiratory viruses beyond SARS-CoV-2, including IAV and RSV. Even during the pandemic in 2021, SARS-CoV-2 alone does not explain pharmaceutical use patterns, indicating a need for broader surveillance.

Furthermore, the study identifies periods where pathogens not targeted by our wastewater surveillance, potentially including rhinovirus and pertussis, went undetected but triggered treatment for symptoms at population scale, offering a potential method for detection of novel pathogens into communities (“Pathogen X”).

## Limitations

An important limitation of this study is that the current WBS of pathogens covers only a subset of possible etiologies of disease leading to pharmaceutical use. For much of the period, only SARS-CoV-2 data from wastewater were available; however, analysis of RSV, IAV, and IBV was included from late October 2022 to June 2024. Additionally, there are strong indications that other etiological agents contribute to the population symptom burden, as exemplified by our observations on rhinovirus and pertussis. Extending the timeframe of combined analyses and increasing the number of infectious diseases monitored in wastewater may help to more accurately align pharmaceutical data with disease etiology. The current analysis may cause misattribution of symptoms caused by unsurveilled (and therefore unknown) etiology.

A second limitation relates to the pharmaceutical monitoring methods used. We employed a liquid chromatography-mass spectrometry (LCMS) technique with direct large-volume injection, which facilitated the efficient processing of numerous samples at the expense of sensitivity. More sensitive methods would improve baseline estimation and enable analysis of pharmaceuticals at lower concentrations, but these are more time-consuming and costly, potentially limiting the feasibility of high-frequency measurements.

A final limitation is in our modeling approach. Our OLS modeling assumptions may oversimplify the intricate interactions between pathogen loads and pharmaceutical usage, assuming uniform relationships across different geographical and socio-demographic contexts. Additionally, the model does not accommodate variations in disease severity, which may be tempered by vaccinations or natural immunity, nor does it distinguish between different pathogen variants or subtypes, which might have unique shedding rates and associated symptomatology [35]. Enhancing the analysis to cover extended time periods and to provide finer temporal resolution beyond the current 13-day intervals could delineate the dynamics between viral exposure and pharmaceutical consumption more accurately. Such improvements are important for assessing temporal relation of pathogen activity and drug usage.

## Conclusion

This study combined chemical and microbial surveillance – analyzing pharmaceuticals used to treat symptoms of acute respiratory infections and influenza-like illnesses (coughing, fever, and pain) alongside major respiratory viruses. Parallel analysis demonstrates that pharmaceutical loads can serve as indicators of symptomatic trends at the population level. The integrated analysis established a correlation between pharmaceutical and respiratory viral wastewater loads over time, allowing us to estimate the associated disease burden. Importantly, it highlights the capacity to detect periods of increased health burdens attributable to unsurveilled pathogens, as indicated by clinical data pointing to rhinovirus and pertussis.

Overall, the findings highlight a potential approach for detection of emerging diseases in a community using untargeted approaches. Further, the study advocates for enhanced centralized coordination and interdisciplinary collaboration within chemical and microbial WBS frameworks. Standardizing procedures across sample collection, storage, analysis, and data management can streamline resource use, minimize costs, and ensure consistent data, which are vital for exploiting the full potential of WBS.

## 3 Methods

### Chemicals and Reagents

Details on the chemicals and solutions used for analyzing the fifteen chemical markers - acetaminophen, candesartan, clarithromycin, codeine, dextrorphan, diclofenac, metronidazole, morphine, N-desmethylpheniramine, naproxen, paraxanthine, pheniramine, sulfamethoxazole, tramadol, and trimethoprim - in wastewater are provided in SI 1.2.

### Wastewater Sample Collection and Storage

The sampling campaign, spanning from January 2021 to June 2024, involved the collection of 24-hour composite wastewater samples from the influent of ten wastewater treatment plants (WWTPs) across Switzerland. These samples were collected using on-site sampling infrastructure. Small molecule markers were analyzed in samples collected every 13th day. Together with the daily analyses of samples from Zurich in 2021 (Figure SI5 –SI7), this resulted in the chemical analysis of 1172 wastewater samples. Additionally, viral RNA analysis was conducted initially daily but limited to four treatment plants (Zurich, Chur, Geneva, and Lugano) and focused exclusively on SARS-CoV-2. This scope expanded in late October 2022 to include RSV, IAV, and IBV, and by July 2023, to all ten treatment plants but with a reduced frequency of 5 days per week. In total, microbial analyses were performed on 5480 samples.

Sampling methodologies varied across the WWTPs: volume-proportional sampling was employed for those in Zurich, Bern, Solothurn, Chur, Schwyz, Geneva, and Lugano. In contrast, the Basel WWTP used flow-proportional sampling, and the Lausanne WWTP used time-proportional sampling (Table SI1). The Basel WWTP samples also composited multiple days when sampling occurred on weekends and holidays.

During the 24-hour sampling period, samples were stored at 4°C in auto-samplers. Post-collection, samples destined for chemical analysis were transferred to muffled 100 mL glass bottles and stored at -20°C until analysis. Samples for viral RNA analysis were processed following methodologies outlined in previous studies [36–38].

### Wastewater Sample Preparation for Small Molecule Analysis

The wastewater samples were thawed and centrifuged at room temperature at 280 g for 10 minutes using a Centrifuge 5427 R (Eppendorf). Following centrifugation, 600 µL of the supernatant was transferred to a new glass vial. This supernatant was then spiked with 10 µL of an ethanol-based isotope-labeled internal standard (ISTD) mix containing the 15 structurally identical ISTDs, reaching a final concentration of 1000 ng/L, except for Paracetamol-D4 and Paraxanthine-D6, which had final concentrations of 10µg/L (SI 1.2).

A twelve-point calibration in Evian mineral water, ranging from 10 ng/L to 40 µg/L, was prepared from four ethanol-based working standard solutions. For acetaminophen, this range was extended with four additional calibration points, reaching up to 500 µg/L.

For each measurement batch, three independently prepared samples were analyzed to determine the precision of the analytical method. Additionally, we spiked aliquots of a specific wastewater sample with the target analytes at seven different concentration levels, ranging from 50 ng/L to 10 µg/L, to assess analyte recoveries in the sample matrix (SI 1.5).

### LC-HRMS Measurements

The wastewater samples were analyzed for the target substances using a large volume direct injection into a reversed-phase liquid chromatography system coupled to a high-resolution mass spectrometer (LC-HRMS). The chromatographic system comprised a PAL auto-sampler (CTC Analytics, Switzerland) and a Dionex UltiMate3000 RS pump (Thermo Fisher Scientific, USA). A 100 µL sample was injected onto a reversed-phase C18 column (Atlantis® T3 3 µm, 3.0 × 150 mm; Waters). Chromatographic separation was conducted at a flow rate of 300 µL/min using a mobile phase gradient, starting with 100% eluent A (water with 0.1% v/v formic acid). After 1.5 minutes, eluent B (methanol with 0.1% v/v formic acid) was ramped up to 95% over 17 minutes, held for 10 minutes, and then returned to the initial conditions, followed by a 4-minute re-equilibration phase.

The samples were analyzed on a hybrid quadrupole-orbitrap high-resolution mass spectrometer (Orbitrap ExplorisTM 240 from Thermo Fisher Scientific) operating in positive ionization mode (ESI, 3.5 kV). Full scans were recorded at a resolution of 120,000 (at m/z 200), followed by five data-dependent MS/MS experiments at a resolution of 30,000 (at m/z 200). Fragmentation of target ions was based on an inclusion mass list and was performed using higher-energy collision-induced dissociation (HCD) with stepped collision energies of 15, 45, and 90%.

The HRMS data were analyzed using TraceFinder 5.1 (Thermo Fisher, USA). Target analytes were quantified from the extracted ion chromatogram of the MS full-scan, based on the area ratio of the reference standard (STD) to the corresponding ISTD of the analyte. Detailed information on the LC-HRMS settings and measurement quality control is provided in SI1.4 and SI2.1.

### Population-Normalized WWastewater daily population Loads

The concentrations measured in the wastewater samples (*c*_*sample*_, chemical analytes in milligrams per liter [mgL^-1^], viral RNA in gene copies per liter [gcL^-1^]) were multiplied by the daily wastewater volume (*Flow*_*WWTP*_, m^3^d^-1^). This product was then divided by the estimated population in the catchment area (*Pop*_*WWTP*_, p), resulting in population-normalized wastewater loads (DPL, mg d^-1^1000p^-1^ for chemical markers and gc d^-1^1000p^-1^ for viral loads), as described in Equation 1.

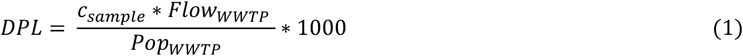

For Lugano WWTP, three data points were excluded due to unusually high loads, likely from sewer disposal or industrial sources.

### Swiss Sentinella Practice-Based Research Network

Data on weekly consultations for influenza-like illnesses and acute respiratory infection symptoms across Switzerland, along with associated laboratory testing results, were obtained from the Swiss Sentinella Network. The data is managed by the Swiss Federal Office of Public Health and is publicly available [39]. The Sentinella network comprises a volunteer cohort of 160 to 180 general practitioners, internists, and pediatricians who contribute data on the incidence of circulating respiratory virus infections across Switzerland [40,41].

Participating practitioners submit weekly reports on initial consultations and send nasopharyngeal swabs from cases to the National Reference Centre for Influenza (NRCI) for diagnostic analysis. The samples are tested for several respiratory viruses, including SARS-CoV-2, RSV, IAV, IBV, as well as other respiratory viruses such as adenovirus, rhinovirus, bocavirus, additional coronaviruses (229E, HKU1, NL63, OC43), metapneumovirus, and parainfluenza viruses (types 1-4).

We derived population-normalized estimates of specific respiratory infection cases by multiplying the test positivity rate by the population-normalized consultation incidence. The population-normalized consultation numbers (cases per 100‘000 inhabitants) were provided by the FOPH, based on the relative population coverage of the participating doctors within the Sentinella system.

### SARS-CoV-2 cases

SARS-CoV-2 case data were sourced from the Swiss mandatory reporting system for infectious diseases, overseen by the Federal Office of Public Health (FOPH). Catchment-level data for Zurich, Chur, Geneva, and Lugano based on postal codes corresponding to the catchment areas of the respective WWTPs. This data is publicly accessible for the duration of the study period.

### Wastewater Respiratory Virus and Pharmaceutical Load Modeling

The wastewater pharmaceutical loads (*DL*_*t*_) were modeled as a function of the viral gene copies of the key respiratory viruses SARS-CoV-2, RSV, IAV, IBV. The equation used is:

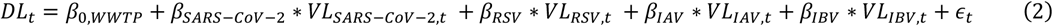

In this model:

- *β*_0,*WWTP*_ represents the WWTP-specific baseline mean pharmaceutical load during periods of low respiratory virus activity (June 2021 and July to August 2023), as determined from wastewater and sentinel surveillance data (Figure 2).
- *β*_1_, *β*_2_, *β*_3_, *β*_4_ are the coefficients for the viral loads of SARS-CoV-2, RSV, IAV, and IBV, respectively.
- *VL*_*SARS*−*CoV*−2,*t*_, *VL*_*RSV,t*_, *VL*_*IAV,t*_, *VL*_*IBV,t*_ denote the 7-day median viral gene copy loads for each respective virus at day *t*. For RSV, IAV, and IBV, this refers to copies of the Matrix protein gene (M), and for SARS-CoV-2, it refers to the Nucleoprotein gene locus 2 (N2), as detailed in [41].
- The error term *ϵ*_*t*_ *∼ N*(0,*σ*^2^) represents the unexplained variability in the model. It follows a normal distribution with a mean of 0 and a constant variance *σ*^2^. This term accounts for random fluctuations and factors not captured by the viral load variables.

We performed Ordinary Least Squares (OLS) regression to determine estimates of the coefficients (*β*_1_, *β*_2_, *β*_3_, *β*_4_), which quantify the contribution of virus-specific gene copy loads to the baseline-subtracted pharmaceutical load. A single OLS model was applied across all treatment plants, assuming a consistent relationship across locations. Diagnostic information for the OLS regressions is provided in SI2.4.1.

All analyses were conducted in Python 3.10 using the statsmodels 0.14.0 package for OLS regression. Access to code and computational environments used for evaluation is provided in the data and code availability statements.

## 4 Data Availability

The full dataset and corresponding analysis and visualization scripts will be available in Eawag’s Research Data Institutional Collection (ERIC-open) at https://doi.org/10.25678/000D6F.

## 6 Acknowledgments

The authors sincerely thank the operators of the wastewater treatment plants (WWTPs) involved in the Swiss wastewater surveillance program, whose essential support with sampling and data sharing made this research possible. We extend our gratitude to our colleagues at the University of Lausanne (UNIL), particularly Livia Andrani, Olivier Delémont, and Pierre Esseiva, for their assistance in coordinating sampling efforts. We also wish to express special thanks to Philipp Longrée for his expertise and support with the LC-HRMS analysis. Additionally, we thank Sentinella, the Swiss Sentinel Surveillance Network, and the Swiss Federal Office of Public Health (SFOPH) for providing surveillance data. This work was supported by SFOPH under contract numbers 142003899/321-446/1, 142004636/421-31/18, 142006108/334.0-101/26, and 142004331/421-31/15, as well as by the Swiss National Science Foundation through grant CRSII5_205933.

## 7 Author Contributions

Conceptualizations: S.B., H.S., C.O., T.R.J.

Methodology; S.B., M.S., H.S., C.O.

Measurements: S.B., M.S.

Data Analysis: S.B., M.S., P.S.

Drafting of Manuscript; S.B.

Critical revisions to the draft; T.R.J., C.O., H.S., P.S.

Completion and approval of final submission: S.B., M.S., T.R.J., P.S., C.O., H.S.

