## Supporting Information for "Parallel Analysis of Pharmaceuticals and Viruses in Wastewater: An Untargeted Approach to Tracking Population-Level Respiratory Illness Burden"

### Contents

SI 1 Materials and methods ... 3
SI 1.1 Wastewater treatment plant (WWTP) characteristics and wastewater sampling procedure ... 3
SI 1.2 Chemicals and solutions ... 8
SI 1.3 Chemical marker characteristics ... 9
SI 1.4 Analytical instrumentation and method ... 10
SI 1.5 Quantification method and quality control ... 10
SI 1.5.1 Quantification ... 10
SI 1.5.2 Matrix effects ... 10
SI 1.5.3 Limit of quantification ... 11
SI 1.5.4 Accuracy ... 11
SI 1.5.5 Precision ... 11
SI 2 Results and discussion ... 12
SI 2.1 LCMS quantification ... 12
SI 2.2 Spatiotemporal patterns of chemical and virus loads in wastewater ... 16
SI 2.2.1 Spatiotemporal pattern of baseline-subtracted loads for pharmaceuticals ... 20
SI 2.2.2 Influenza subtype clinical surveillance data ... 21
SI 2.2.3 Temporal trends of cases from mandatory reporting system and sentinel consultations ... 22
SI 2.2.4 Distribution of respiratory viruses from sentinel clinical testing ... 23
SI 2.3 Correlation analysis of wastewater markers ... 23
SI 2.3.1 Pearson correlations chemical and virus loads (location-specific) ... 23
SI 2.3.2 Pearson correlations chemical and virus loads (location independent) ... 24
SI 2.3.3 Pearson correlations baseline-subtracted chemical and virus loads (location independent) ... 24
SI 2.3.4 Scatter plot of viral and pharmaceutical loads (location independent) ... 25
SI 2.4 Regression analyses of pharmaceutical loads in wastewater and viral exposure ... 29
SI 2.4.1 Ordinary least squares (OLS) analyses of pharmaceutical and viral loads in wastewater ... 29
SI 2.4.2 Ordinary least squares analyses with clinical cases as predicting variables ... 41
SI 3 In-sample stability of chemical markers ... 47
SI 3.1 Sample preparation and analytical procedure ... 47
SI 3.2 Data analysis ... 47
SI 3.3 Stability results ... 47
SI 4 Licensed pharmaceutical products in Switzerland containing target chemicals ... 53
SI 5 References ... 54

### SI 1 Materials and methods

#### SI 1.1 Wastewater treatment plant (WWTP) characteristics and wastewater sampling procedure

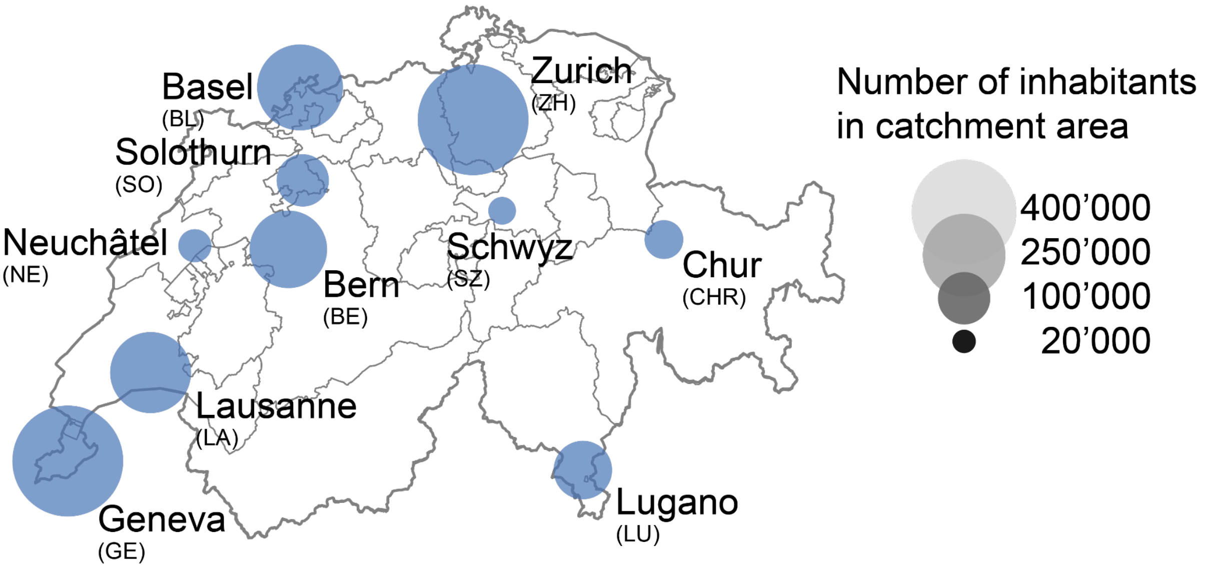


Figure SI 1: Geographic distribution of wastewater treatment plants (WWTPs) included in the study. The location of each WWTP is indicated by the center of a dot, with the dot size proportional to the catchment population served by the respective plant. The selection encompasses WWTPs from the different linguistic regions, including German-speaking (ZH, BL, BE, SO, CHR, SZ), French-speaking (GE, LA, NE), and Italian-speaking (LU) areas, representing both rural and urban environments.

Table SI 1: Characteristics of wastewater treatment plants (WWTPs) and details of the wastewater sampling procedures, as reported by WWTP operators.
(PE**:** Population Equivalent**,** n: count)

| **Location** | **Zurich** | **Basel** | **Bern** | **Solothurn** | **Chur** | **Schwyz** | **Geneva** | **Lausanne** | **Neuchâtel** | **Lugano** |
|---|---|---|---|---|---|---|---|---|---|---|
| **Name of the Wastewater Treatment Plant** | ARA Werdhölzli | ProRheno AG | ara region bern ag | ARA Emmenspitz | ARA Chur | ARA-Schwyz | STEP Aïre | STEP EPURA | STEP Neuchâtel | IDA-Bioggio |
| **Date of Processing** | 31.05.2023 | 03.01.2023 | 15.02.2023 | 26.1.2023 | 27.12.2023 | 26.01.2023 | 10.02.2023 | 16.05.2023 | 10.7.2024 | 10.02.2023 |
| **Residential population [p]** | 471275 | 273075 | 224557 | 95793 | 53123 | 31164 | 454433 | 247824 | 40697 | 123240 |
| **Date of Census** | 2022 | 2022 | 31.12.2021 | 1.1.2022 | - | 1.Januar 2022 | 31.12.2022 | 31.12.2022 | 1.1.2024 | 31.12.2021 |
| **Expansion Size [PE]** | 670000 | 477000 | 500000 | 125000 | 110000 | 70000 | 600000 | 350000 | 65000 | 186700 |
| **Current Total Load [PE]** | 693914 | 270000 | 400000 | 105000 | 75000 | 52000 | 696000 | 276612 | 47500 | 124416 |
| **Loads from Industrial Sources [PE]** | | 220000 | 103000 | - | 10000 | - | - | - | - | 11000 |
| **Most reliable number residential population** | Census | Census | Census | Census | Census | Census | Census | Census | - | Census |
| **Weekday population fluctuations** | Yes | No | Yes | No | No | No | Yes | - | No | Yes |
| **Increase during workdays** | Increase in people and discharge | - | Increase in people and discharge | - | - | - | Increase in people | - | - | Increase in people and discharge |
| **Number of hospitals in catchment** | 14 | 16 | 10 | >1 | >1 | 1 | 7 | 6 | 2 | >1 |
| **Hospital beds of all the hospitals listed above [n]** | 3665 | 2331 | Last survey in 2010: 2216 beds | - | - | 121 | Unknown | 1732 | - | - |
| **pump stations in catchment [n]** | 38 | 3 | 47 | 14 | 5 | 14 | 33 | 2 | 15 | 25 |
| **Exfiltration** | No | Yes | - | Yes | - | Yes | No | Yes | No | Yes |
| **Max. flow distance [km]** | 15 | 13 | - | 20 | 20 | 11 | 26.31 | 17 | 13 | 15 |
| **Average hydraulic sewer residence time [h]** | 1 | 1.5 | - | 3 | 5 | 1.2 | 3 | 0.66 | 1.5 | 3 |
| **Location inflow measurement** | After sand trap | After sand trap | After the sand trap | In effluent | After screen | Before pumping station. | Inflow | - | Before screen | After screen (4 inluents) |
| **Inflow measurement method** | Venturi | Radar (Endress+Hauser FMR60) | MID | Venturi | Venturi | Venturi | Water level and velocity | - | Water level and velocity | Venturi, MID |
| **Flow meter calibration interval [months]** | 0 | 0 | 12 | 6 | 12 | 0 | 0 | - | 0 | 12 |
| **Flow measurement accuracy estimation [%]** | - | 1 | ± 2 % | - | 5 | - | 5 | - | 10 | 2 |
| **Flow measuremnt accuracy manufacturer [%]** | 10% | 1% | -0.88% | 5 | 5 | 5-10 | unknown | - | 5 | 2 |
| **Lifting pump station operation** | Continuously | Continuously | None | Continuously | None | Continuously | None | None | - | None |
| **Wastewater volume on dry weather day [$m^3d^{-1}$]** | 143655 | 173000 | 63303 | 52000 | 13000 | 12000 | 132732 | - | 10500 | 35000 |
| **Minimum flow on dry weather day [$Ls^{-1}$]** | 750 | 500 | 265 | 500 | 40 | 83 | 700 | - | 75 | 250 |
| **Maximum flow on dry weather days [$Ls^{-1}$]** | 3000 | 1000 | 1165 | 1000 | 250 | 242 | 2300 | - | 200 | 600 |
| **Maximum flow on rainy weather day [$Ls^{-1}$]** | 6000 | 2000 | 3000 | 4500 | 550 | 1500 | 12000 | - | 1000 | 2500 |
| **Maximum threshold volume considered for evaluation [$m^2 d^{-1}$] (*empirically determined)** | 420000 | 120000 | 200000 | 130000 | 40000 | 70000 | 350000 | 150000 | 35000 | 150000 |

| | | | | | | | | | | |
|---|---|---|---|---|---|---|---|---|---|---|
| **Sampling location** | After the fine screen | After sand trap (until July 2022), then after fine screen | After fine screen | After sand trap | After fine screen | After fine screen | Before coarse screen. | - | Between coarse screen and sand trap | Before coarse screen |
| **Manufacturer and type of sampling device** | WaterSam WS316 MS3 | Bühler BU4011 | Bühler 4010 | Gerber WS 312 | Gerber | Endress&Hauser CSF48-MD50/0 | Endress&Hauser CSF48 | Teledyne ISCO | Hach Bühler 4011 | Endress&Hauser CSF48 |
| **Temperature in sampling device [°C]** | 4°C or cooler | 4°C or cooler | 4°C or cooler | 4°C or cooler | 4°C or cooler | 4°C or cooler | 4°C or cooler | 4°C or cooler | 4°C or cooler | 4°C or cooler |
| **Time until sample freezing [h]** | 7 | 1 | 0.75 | 1 | 2 | 0 | 0.5 | 2 | 8 | 0.5 |
| **Bottles in sampler [n]** | 4 | 4 | 1 | 4 | 4 | 1 | 1 | 1 | 4 | 4 |
| **Total volume in sampler [L]** | 72 | 40 | 30 | 48 | 25 | 30 | 20 | 20 | 10 | 48 |
| **Composite sample duration [h]** | 24 | 24 (48-72 on weekend/holidays) | 24 | 24 | 24 | 24 | 24 | 24 | 24 | 24 |
| **Material of sampling bottles** | PE | Plastic | PE-LD | Plastic | - | Plastic | PEHD | Plastic | Plastic | PEHD |
| **Sampling mode** | Volume-proportional | Flow-proportional | Volume-proportional | Volume-proportional | Volume-proportional | Volume-proportional | Volume-proportional | Time-proportional | Volume-proportional | Volume-proportional |
| **Sampling interval** | 800 $m^3$ | 10 min | - | 15 $m^3$ | 90 $m^3$ | 60 $m^3$ | 1460 $m^3$ | 5 min | 175 $m^3$ | 500 $m^3$ |
| **Storage temperature for collected sample at WWTP [°C]** | -20 | -20 | -20 | -20 | -20 | -20 | -20 | -20 | -20 | -20 |

Maximum threshold flow volumes were determined based on dates when pharmaceutical loads were systematically under- or overestimated due to high inflows associated with heavy rainfall events. These events likely led to the discharge of untreated wastewater, resulting in the underestimation of pharmaceutical loads and introducing uncertainty in both sampling and flow measurement at the treatment plant. Consequently, for wastewater samples collected on days when the inflow volume at the treatment plant exceeded the maximum threshold volume specified in Table SI 1, the corresponding pharmaceutical load data were excluded from analysis due to the associated uncertainties. In contrast, viral gene copy load data from these dates were still considered reliable, as they represent 7-day centered median values, which reduce the impact of such outliers.

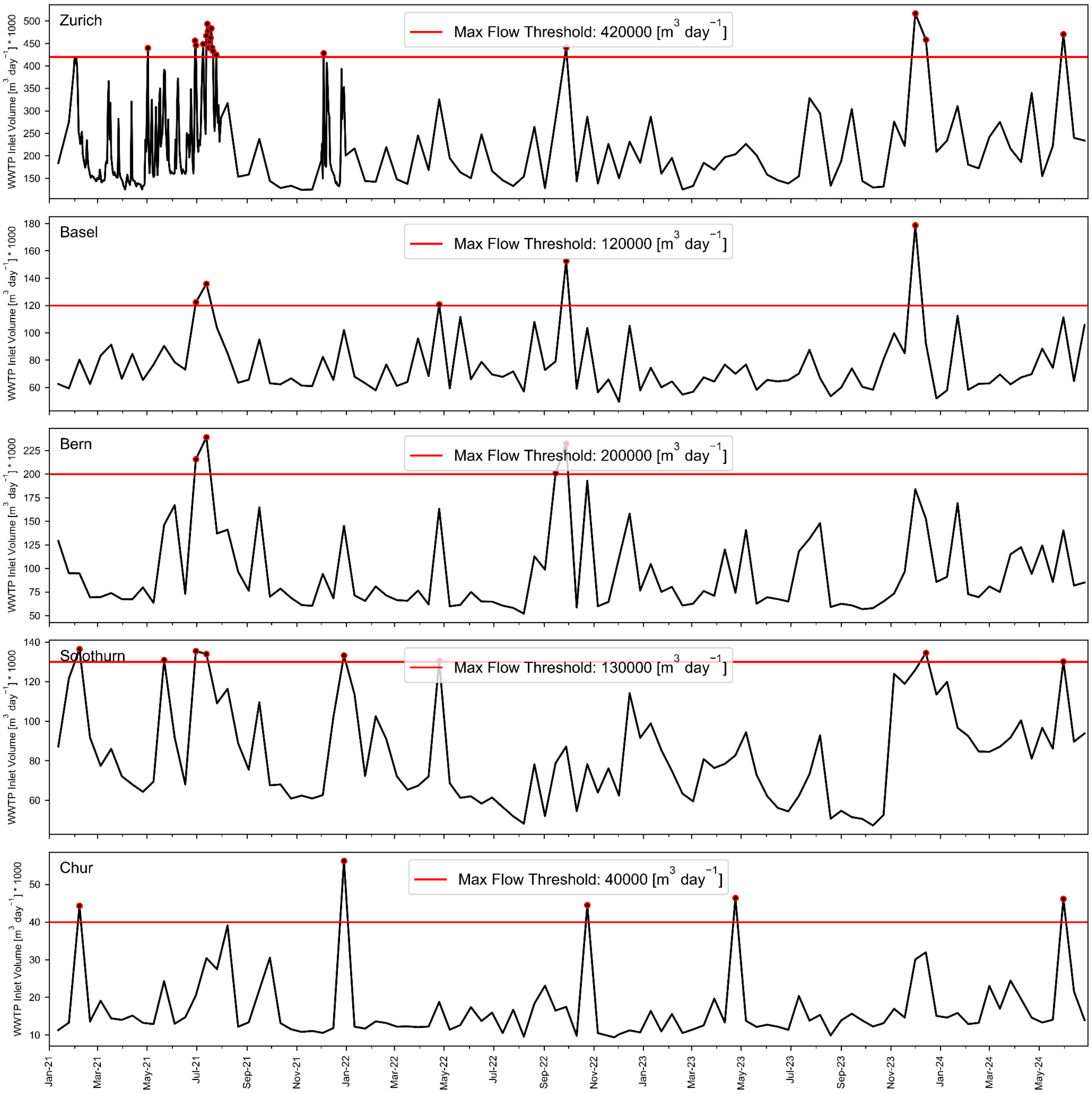


Figure SI 2: Daily wastewater inlet volumes at wastewater treatment plants (WWTPs). Black lines represent the daily inlet volumes on the days samples were collected for chemical analysis. The red line denotes the empirically determined, location-specific maximum volume threshold, above which daily load estimates are considered unreliable due to the discharge of untreated wastewater and potential irregularities in flow measurement and sampling. Red dots indicate the days when inlet volumes exceeded this threshold; data from these days were excluded from further analysis in the study.

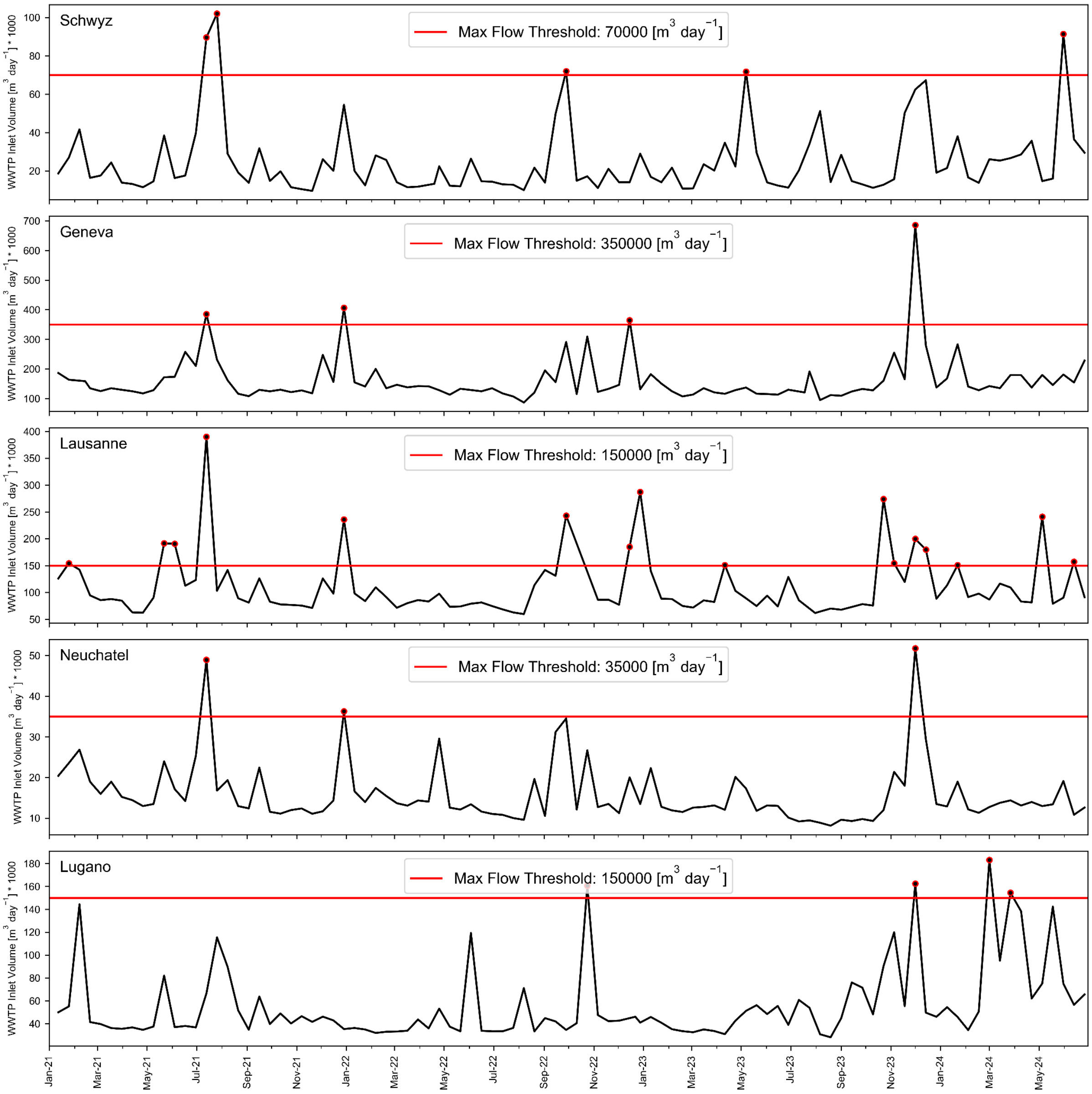


Figure SI 3: Daily wastewater inlet volumes at wastewater treatment plants (WWTPs). Black lines represent the daily inlet volumes on the days samples were collected for chemical analysis. The red line denotes the empirically determined, location-specific maximum volume threshold, above which daily load estimates are considered unreliable due to the discharge of untreated wastewater and potential irregularities in flow measurement and sampling. Red dots indicate the days when inlet volumes exceeded this threshold; data from these days were excluded from further analysis in the study.

#### SI 1.2 **Chemicals and solutions**

Organic solvents were of HPLC grade purity (> 98%) and supplied by Merck (ethanol) or Fisher Chemical (methanol and acetonitrile). Ultrapure water was obtained from a laboratory purification system (Arium Pro, Sartorius). Concentrated formic acid (>98%) was supplied by Merck.

Stock solutions of the individual analytes (STDs & ISTDs) were prepared at a concentration of 1 g/L in ethanol, methanol, or in acetonitrile (ACN) as described in Table SI 2. Individual stock solutions were combined into working mix solutions of 500, 60, 6, and 0.6 µg/L in EtOH. For acetaminophen, an additional working solution of 6 mg/L in ethanol was used for the high calibration range, extending up to 5000 µg/L. The isotope-labeled standard mix was prepared in ethanol at a concentration of 60 µg/L, with acetaminophen-D4 and paraxanthine-D6 included at a higher concentration of 600 µg/L. All solutions were stored at -20°C.

Table SI 2: Analytical reference standards (STD) and corresponding isotope-labelled standards (ISTD)

| Substance | CAS-No | Molecular formula | Manufacturer | Solvent | Conc. [g/L] | Corresponding ISTD |
|---|---|---|---|---|---|---|
| Dextrorphan | 125-73-5 | C17H23N1O1 | Lipomed AG | MeOH | 1 | Dextrorphan-D3 |
| Codeine | 76-57-3 | C18H21N1O3 | Lipomed AG | MeOH | 1 | Codeine-D6 |
| Morphine | 57-27-2 | C17H19N1O3 | Lipomed AG | MeOH | 1 | Morphine-D3 |
| Tramadol | 27203-92-5 | C16H25N1O2 | Sigma (*HCl) | EtOH | 1 | Tramadol-D6 |
| Diclofenac | 15307-86-5 | C14H11N1O2Cl2 | Sigma (*Na) | EtOH:H2O (1:1) | 1 | Diclofenac-D4 |
| Naproxen | 22204-53-1 | C14H14O3 | TRC-Canada | MeOH | 1 | Naproxen-D3 |
| Paracetamol (Acetaminophen) | 103-90-2 | C8H9N1O2 | Cerilliant | MeOH | 1 | Paracetamol-D4 |
| Pheniramine | 86-21-5 | C16H20N2 | Sigma-Aldrich (*maleate) | EtOH | 1 | Pheniramine-D6 |
| N-desmethylpheniramine | 19428-44-5 | C15H18N2 | LGC (*maleate) | MeOH | 1 | Pheniramine-D6 |
| Clarithromycin | 81103-11-9 | C38H69N1O13 | TCI EUROPE | EtOH | 1 | Clarithromycin-D3 |
| Metronidazole | 443-48-1 | C6H9N3O3 | Sigma-Aldrich | EtOH | 1 | Metronidazol-D4 |
| Sulfamethoxazole | 723-46-6 | C10H11N3O3S1 | Sigma-Aldrich | EtOH | 1 | Sulfamethoxazol-D4 |
| Trimethoprim | 738-70-5 | C14H18N4O3 | Sigma-Aldrich | EtOH | 1 | Trimethoprim-D3 |
| Candesartan | 139481-59-7 | C24H20N6O3 | TRC-Canada | ACN | 1 | Candesartan-D5 |
| Paraxanthine | 611-59-6 | C7H8N4O2 | Cerilliant | MeOH | 1 | Paraxanthine-D6 |
| Dextrorphan-D3 | - | C17H20[2]H3N1O1 | Lipomed AG | MeOH | 1 | - |
| Codeine-D6 | 1007844-34-9 | C18H15[2]H6N1O3 | Lipomed AG | MeOH | 1 | - |
| Morphine-D3 | 67293-88-3 | C17H16[2]H3N1O3 | Sigma-Aldrich | MeOH | 1 | - |
| Tramadol-D6 | - | C16H19[2]H6N1O2 | TRC-Canada (*HCl) | MeOH | 1 | - |
| Diclofenac-D4 | 153466-65-0 | C14H7[2]H4N1O2Cl2 | TRC-Canada | MeOH | 1 | - |
| Naproxen-D3 | | C14H11[2]H3O3 | C/D/N Isotopes Inc. | ACN | 1 | - |
| Paracetamol-D4 (Acetaminophen-D4) | 64315-36-2 | C8H5[2]H4N1O2 | TRC-Canada | EtOH | 1 | - |
| Lidocaine-D10 | 1189959-13-4 | C14H12[2]H10N2O1 | TRC-Canada (*HCl) | EtOH | 1 | - |
| Pheniramine-D6 | | C16H14[2]H6N2 | TRC-Canada | MeOH | 1 | - |
| Clarithromycin-D3 | 959119-17-6 | C38H66[2]H3N1O13 | TRC-Canada | MeOH | 0.5 | - |
| Metronidazol-D4 | 1261392-47-5 | C6H5[2]H4N3O3 | TRC-Canada | EtOH | 1 | - |
| Sulfamethoxazol-D4 | 1020719-86-1 | C10H7[2]H4N3O3S1 | TRC-Canada | ACN | 1 | - |
| Trimethoprim-D3 | 1189923-38-3 | C14H15[2]H3N4O3 | TRC-Canada | MeOH | 1 | - |
| Candesartan-D5 | 1189650-58-5 | C24H15[2]H5N6O3 | TRC-Canada | MeOH | 1 | - |
| Paraxanthine-D6 | 117490-41-2 | C7H2[2]H6N4O2 | Cayman Chemical | MeOH | 0.1 | - |

For the quantification of Pheniramine and N-desmethylpheniramine using LC-MS, the isotope-labeled internal standard was changed from Lidocaine-D10 to Pheniramine-D6 during the course of the project. Consequently, samples collected from April 11, 2023, onwards in Bern, Geneva, Lausanne, and Neuchâtel, and from July 11, 2023, onwards in Zurich, Basel, Solothurn, Chur, Schwyz, and Lugano were quantified using Pheniramine-D6. Samples collected before these dates were quantified using Lidocaine-D10. To account for potential deviations in absolute concentrations, relative recoveries were determined. Additionally, aliquots of spiked wastewater and mineral water (referred to as 'eternity samples') were quantified in each measurement batch to ensure consistency over time, as detailed in the long-term stability analysis (SI 3.3).

SI 1.3 **Chemical marker characteristics**

#### Opioid analog

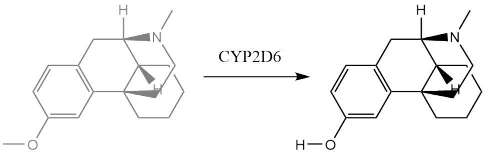


Dextromethorphan

**Dextrorphan**
(metabolite)

#### Opioids

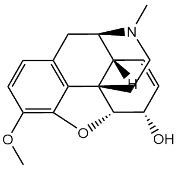

**Codeine**

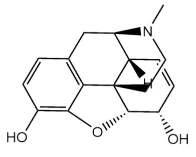

Morphine

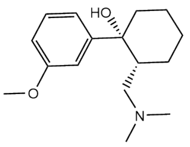

Tramadol

#### Non-opioid analgesic

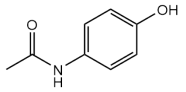

**Acetaminophen / Paracetamol**

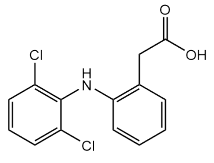

Diclofenac
(NSAID)

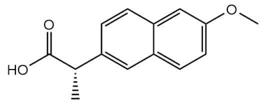

Naproxen
(NSAID)

#### Antihistamines

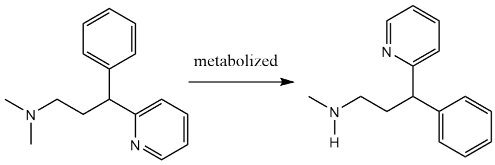


**Pheniramine**
(1st generation antihistamine)

**N-desmethylpheniramin**e
(metabolite)

#### Antibiotics

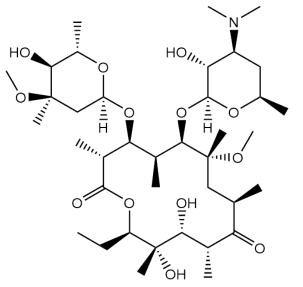

**Clarithromycin**
(macrolide)

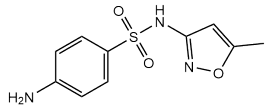

Sulfamethoxazole
(sulfonamide)

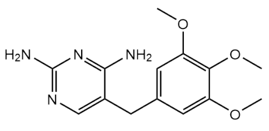

Trimethoprim
(antifolate)

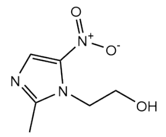

Metronidazole
(nitromidazole)

#### Controls

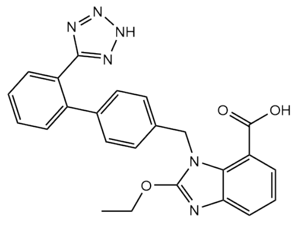

Candesartan
(angiotensin-receptor blocker)

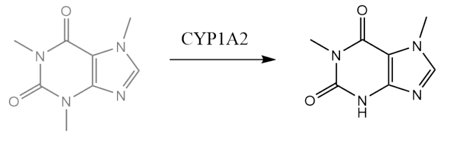


Caffeine

Paraxanthine
(metabolite)

Figure SI 4: Molecular structures of small molecule wastewater markers. Analytes highlighted in bold demonstrated a positive correlation with viral RNA gene copy loads of respiratory viruses, including SARS-CoV-2, Influenza A (IAV), and Respiratory Syncytial Virus (RSV) in wastewater samples.

#### SI 1.4 **Analytical instrumentation and method**

Table SI 3: Parameter settings used for HRMS/MS measurements with Exploris$^{TM}$ 240 mass spectrometer

| Parameter | Value |
|---|---|
| Ionization mode | positive |
| Spray voltage [kV] | 3.5 |
| Ion transfer tube temperature [°C] | 320 |
| Sheet gas flow (nitrogen) [AU] | 40 |
| Auxiliary gas flow (nitrogen) [AU] | 10 |
| Sweep gas flow [AU] | 0 |
| RF Lens [%] | 60 |
| Spectrum data type | Profile |
| Maximum injection time MS1 [ms] | 100 |
| Scan range MS1 [m/z] | 100 - 1000 |
| Resolution MS1 | 120000 |
| Internal calibration | Yes (Run Start EASY-IC) |
| Data-dependent mode | Number of scans |
| Data-dependent trigger | Ions of mass list |
| Isolation window [m/z] | 1 |
| Resolution MS/MS | 30000 |
| Collision energy mode | Stepped |
| Collision energy type | Normalized |
| HCD collision energies [%] | 15, 45, 90 |

AU: arbitrary units
ms: milliseconds
m/z: mass to charge ratio

#### SI 1.5 **Quantification method and quality control**

##### SI 1.5.1 **Quantification**

Target analytes were quantified based on the area ratio of the analyte and its selected isotope-labelled internal standard (ISTD) using Tracefinder 5.1 software. Structurally identical ISTDs were available for all analytes, except for the metabolie N-desmethylpheniramine. The target peaks were extracted from full scans with a mass tolerance of 5 ppm, and only peaks with at least five data points and a signal-to-noise ratio greater than 10 were considered for quantification. Extracted peaks were verified as target analytes by comparing the MS/MS spectra, retention time, and isotopic pattern to those of the corresponding reference standards and library spectra from MZCloud. The twelve-point calibration in Evian mineral water included the following concentration levels: 10, 25, 50, 100, 250, 500, 1000, 2500, 5000, 10000, 25000, 40000ng/L. For acetaminophen the calibration extended to 100, 250, 500 µg/L.
Linear calibration was applied to all substances, with regression weighting set to 1/x.
All quality control parameters of the LC-HRMS measurements that are described below were determined for each measurement batch independently.

##### SI 1.5.2 **Matrix effects**

The impact of matrix effects during analysis was evaluated by determining the absolute recoveries specific to each analyte. For target analytes with structurally identical ISTD, the absolute recovery is described as the ratio of the ISTD area in the matrix samples ($ISTD\ Area_{Matrix}$) to their average area in the calibration series ($Average\ ISTD\ Area_{Calibration}$), as described in equation SI 1:

$$Absolute\ Recovery = \frac{ISTD\ Area_{Matrix}}{Average\ ISTD\ Area_{Calibration}} \qquad (SI\ 1)$$

If a structurally identical ISTD was unavailable, the absolute recovery was calculated based on the difference of the target peak area in spiked samples ($STD\ Area_{spiked\ sample}$) and the area in the unspiked sample ($STD\ Area_{unspiked\ sample}$) and set into relation with the area in the respective calibration standard ( $STD\ Area_{Calibration}$) (Equation SI 2).

$$Absolute\ recovery_{non-identical\ ISTD} = \frac{STD\ Area_{spiked\ sample} - STD\ Area_{unspiked\ sample}}{STD\ Area_{corresponding\ calibration\ standard}} \qquad (SI\ 2)$$

##### SI 1.5.3 **Limit of quantification**

The limit of quantification (LOQ) was determined based on the lowest concentration in the wastewater matrix, at which the quantification criteria (i. signal-to-noise ratio larger than 10 and ii. at least five data points) was met. If such an LOQ in the wastewater matrix was not determinable, the LOQ was determined based on the lowest quantified calibration level multiplied by the inverse of the absolute recovery (Equation SI 3).

$$LOQ_{matrix}\ [ngL^{-1}] = \frac{LOQ_{solvent}}{Absolute\ recovery} \qquad (SI\ 3)$$

Additionally, if the target analyte was detected in the matrix blanks (Evian mineral water spiked with ISTD and processed as the samples) an LOQ based on the equation below, with sd being the standard deviation, was calculated (Equation SI 4).

$$LOQ_{matrix\ blank}[ngL^{-1}] = Average\ C_{matrix\ blank} + 10 * sd(C_{matrix\ blanks}) \qquad (SI\ 4)$$

If multiple LOQ values were obtained the higher value was considered.

##### SI 1.5.4 **Accuracy**

The accuracy of the measurements was assessed by determining the relative recovery in wastewater samples that were deliberately spiked with specific concentrations as described in the equation below (Equation SI 5). In each measurement batch, a wastewater sample was spiked with the target analytes at the following concentrations; 50, 100, 250, 500, 1000, 2500, 5000, 10000ng/L.

$$Relative\ recovery\ [\%] = \frac{C_{spiked\ sample} - C_{unspiked\ sample}}{C_{spiked}} * 100 \qquad (SI\ 5)$$

Relative recoveries were only calculated if the concentration in the unspiked sample was lower than twice the spiked concentration. If the concentration in the unspiked sample was lower than twice the LOQ only recoveries from samples with a spiked concentration above four times the LOQ were reported. Recoveries were not considered if the total concentration in the spiked sample exceeded the highest calibration point concentration.

##### SI 1.5.5 **Precision**

Three aliquots of the same wastewater sample were prepared and analyzed separately in every measurement batch to assess the overall precision of the analytical method.

The precision was computed as the relative standard deviation of the three measurements (Equation SI 6).

$$Relative\ standard\ deviation\ [\%] = \frac{std\ (C_{Replicates})}{mean\ (C_{Replicates})} * 100 \qquad (SI\ 6)$$

### SI 2 Results and discussion

#### SI 2.1 **LCMS quantification**

Table SI 4: Summary of LCMS Analytics Quality Control and Quantification. The data are presented as average values for individual measurement batches. Quality control parameters determined as described above. (m/z: Mass to charge ratio, RT: Retention time, LOQ: Limit of quantification, SD: Standard deviation, ND: Not determinable)

| Substance | Measurement batch ID | m/z theoretical [M+H]+I | m/z delta [ppm] | RT measured [min] | SD RT [min] | LOQ [ng/L] | Sample conc. [ng/L] | Samples above LOQ [%] | Absolute recovery | Relative recovery [%] | Precision [%] | Number of samples |
|---|---|---|---|---|---|---|---|---|---|---|---|---|
| Acetaminophen | 220613 | 152.0706 | 0.46 | 10.4 | 0.02 | 500 | 68428 | 99.3 | 0.51 | ND | 7.5 | 148 |
| Acetaminophen | 220623 | 152.0706 | 0.28 | 10.5 | 0.01 | 500 | 88752 | 100 | 0.6 | ND | 2.2 | 38 |
| Acetaminophen | 220704 | 152.0706 | 0.44 | 10.4 | 0.02 | 500 | 27054 | 97.2 | 0.63 | 120.2 | 9.7 | 143 |
| Acetaminophen | 220803 | 152.0706 | 0.20 | 10.5 | 0.01 | 500 | 28486 | 100 | 0.57 | ND | 2.7 | 81 |
| Acetaminophen | 221021 | 152.0706 | 0.06 | 10.2 | 0.02 | 500 | 23838 | 100 | 0.63 | 89.2 | 4.9 | 187 |
| Acetaminophen | 230109 | 152.0706 | 0.38 | 9.9 | 0.02 | 500 | 45478 | 99.4 | 0.57 | 125.6 | 3.2 | 181 |
| Acetaminophen | 230424 | 152.0706 | 0.20 | 10 | 0.01 | 500 | 40199 | 100 | 0.45 | 73.6 | 3.9 | 45 |
| Acetaminophen | 230711 | 152.0706 | -0.17 | 10.3 | 0.01 | 500 | 64418 | 99 | 0.76 | 89.9 | 7.1 | 100 |
| Acetaminophen | 231106 | 152.0706 | 1.06 | 10.3 | 0.03 | 500 | 27980 | 100 | 0.51 | ND | 0.3 | 45 |
| Acetaminophen | 240131 | 152.0706 | 0.35 | 10.3 | 0.01 | 500 | 58097 | 100 | 0.57 | ND | 2.2 | 127 |
| Acetaminophen | 240701 | 152.0706 | 1.01 | 10.6 | 0.06 | 500 | 63367 | 99.3 | 0.62 | 115.4 | 1.9 | 143 |
| Candesartan | 220613 | 441.167 | 0.53 | 18.8 | 0.05 | 50 | 476 | 100 | 0.69 | 108.2 | 5.4 | 148 |
| Candesartan | 220623 | 441.167 | 0.47 | 18.8 | 0.04 | 50 | 521 | 97.4 | 0.71 | 105.3 | 4 | 38 |
| Candesartan | 220704 | 441.167 | 0.42 | 18.8 | 0.06 | 50 | 478 | 99.3 | 0.76 | 100 | 3.1 | 143 |
| Candesartan | 220803 | 441.167 | 0.08 | 18.7 | 0.03 | 50 | 345 | 100 | 0.73 | 100 | 1.8 | 81 |
| Candesartan | 221021 | 441.167 | 0.23 | 18.6 | 0.07 | 50 | 305 | 97.9 | 0.75 | 98.6 | 6.1 | 187 |
| Candesartan | 230109 | 441.167 | 0.50 | 18.3 | 0.04 | 50 | 483 | 100 | 0.65 | 100.7 | 1 | 181 |
| Candesartan | 230424 | 441.167 | 0.26 | 18.3 | 0.06 | 50 | 533 | 100 | 0.47 | 106.5 | 6.4 | 45 |
| Candesartan | 230711 | 441.167 | 0.04 | 18.5 | 0.04 | 50 | 514 | 100 | 0.6 | 108.7 | 3.2 | 100 |
| Candesartan | 231106 | 441.167 | 1.36 | 18.6 | 0.02 | 50 | 452 | 100 | 0.61 | 95.4 | 3.2 | 45 |
| Candesartan | 240131 | 441.167 | 0.06 | 18.7 | 0.01 | 50 | 486 | 100 | 0.73 | 102.3 | 4.5 | 127 |
| Candesartan | 240701 | 441.167 | -0.15 | 18.8 | 0 | 50 | 476 | 100 | 0.67 | 96.2 | 3.9 | 143 |
| Clarithromycin | 220613 | 748.4842 | 0.43 | 16.4 | 0.07 | 60 | 159 | 92.6 | 0.82 | 104.2 | 7.7 | 148 |
| Clarithromycin | 220623 | 748.4842 | 0.51 | 16.4 | 0.05 | 60 | 182 | 89.5 | 0.8 | 102.7 | 5.7 | 38 |
| Clarithromycin | 220704 | 748.4842 | 0.30 | 16.5 | 0.08 | 60 | 181 | 76.2 | 0.86 | 99.9 | 1.4 | 143 |
| Clarithromycin | 220803 | 748.4842 | 0.20 | 16.6 | 0.05 | 60 | 157 | 96.3 | 0.75 | 96.6 | 0.9 | 81 |
| Clarithromycin | 221021 | 748.4842 | 0.04 | 16.3 | 0.08 | 60 | 155 | 94.1 | 0.73 | 102 | 10.8 | 187 |
| Clarithromycin | 230109 | 748.4842 | 0.28 | 16.2 | 0.07 | 60 | 168 | 89 | 0.72 | 101.4 | 7 | 181 |
| Clarithromycin | 230424 | 748.4842 | 0.44 | 16.2 | 0.09 | 60 | 248 | 95.6 | 0.77 | 98.6 | 6.7 | 45 |
| Clarithromycin | 230711 | 748.4842 | 0.02 | 16.6 | 0.08 | 60 | 203 | 94 | 0.81 | 102.4 | 1.6 | 100 |
| Clarithromycin | 231106 | 748.4842 | 1.67 | 16.3 | 0.02 | 60 | 155 | 88.9 | 0.49 | 102.1 | 2.5 | 45 |
| Clarithromycin | 240131 | 748.4842 | 0.22 | 16.6 | 0 | 60 | 188 | 92.9 | 0.61 | 98.9 | 2.1 | 127 |
| Clarithromycin | 240701 | 748.4842 | 0.03 | 16.7 | 0.09 | 60 | 232 | 95.8 | 0.59 | 89.3 | 4 | 143 |
| Codeine | 220613 | 300.1594 | 0.47 | 9 | 0.02 | 45 | 196 | 95.3 | 0.86 | 104.2 | 6.3 | 148 |
| Codeine | 220623 | 300.1594 | 0.28 | 9 | 0.02 | 45 | 202 | 97.4 | 0.96 | 108.3 | 7.5 | 38 |
| Codeine | 220704 | 300.1594 | 0.28 | 9 | 0.03 | 45 | 148 | 75.5 | 0.88 | 106 | 3.1 | 143 |
| Codeine | 220803 | 300.1594 | 0.08 | 9.1 | 0.02 | 45 | 105 | 89.9 | 0.93 | 107.9 | ND | 79 |
| Codeine | 221021 | 300.1594 | 0.18 | 8.9 | 0.02 | 45 | 102 | 79.3 | 0.97 | 100.3 | ND | 184 |
| Codeine | 230109 | 300.1594 | 0.63 | 8.7 | 0.02 | 45 | 160 | 92.3 | 0.75 | 103.3 | 6.5 | 181 |
| Codeine | 230424 | 300.1594 | 0.31 | 8.7 | 0.02 | 45 | 158 | 93.3 | 0.82 | 101.9 | 5.9 | 45 |
| Codeine | 230711 | 300.1594 | 0.28 | 9 | 0.02 | 45 | 182 | 93.9 | 0.85 | 107.9 | 1.6 | 99 |

| Substance | Measurement batch ID | m/z theoretical [M+H]+I | m/z delta [ppm] | RT measured [min] | SD RT [min] | LOQ [ng/L] | Sample conc. [ng/L] | Samples above LOQ [%] | Absolute recovery | Relative recovery [%] | Precision [%] | Number of samples |
|---|---|---|---|---|---|---|---|---|---|---|---|---|
| Codeine | 231106 | 300.1594 | 1.61 | 8.8 | 0.03 | 45 | 228 | 80.4 | 0.88 | 93.4 | 2.3 | 46 |
| Codeine | 240131 | 300.1594 | 0.82 | 8.8 | 0.02 | 45 | 194 | 89 | 0.92 | 102.6 | 5.9 | 127 |
| Codeine | 240701 | 300.1594 | 0.23 | 9 | 0.04 | 45 | 160 | 88.7 | 0.79 | 100 | 6.7 | 142 |
| Dextrorphan | 220613 | 258.1852 | 0.43 | 11.8 | 0.03 | 10 | 71 | 96.6 | 0.65 | 107.2 | 7.4 | 148 |
| Dextrorphan | 220623 | 258.1852 | 0.26 | 11.9 | 0.02 | 10 | 62 | 94.7 | 0.77 | 107.1 | 4.5 | 38 |
| Dextrorphan | 220704 | 258.1852 | 0.17 | 11.9 | 0.04 | 10 | 54 | 88.1 | 0.69 | 107.2 | 2 | 143 |
| Dextrorphan | 220803 | 258.1852 | -0.11 | 12 | 0.03 | 10 | 51 | 96.2 | 0.69 | 105.9 | ND | 79 |
| Dextrorphan | 221021 | 258.1852 | 0.18 | 11.7 | 0.04 | 10 | 58 | 94 | 0.7 | 96.9 | ND | 184 |
| Dextrorphan | 230109 | 258.1852 | 0.50 | 11.5 | 0.04 | 10 | 77 | 99.4 | 0.63 | 103.3 | 5.1 | 181 |
| Dextrorphan | 230424 | 258.1852 | 0.32 | 11.5 | 0.04 | 10 | 98 | 100 | 0.72 | 98.4 | 4.5 | 45 |
| Dextrorphan | 230711 | 258.1852 | 0.17 | 12 | 0.04 | 10 | 81 | 99 | 0.72 | 108.6 | 1.3 | 99 |
| Dextrorphan | 231106 | 258.1852 | 1.51 | 11.6 | 0.03 | 10 | 46 | 95.7 | 0.8 | 92.7 | 1.5 | 46 |
| Dextrorphan | 240131 | 258.1852 | 0.91 | 11.7 | 0.04 | 10 | 63 | 100 | 0.72 | 97.6 | 2.2 | 127 |
| Dextrorphan | 240701 | 258.1852 | 0.66 | 12 | 0.07 | 10 | 64 | 99.3 | 0.68 | 90.2 | 1 | 142 |
| Diclofenac | 220613 | 296.024 | 0.14 | 20.6 | 0.01 | 100 | 1658 | 100 | 0.78 | 116 | 7.9 | 148 |
| Diclofenac | 220623 | 296.024 | 0.11 | 20.6 | 0.01 | 100 | 1236 | 97.4 | 0.8 | 107.3 | 3.1 | 38 |
| Diclofenac | 220704 | 296.024 | 0.09 | 20.6 | 0.02 | 100 | 1690 | 98.6 | 0.85 | 107.3 | 0.8 | 143 |
| Diclofenac | 220803 | 296.024 | -0.28 | 20.6 | 0.01 | 100 | 1624 | 100 | 0.84 | 104.4 | 8.7 | 81 |
| Diclofenac | 221021 | 296.024 | 0.11 | 20.4 | 0.02 | 100 | 1334 | 100 | 0.79 | 106.2 | 4.6 | 187 |
| Diclofenac | 230109 | 296.024 | 0.38 | 20.2 | 0.01 | 100 | 1542 | 100 | 0.88 | 98.3 | 4.2 | 181 |
| Diclofenac | 230424 | 296.024 | 0.16 | 20.2 | 0.02 | 100 | 1760 | 100 | 0.69 | 100.4 | 1.5 | 45 |
| Diclofenac | 230711 | 296.024 | -0.11 | 20.4 | 0.02 | 100 | 1359 | 100 | 0.88 | 101.2 | 1.1 | 100 |
| Diclofenac | 231106 | 296.024 | 1.41 | 20.5 | 0 | 100 | 1431 | 100 | 0.65 | 98.6 | 1.6 | 45 |
| Diclofenac | 240131 | 296.024 | 0.59 | 20.6 | 0 | 100 | 1252 | 100 | 0.92 | 101.9 | 2.6 | 127 |
| Diclofenac | 240701 | 296.024 | -0.14 | 20.5 | 0 | 100 | 1266 | 100 | 0.89 | 91.7 | 2.1 | 143 |
| Metronidazole | 220613 | 172.0717 | 0.33 | 10.1 | 0.02 | 25 | 194 | 97.3 | 0.52 | 109.8 | 6 | 148 |
| Metronidazole | 220623 | 172.0717 | 0.27 | 10.2 | 0.02 | 25 | 194 | 97.4 | 0.61 | 114.5 | 3.7 | 38 |
| Metronidazole | 220704 | 172.0717 | 0.13 | 10.2 | 0.02 | 25 | 148 | 97.9 | 0.53 | 106.9 | 1.2 | 143 |
| Metronidazole | 220803 | 172.0717 | -0.09 | 10.1 | 0.02 | 25 | 164 | 100 | 0.57 | 100.7 | 4.3 | 81 |
| Metronidazole | 221021 | 172.0717 | 0.07 | 9.9 | 0.02 | 25 | 145 | 100 | 0.55 | 105 | 2.5 | 187 |
| Metronidazole | 230109 | 172.0717 | 0.44 | 9.7 | 0.03 | 25 | 150 | 97.8 | 0.45 | 104.1 | 3.1 | 181 |
| Metronidazole | 230424 | 172.0717 | 0.27 | 9.7 | 0.02 | 25 | 161 | 100 | 0.47 | 102.9 | 1.2 | 45 |
| Metronidazole | 230711 | 172.0717 | 0.11 | 10.1 | 0.02 | 25 | 182 | 97 | 0.73 | 104.5 | 1.2 | 100 |
| Metronidazole | 231106 | 172.0717 | 1.27 | 10.1 | 0.02 | 25 | 150 | 100 | 0.59 | 100.6 | 1.7 | 45 |
| Metronidazole | 240131 | 172.0717 | 0.93 | 10.2 | 0.01 | 25 | 191 | 96.1 | 0.48 | 104 | 16.1 | 127 |
| Metronidazole | 240701 | 172.0717 | 1.24 | 10.3 | 0.04 | 25 | 172 | 97.2 | 0.54 | 105.2 | 6.2 | 143 |
| Morphine | 220613 | 286.1438 | 0.21 | 7.4 | 0.02 | 100 | 1046 | 98.6 | 1.04 | 85.8 | 15.7 | 148 |
| Morphine | 220623 | 286.1438 | 0.18 | 7.4 | 0.02 | 100 | 570 | 94.7 | 1.25 | 105.5 | 9.7 | 38 |
| Morphine | 220704 | 286.1438 | -0.01 | 7.4 | 0.02 | 100 | 359 | 67.1 | 0.98 | 90.3 | 6.4 | 143 |
| Morphine | 220803 | 286.1438 | -0.15 | 7.5 | 0.02 | 100 | 958 | 97.5 | 1.04 | 97.2 | ND | 79 |
| Morphine | 221021 | 286.1438 | 0.10 | 7.3 | 0.03 | 100 | 900 | 93.5 | 0.97 | 85.5 | ND | 184 |
| Morphine | 230109 | 286.1438 | 0.58 | 7.1 | 0.03 | 100 | 651 | 87.3 | 0.7 | 101.2 | 9.5 | 181 |
| Morphine | 230424 | 286.1438 | 0.26 | 7.2 | 0.01 | 100 | 613 | 91.1 | 0.84 | 93.6 | 10.6 | 45 |
| Morphine | 230711 | 286.1438 | -0.01 | 7.5 | 0.03 | 100 | 685 | 92.9 | 0.87 | 100.9 | 8 | 99 |
| Morphine | 231106 | 286.1438 | 1.43 | 7.2 | 0.05 | 100 | 628 | 91.3 | 0.96 | 90.4 | 2.4 | 46 |
| Morphine | 240131 | 286.1438 | 0.72 | 7.3 | 0.02 | 100 | 690 | 90.6 | 0.98 | 81.3 | 4 | 127 |
| Morphine | 240701 | 286.1438 | 0.24 | 7.4 | 0.04 | 100 | 624 | 85.9 | 0.87 | 79.2 | 3.3 | 142 |

| Substance | Measurement batch ID | m/z theoretical [M+H]+I | m/z delta [ppm] | RT measured [min] | SD RT [min] | LOQ [ng/L] | Sample conc. [ng/L] | Samples above LOQ [%] | Absolute recovery | Relative recovery [%] | Precision [%] | Number of samples |
|---|---|---|---|---|---|---|---|---|---|---|---|---|
| N-desmethylpheniramine | 220613 | 227.1543 | 0.40 | 10.5 | 0.17 | 20 | 42 | 42.6 | 0.79 | 98.2 | 10.1 | 148 |
| N-desmethylpheniramine | 220623 | 227.1543 | 0.15 | 10.8 | 0 | 20 | 39 | 42.1 | 0.97 | 108.2 | 9.8 | 38 |
| N-desmethylpheniramine | 220704 | 227.1543 | 0.18 | 10.8 | 0 | 20 | 41 | 32.9 | 0.9 | 107.8 | 3.7 | 143 |
| N-desmethylpheniramine | 220803 | 227.1543 | -0.03 | 10.5 | 0.04 | 20 | 28 | 34.6 | 0.79 | 98 | 3.4 | 81 |
| N-desmethylpheniramine | 221021 | 227.1543 | 0.35 | 10.2 | 0.04 | 20 | 46 | 36.9 | 0.82 | 100.2 | ND | 187 |
| N-desmethylpheniramine | 230109 | 227.1543 | 0.50 | 10.2 | 0.02 | 20 | 84 | 15.5 | 0.7 | 102.5 | ND | 181 |
| N-desmethylpheniramine | 230424 | 227.1543 | 0.41 | 10.2 | 0.02 | 20 | 53 | 82.2 | 0.89 | 101.7 | 11.6 | 45 |
| N-desmethylpheniramine | 230711 | 227.1543 | -0.08 | 10.5 | 0.04 | 20 | 39 | 58 | 0.95 | 99.9 | ND | 100 |
| N-desmethylpheniramine | 231106 | 227.1543 | 1.46 | 10.1 | 0.04 | 20 | 41 | 31.1 | 0.79 | 88.1 | 20.2 | 45 |
| N-desmethylpheniramine | 240131 | 227.1543 | 1.13 | 10.5 | 0.02 | 20 | 53 | 66.9 | 0.86 | 116.5 | 4.8 | 127 |
| N-desmethylpheniramine | 240701 | 227.1543 | 1.07 | 10.8 | 0.11 | 20 | 42 | 53.1 | 1.11 | 103.3 | 4.4 | 143 |
| Naproxen | 220613 | 231.1016 | 0.29 | 19.2 | 0.01 | 300 | 2038 | 99.3 | 0.51 | 119.8 | 6 | 148 |
| Naproxen | 220623 | 231.1016 | 0.23 | 19.2 | 0.01 | 300 | 1502 | 97.4 | 0.62 | 107.8 | 5.7 | 38 |
| Naproxen | 220704 | 231.1016 | 0.20 | 19.2 | 0.01 | 300 | 1577 | 97.2 | 0.66 | 109 | 4.1 | 143 |
| Naproxen | 220803 | 231.1016 | -0.15 | 19.2 | 0.01 | 300 | 1763 | 100 | 0.63 | 111.5 | 4.8 | 81 |
| Naproxen | 221021 | 231.1016 | 0.27 | 19 | 0.01 | 300 | 1539 | 98.9 | 0.58 | 93.2 | 7.7 | 187 |
| Naproxen | 230109 | 231.1016 | 0.59 | 18.8 | 0.01 | 300 | 1828 | 98.9 | 0.69 | 99.6 | 5.5 | 181 |
| Naproxen | 230424 | 231.1016 | 0.51 | 18.8 | 0.01 | 300 | 1857 | 100 | 0.46 | 106.2 | 3.5 | 45 |
| Naproxen | 230711 | 231.1016 | 0.25 | 19.1 | 0.02 | 300 | 1901 | 99 | 0.73 | 106.2 | 2.2 | 100 |
| Naproxen | 231106 | 231.1016 | 1.52 | 19 | 0.01 | 300 | 1527 | 100 | 0.5 | 88.9 | 3.5 | 45 |
| Naproxen | 240131 | 231.1016 | 1.22 | 19 | 0.01 | 300 | 1809 | 97.6 | 0.77 | 100.5 | 3 | 127 |
| Naproxen | 240701 | 231.1016 | 1.36 | 19.3 | 0.07 | 300 | 1714 | 98.6 | 0.81 | 88.1 | 3.2 | 143 |
| Paraxanthine | 220613 | 181.072 | 0.09 | 11.2 | 0.02 | 500 | 26744 | 96.6 | 0.64 | ND | 8.1 | 148 |
| Paraxanthine | 220623 | 181.072 | -0.08 | 11.2 | 0.01 | 500 | 24303 | 100 | 0.72 | ND | 3 | 38 |
| Paraxanthine | 220704 | 181.072 | -0.03 | 11.2 | 0.01 | 500 | 17694 | 86 | 0.71 | 107.9 | 5 | 143 |
| Paraxanthine | 220803 | 181.072 | -0.28 | 11.3 | 0.01 | 500 | 25476 | 100 | 0.74 | ND | 1.2 | 81 |
| Paraxanthine | 221021 | 181.072 | -0.20 | 11 | 0.01 | 500 | 21814 | 100 | 0.79 | 100.4 | 2.6 | 187 |
| Paraxanthine | 230109 | 181.072 | 0.13 | 10.8 | 0.01 | 500 | 20455 | 93.9 | 0.57 | 91 | 2.6 | 181 |
| Paraxanthine | 230424 | 181.072 | 0.09 | 10.8 | 0.01 | 500 | 20486 | 88.9 | 0.54 | 97.4 | 3.2 | 45 |
| Paraxanthine | 230711 | 181.072 | -0.26 | 11.1 | 0.01 | 500 | 24761 | 100 | 0.66 | 112.7 | 8.4 | 100 |
| Paraxanthine | 231106 | 181.072 | 0.86 | 11.1 | 0.03 | 500 | 18790 | 100 | 0.61 | ND | 1 | 45 |
| Paraxanthine | 240131 | 181.072 | 0.58 | 11.2 | 0.02 | 500 | 22208 | 100 | 0.78 | ND | 2.3 | 127 |
| Paraxanthine | 240701 | 181.072 | 0.43 | 11.2 | 0.03 | 500 | 20914 | 100 | 0.77 | ND | 1.2 | 143 |
| Pheniramine | 220613 | 241.1699 | 0.08 | 10.6 | 0.06 | 20 | 52 | 64.9 | 0.64 | 80.8 | 4.1 | 148 |
| Pheniramine | 220623 | 241.1699 | -0.14 | 10.6 | 0.05 | 20 | 61 | 50 | 0.78 | 90.6 | 2.6 | 38 |
| Pheniramine | 220704 | 241.1699 | 0.08 | 10.7 | 0.06 | 20 | 57 | 41.3 | 0.81 | 97.9 | 6.7 | 143 |
| Pheniramine | 220803 | 241.1699 | -0.22 | 10.7 | 0.05 | 20 | 39 | 84 | 0.8 | 101.7 | 10.6 | 81 |
| Pheniramine | 221021 | 241.1699 | 0.00 | 10.5 | 0.05 | 20 | 57 | 49.2 | 0.77 | 90.6 | ND | 187 |
| Pheniramine | 230109 | 241.1699 | 0.71 | 10.4 | 0.03 | 20 | 72 | 69.1 | 0.74 | 109.8 | 2.9 | 181 |
| Pheniramine | 230424 | 241.1699 | 0.52 | 10.4 | 0.02 | 20 | 86 | 97.8 | 0.9 | 103 | 6.8 | 45 |
| Pheniramine | 230711 | 241.1699 | 0.19 | 10.7 | 0.04 | 20 | 63 | 76 | 1.01 | 102.7 | 4.9 | 100 |
| Pheniramine | 231106 | 241.1699 | 2.02 | 10.4 | 0 | 20 | 52 | 60 | 0.76 | 90.7 | 15.4 | 45 |
| Pheniramine | 240131 | 241.1699 | 1.34 | 10.6 | 0.01 | 20 | 66 | 85 | 0.68 | 97.4 | 7.4 | 127 |
| Pheniramine | 240701 | 241.1699 | 1.07 | 11.5 | 0.02 | 20 | 53 | 73.4 | 0.85 | 88.3 | 5.5 | 143 |
| Sulfamethoxazole | 220613 | 254.0594 | 0.41 | 13 | 0.01 | 25 | 231 | 97.3 | 0.62 | 120.4 | 13.9 | 148 |
| Sulfamethoxazole | 220623 | 254.0594 | 0.30 | 13.1 | 0.01 | 25 | 128 | 84.2 | 0.7 | 105.2 | 5.7 | 38 |
| Sulfamethoxazole | 220704 | 254.0594 | 0.25 | 13.1 | 0.02 | 25 | 160 | 78.3 | 0.68 | 101.8 | 10.2 | 143 |

| Substance | Measurement batch ID | m/z theoretical [M+H]+l | m/z delta [ppm] | RT measured [min] | SD RT [min] | LOQ [ng/L] | Sample conc. [ng/L] | Samples above LOQ [%] | Absolute recovery | Relative recovery [%] | Precision [%] | Number of samples |
|---|---|---|---|---|---|---|---|---|---|---|---|---|
| Sulfamethoxazole | 220803 | 254.0594 | -0.03 | 13.1 | 0.01 | 25 | 84 | 88.6 | 0.68 | 98.4 | ND | 79 |
| Sulfamethoxazole | 221021 | 254.0594 | 0.30 | 12.9 | 0.01 | 25 | 100 | 81.5 | 0.71 | 103.3 | ND | 184 |
| Sulfamethoxazole | 230109 | 254.0594 | 0.40 | 12.6 | 0.02 | 25 | 244 | 94.5 | 0.58 | 106.7 | 16 | 181 |
| Sulfamethoxazole | 230424 | 254.0594 | 0.27 | 12.6 | 0.01 | 25 | 253 | 97.8 | 0.61 | 99.1 | 15.7 | 45 |
| Sulfamethoxazole | 230711 | 254.0594 | 0.05 | 12.9 | 0.01 | 25 | 275 | 97 | 0.6 | 104.4 | 8.8 | 99 |
| Sulfamethoxazole | 231106 | 254.0594 | 1.43 | 12.9 | 0.02 | 25 | 251 | 97.8 | 0.58 | 93.8 | 1.5 | 46 |
| Sulfamethoxazole | 240131 | 254.0594 | 0.99 | 12.9 | 0.02 | 25 | 312 | 93.7 | 0.66 | 105.4 | 4 | 127 |
| Sulfamethoxazole | 240701 | 254.0594 | 0.79 | 13 | 0.02 | 25 | 278 | 96.5 | 0.62 | 92.8 | 5 | 142 |
| Tramadol | 220613 | 264.1958 | 0.43 | 11.8 | 0.03 | 20 | 374 | 100 | 0.82 | 103.7 | 5.6 | 148 |
| Tramadol | 220623 | 264.1958 | 0.31 | 11.8 | 0.03 | 20 | 395 | 100 | 0.93 | 102.4 | 1.6 | 38 |
| Tramadol | 220704 | 264.1958 | 0.15 | 11.9 | 0.05 | 20 | 235 | 99.3 | 0.82 | 101.1 | 1.5 | 143 |
| Tramadol | 220803 | 264.1958 | -0.10 | 12 | 0.03 | 20 | 242 | 100 | 0.8 | 101.5 | ND | 79 |
| Tramadol | 221021 | 264.1958 | 0.20 | 11.7 | 0.04 | 20 | 184 | 100 | 0.83 | 99.8 | ND | 184 |
| Tramadol | 230109 | 264.1958 | 0.45 | 11.5 | 0.04 | 20 | 269 | 100 | 0.84 | 100.1 | 2.3 | 181 |
| Tramadol | 230424 | 264.1958 | 0.27 | 11.6 | 0.04 | 20 | 245 | 100 | 0.87 | 97.9 | 4.3 | 45 |
| Tramadol | 230711 | 264.1958 | 0.17 | 12 | 0.04 | 20 | 293 | 100 | 0.89 | 102 | 1 | 99 |
| Tramadol | 231106 | 264.1958 | 1.39 | 11.6 | 0.03 | 20 | 211 | 100 | 0.88 | 96.1 | 0.6 | 46 |
| Tramadol | 240131 | 264.1958 | 0.90 | 11.7 | 0.04 | 20 | 315 | 99.2 | 0.86 | 101 | 2.2 | 127 |
| Tramadol | 240701 | 264.1958 | 0.69 | 12 | 0.07 | 20 | 237 | 99.3 | 0.81 | 90.3 | 0.6 | 142 |
| Trimethoprim | 220613 | 291.1452 | 0.47 | 10.3 | 0.02 | 30 | 198 | 100 | 0.8 | 103 | 6.1 | 148 |
| Trimethoprim | 220623 | 291.1452 | 0.34 | 10.3 | 0.02 | 30 | 160 | 97.4 | 0.93 | 104.3 | 2.7 | 38 |
| Trimethoprim | 220704 | 291.1452 | 0.11 | 10.3 | 0.03 | 30 | 159 | 97.9 | 0.86 | 103.7 | 1.6 | 143 |
| Trimethoprim | 220803 | 291.1452 | 0.17 | 10.4 | 0.02 | 30 | 167 | 100 | 0.87 | 101 | ND | 79 |
| Trimethoprim | 221021 | 291.1452 | 0.32 | 10.1 | 0.02 | 30 | 137 | 98.4 | 0.89 | 98.3 | ND | 184 |
| Trimethoprim | 230109 | 291.1452 | 1.05 | 10 | 0.03 | 30 | 134 | 99.4 | 0.74 | 96.8 | 5 | 181 |
| Trimethoprim | 230424 | 291.1452 | 0.69 | 10 | 0.03 | 30 | 154 | 100 | 0.83 | 98.7 | 6.8 | 45 |
| Trimethoprim | 230711 | 291.1452 | 0.59 | 10.4 | 0.03 | 30 | 171 | 98 | 0.81 | 101.6 | 2 | 99 |
| Trimethoprim | 231106 | 291.1452 | 1.99 | 10 | 0.03 | 30 | 146 | 100 | 0.75 | 90.5 | 3.3 | 46 |
| Trimethoprim | 240131 | 291.1452 | 1.66 | 10.1 | 0.03 | 30 | 126 | 96.9 | 0.82 | 101.3 | 5.1 | 127 |
| Trimethoprim | 240701 | 291.1452 | 0.57 | 10.4 | 0.05 | 30 | 163 | 98.6 | 0.76 | 93.2 | 3.7 | 142 |

#### SI 2.2 **Spatiotemporal patterns of chemical and virus loads in wastewater**

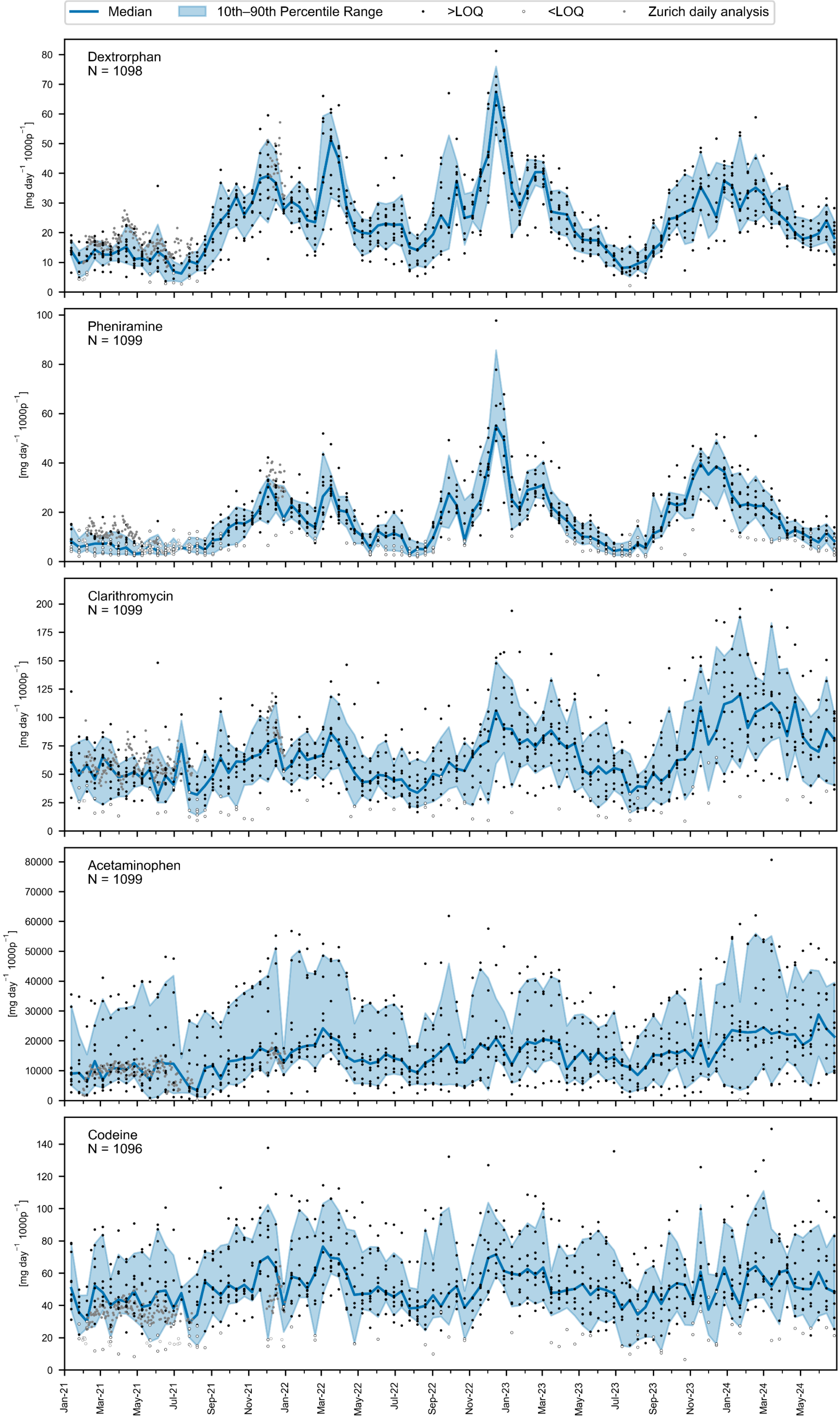


Figure SI 5: Spatiotemporal patterns of pharmaceutical loads in wastewater. Pharmaceutical loads from 10 wastewater treatment plants, sampled every 13th day from 2021 to June 2024, are represented by black dots. Grey dots indicate daily samples from Zurich, collected between February to July 2021 and in December 2021, highlighting day-to-day variability. Samples with values below the limit of quantification (LOQ) are depicted as white dots, with these loads estimated from concentration of 1/2 * LOQ. Day-specific medians are illustrated by interconnected blue dots, with shaded areas indicating the 10th to 90th interpercentile range.

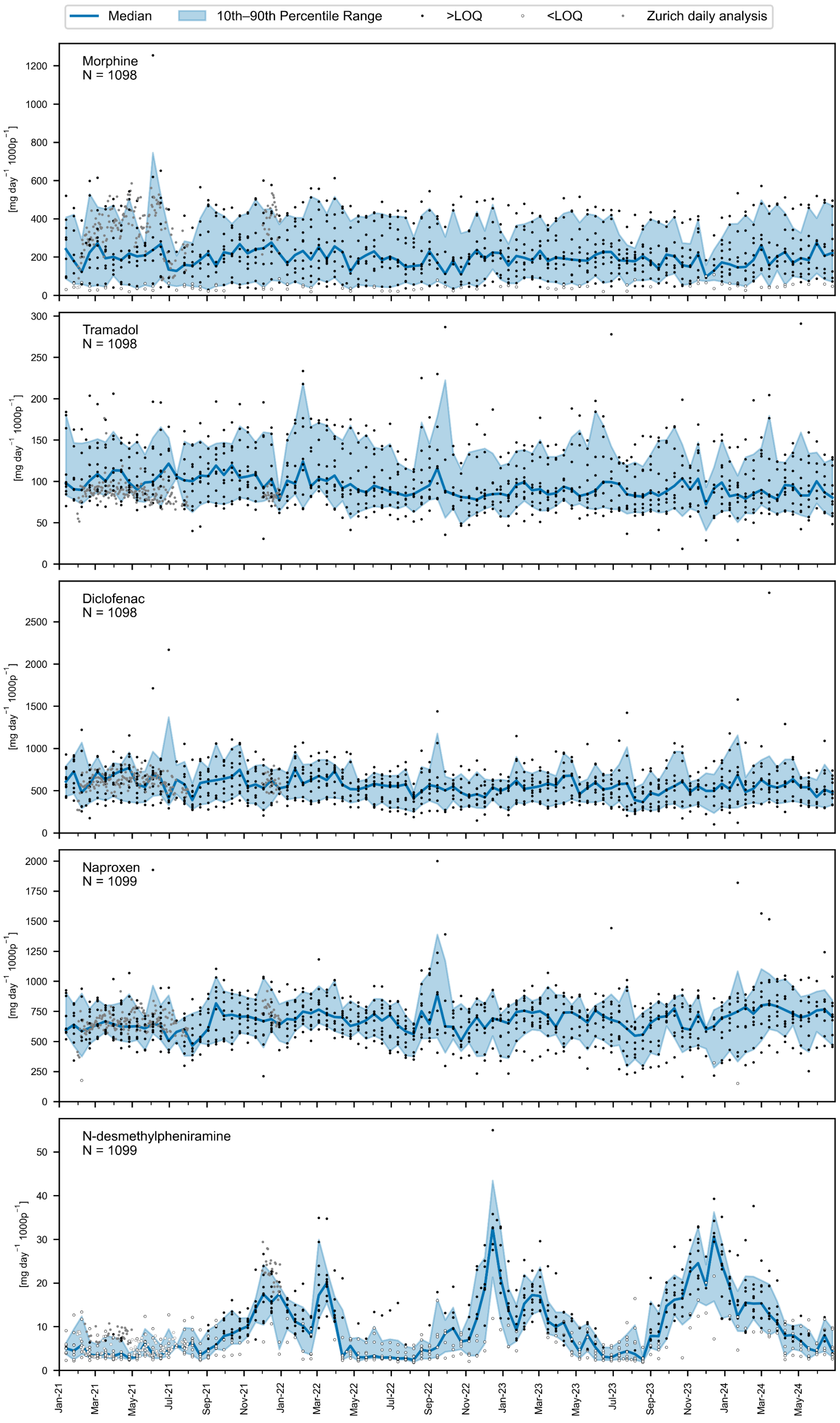


Figure SI 6: Spatiotemporal patterns of pharmaceutical loads in wastewater. Pharmaceutical loads from 10 wastewater treatment plants, sampled every 13th day from 2021 to June 2024, are represented by black dots. Grey dots indicate daily samples from Zurich, collected between February to July 2021 and in December 2021, highlighting day-to-day variability. Samples with values below the limit of quantification (LOQ) are depicted as white dots, with these loads estimated from concentration of 1/2 * LOQ. Day-specific medians are illustrated by interconnected blue dots, with shaded areas indicating the 10th to 90th interpercentile range.

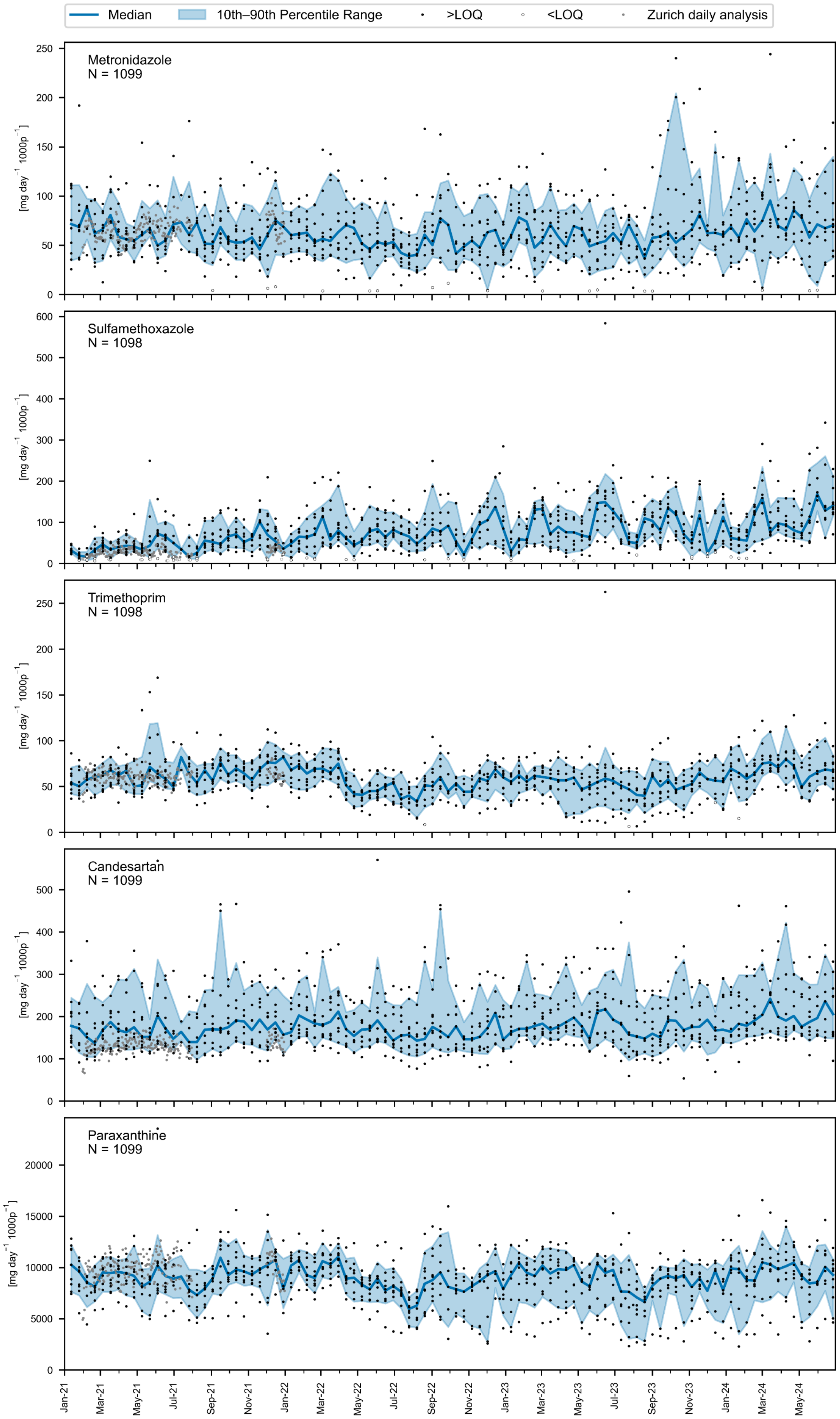


Figure SI 7: Spatiotemporal patterns of pharmaceutical loads in wastewater. Pharmaceutical loads from 10 wastewater treatment plants, sampled every 13th day from 2021 to June 2024, are represented by black dots. Grey dots indicate daily samples from Zurich, collected between February to July 2021 and in December 2021, highlighting day-to-day variability. Samples with values below the limit of quantification (LOQ) are depicted as white dots, with these loads estimated from concentration of 1/2 * LOQ. Day-specific medians are illustrated by interconnected blue dots, with shaded areas indicating the 10th to 90th interpercentile range.

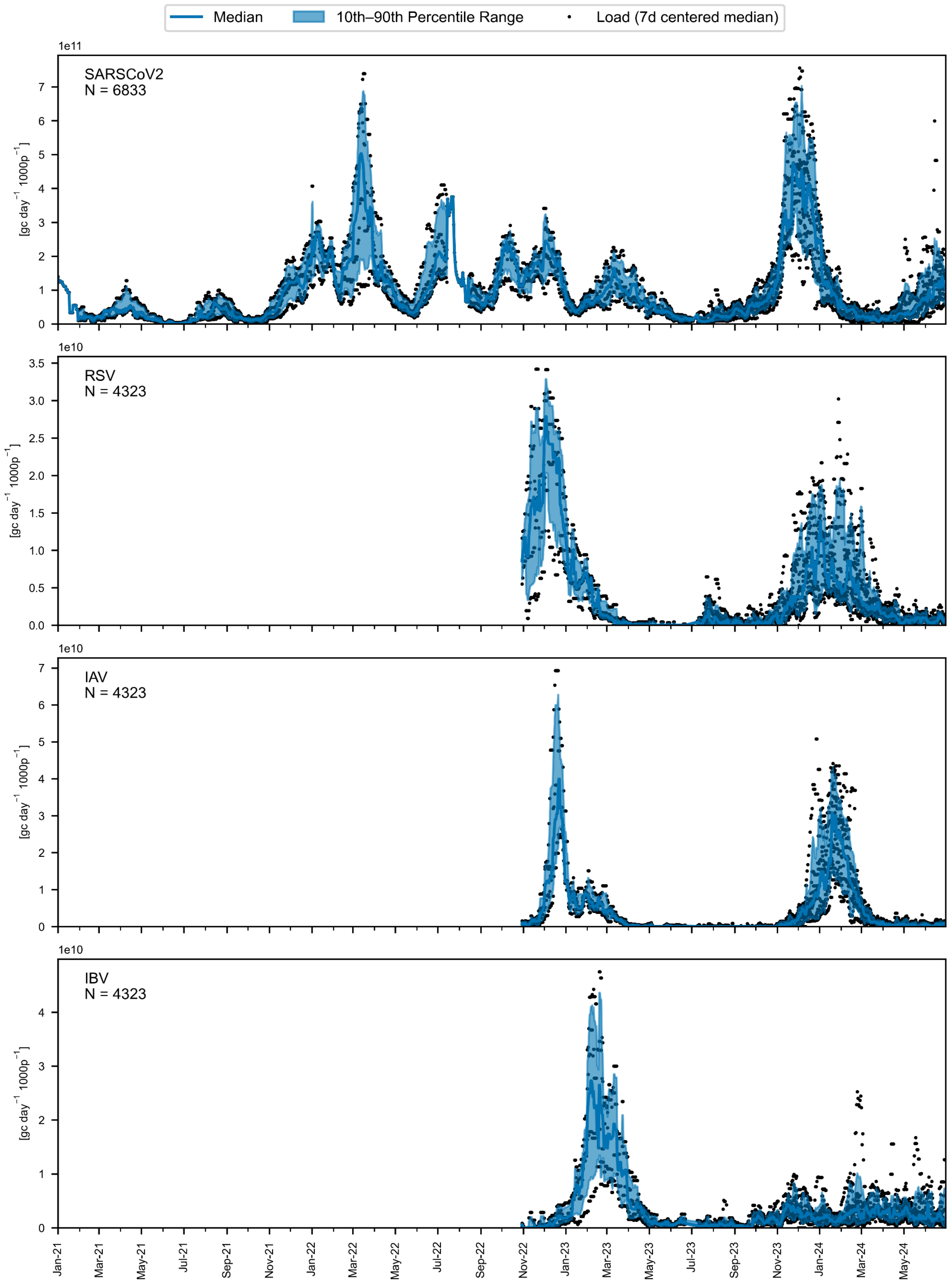


20

21 Figure SI 8: Spatiotemporal patterns of viral loads in wastewater. Day-specific gene copy median loads from SARS-CoV-2, RSV, influenza A (IAV) and influenza B (IBV)
22 are illustrated by interconnected blue dots. Shaded areas indicate the 10th to 90th interpercentile range.

23

#### SI 2.2.1 **Spatiotemporal pattern of baseline-subtracted loads for pharmaceuticals**

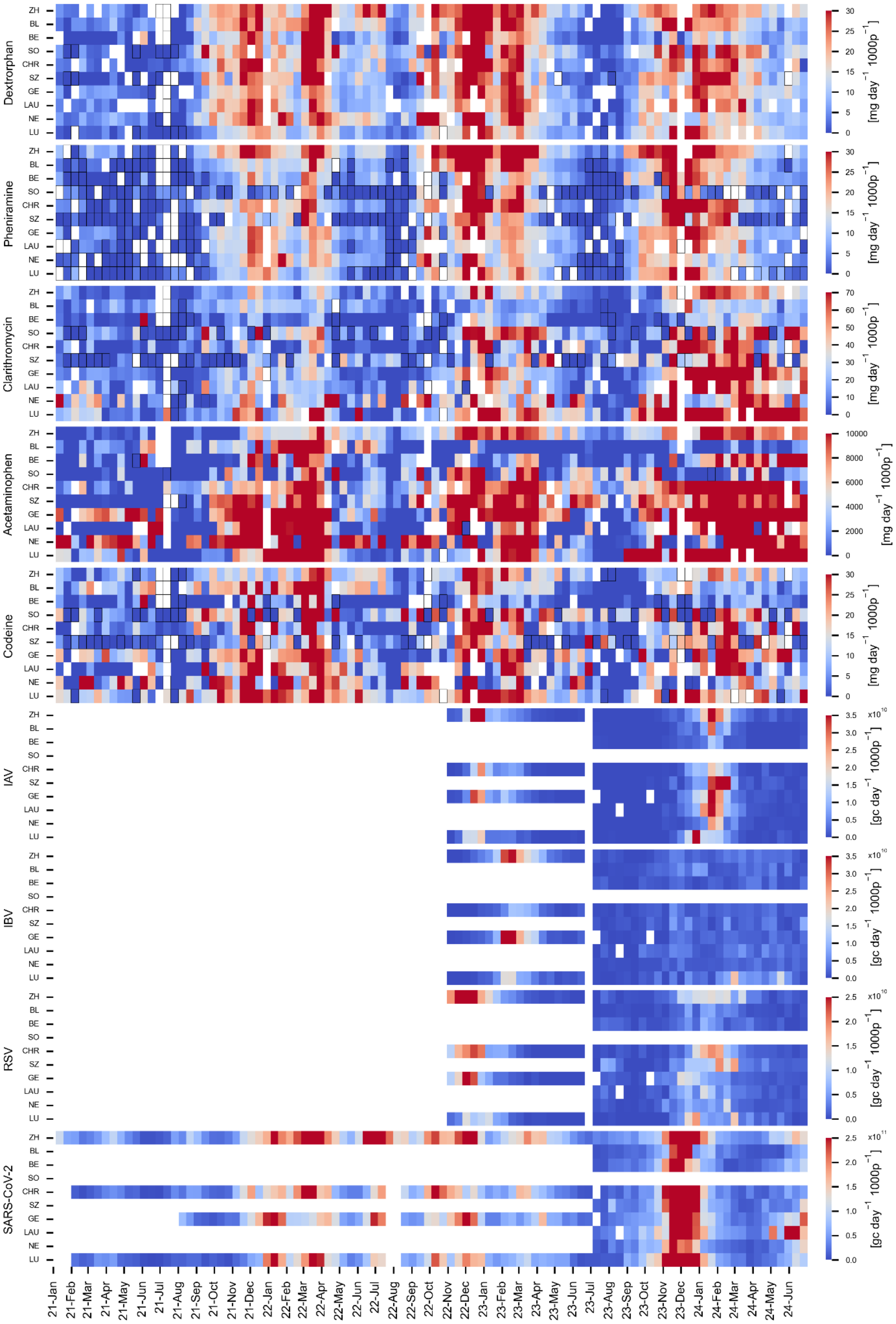


Figure SI 9: Spatiotemporal load patterns of baseline-subtracted pharmaceutical loads and viral gene copy loads in wastewater. Wastewater loads from every 13th day between January 2021 and June 2024 are displayed. Data is shown for individual treatment plants according to the abbreviations on the Y-axis: ZH (Zurich), BL (Basel), BE (Bern), SO (Solothurn), CHR (Chur), SZ (Schwyz), GE (Geneva), LA (Lausanne), NE (Neuchâtel), and LU (Lugano). These are arranged first by language region, then by catchment population size. The coloring represents the load values for pharmaceuticals [mg $d^{-1}$ $1000p^{-1}d^{-1}$] and viruses [gc $d^{-1}$ $1000p^{-1}$] with linear color scaling. For pharmaceuticals, each space refers to individual samples, and for the viral loads, each space reflects values from the centered 7-day median. White space indicates data exclusions due to day-specific wastewater volumes exceeding maximum threshold values or due to lack of sampling. Black-outlined spaces represent pharmaceutical loads from samples with concentrations below the limit of quantification (LOQ), which are estimated based on half the LOQ value. . Baseline loads were determined from location-specific mean values during periods of low respiratory virus exposure (June 2021 and July to August 2023).

##### SI 2.2.2 **Influenza subtype clinical surveillance data**

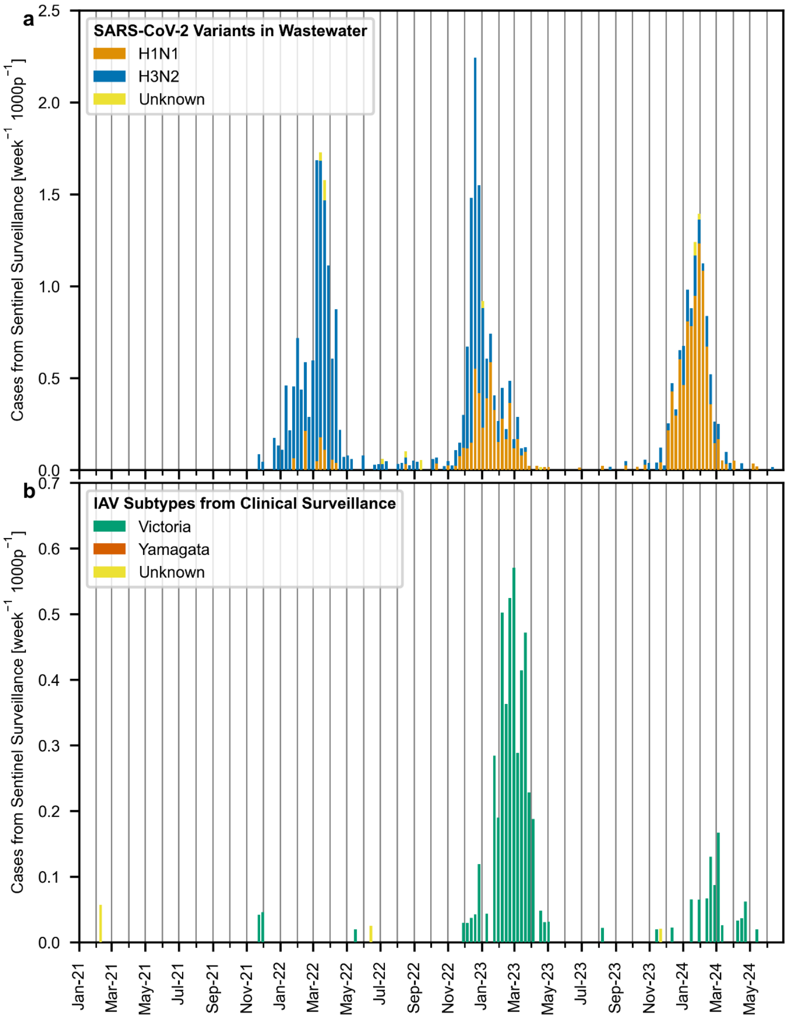


Figure SI 10: Temporal trends in national influenza subtypes from Swiss sentinel surveillance (Sentinella). (a) Case numbers of Influenza A subtypes and (b) case numbers of Influenza B subtypes.

##### SI 2.2.3 **Temporal trends of cases from mandatory reporting system and sentinel consultations**

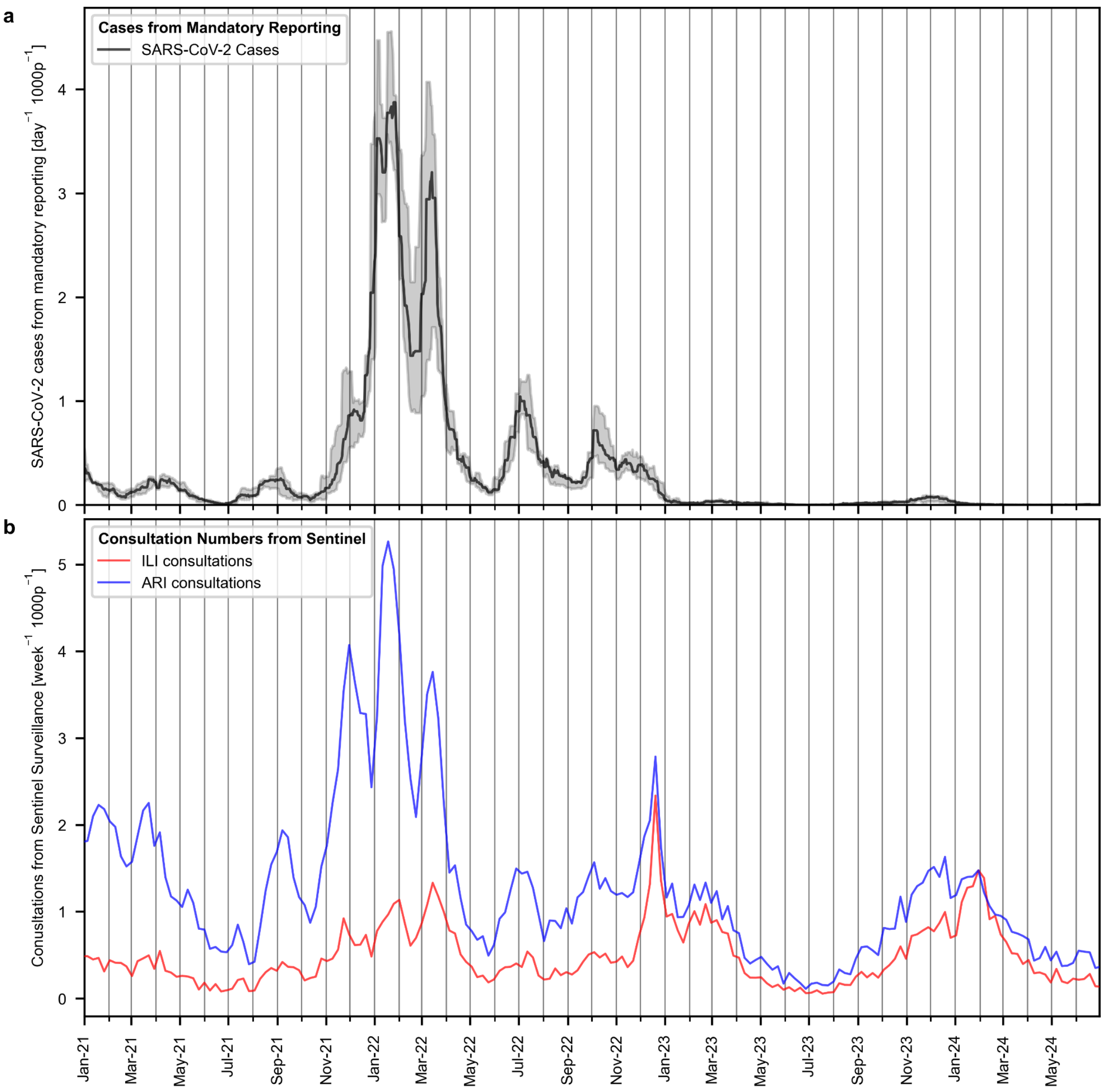


Figure SI 11: National level trends of SARS-COV-2 cases and sentinel consultations. (a) Median values of reported cases from the Swiss mandatory SARS-CoV-2 reporting system, based on the catchment areas of Zurich, Chur, Geneva, and Lugano, with the 10% to 90% interpercentile range represented by shaded areas. (b) General practitioner-based Sentinella data on weekly primary consultations for influenza-like illness (ILI) or acute respiratory infection (ARI) symptoms.

##### SI 2.2.4 **Distribution of respiratory viruses from sentinel clinical testing**

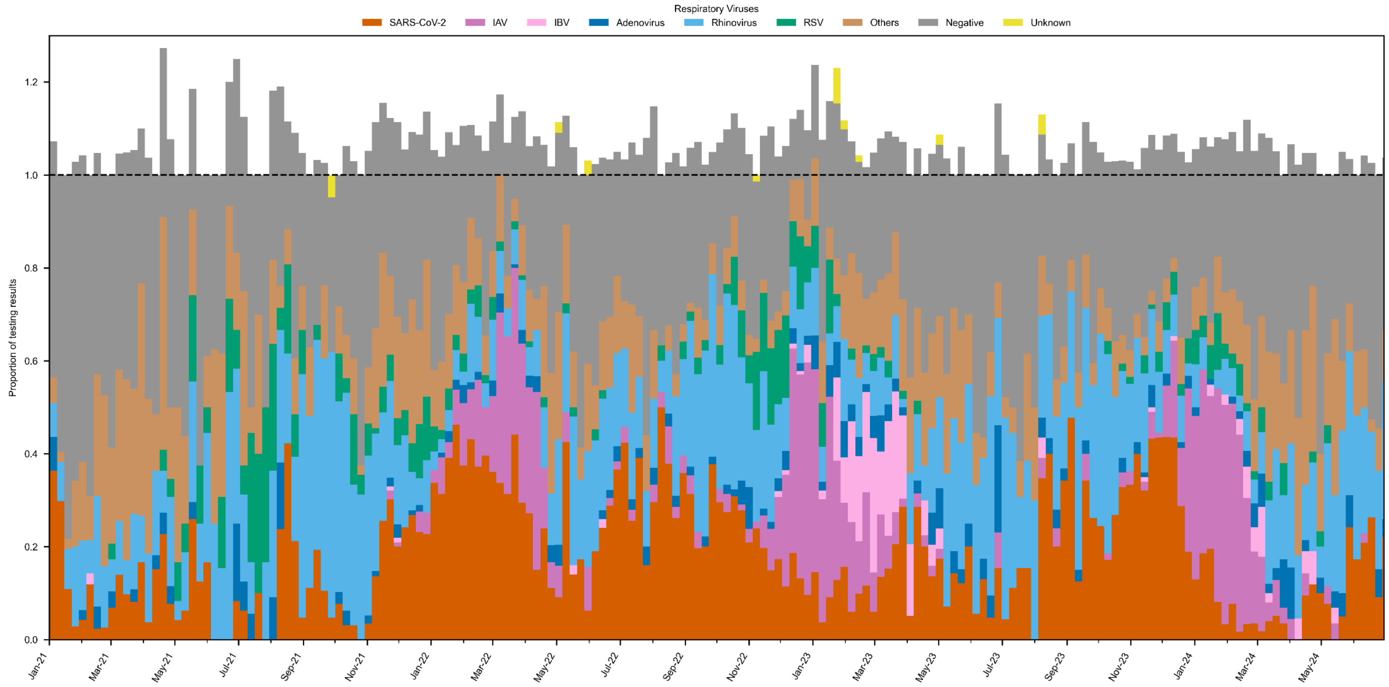


Figure SI 12: Proportion of positive tests for respiratory viruses from the Swiss sentinel surveillance system. This figure represents the proportion of positive test results for each virus, calculated by dividing the number of detections by the total number of samples tested. The "Others" category comprises detections of bocavirus, human coronaviruses (229E, HKU1, NL63, OC43), metapneumovirus, and parainfluenza viruses (types 1-4). The "Negative" category includes cases where individuals presented with symptoms of acute respiratory infection (ARI) or influenza-like illness (ILI) but tested negative for all surveillanced respiratory viruses. The "Unknown" category reflects samples with incomplete data regarding detected respiratory viruses.

The summed proportion may exceed 1 when samples test positive for multiple respiratory viruses. To account for symptom burden, we assume that co-infections are associated with more severe symptoms. Therefore, we calculate the proportion based on the total number of samples rather than the total number of detections.

#### SI 2.3 **Correlation analysis of wastewater markers**

##### SI 2.3.1 **Pearson correlations chemical and virus loads (location-specific)**

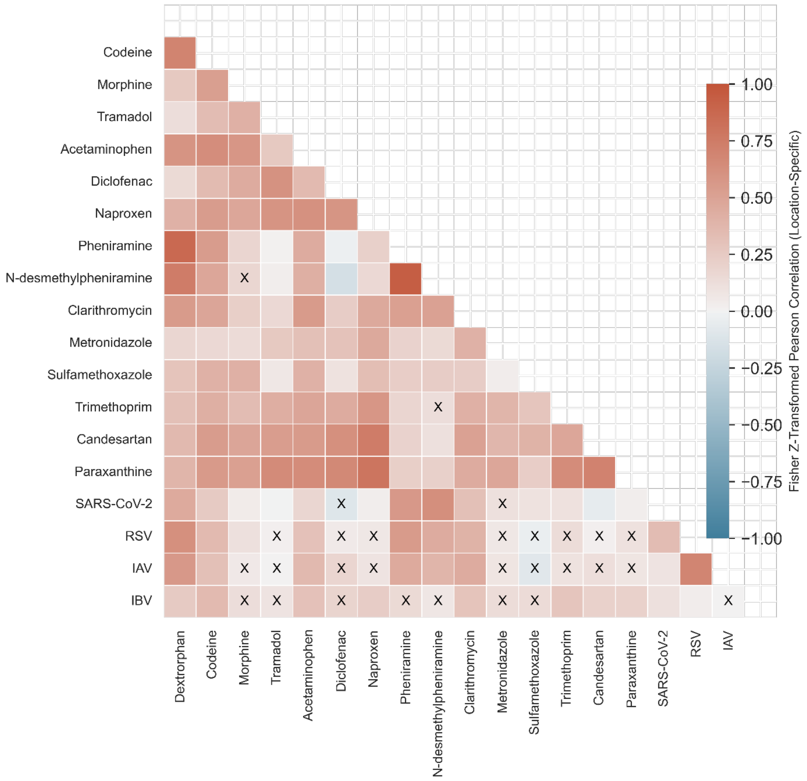


Figure SI 13: Fisher Z-transformed location-specific Pearson correlations of wastewater population loads. Correlations between pairs of wastewater markers were calculated for each of the ten locations and then aggregated using Fisher's Z-transformation. For chemical markers, loads from individual samples collected every 13th day were used, while for viruses, the 7-day centered median of daily values was employed. Correlation values with non-significant p-values (defined here as above 0.01) are marked with a cross. All wastewater pharmaceutical data with concentrations below the limit of quantification (LOQ) were excluded from the analysis.

#### SI 2.3.2 **Pearson correlations chemical and virus loads (location independent)**

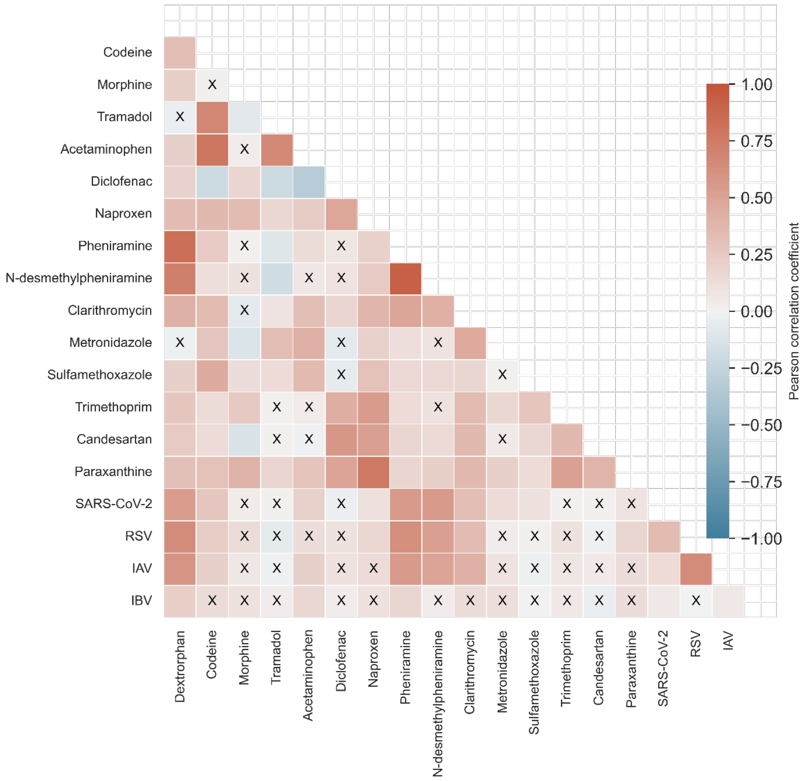


Figure SI 14: Pearson correlations of wastewater population loads. Correlations between pairs of wastewater markers were calculated across all data, independent of location. For pharmaceuticals, loads from individual samples collected every 13th day were used, while for viruses, the 7-day centered median of daily values was employed. Correlation values with p-values above 0.01 are marked with a cross. All wastewater pharmaceutical data with concentrations below the limit of quantification (LOQ) were excluded from the analysis.

#### SI 2.3.3 **Pearson correlations baseline-subtracted chemical and virus loads (location independent)**

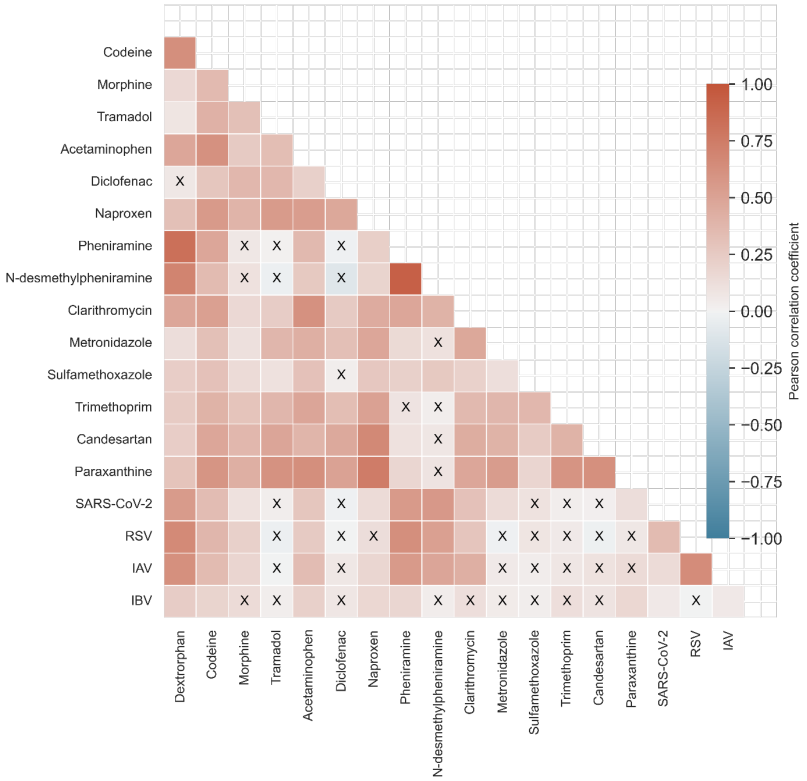


Figure SI 15: Pearson correlations of wastewater population loads. Correlations between pairs of wastewater markers were calculated across all data, independent of location. For pharmaceuticals, baseline-subtracted loads from individual samples collected every 13th day were used, while for viruses, the 7-day centered median of daily values was employed. Correlation values with p-values above 0.01 are marked with a cross. All wastewater pharmaceutical data with concentrations below the limit of quantification (LOQ) were excluded from the analysis. Baseline loads were determined from location-specific mean values during periods of low respiratory virus exposure (June 2021 and July to August 2023).

SI 2.3.4 **Scatter plot of viral and pharmaceutical loads (location independent)**

**Linear correlation of SARS-CoV-2 with pharmaceutical loads in wastewater**

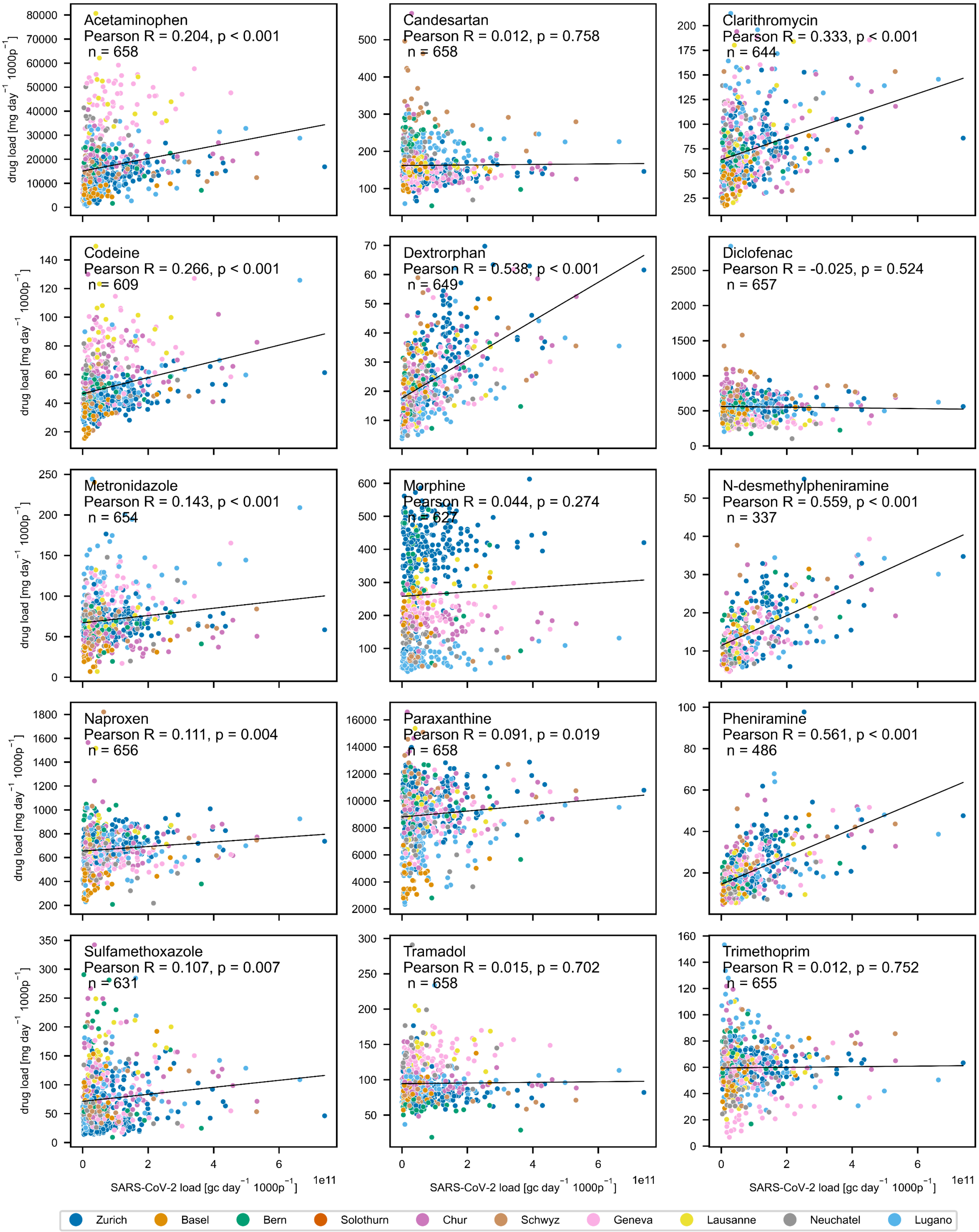


Figure SI 16: Scatterplot showing the correlation between SARS-COV-2 gene copy loads and chemical marker loads in wastewater, with data points colored by location. Pearson correlation coefficients (r), p-values, and the number of observations (n) are indicated for each marker.

#### Correlation of RSV with Pharmaceutical Loads in Wastewater

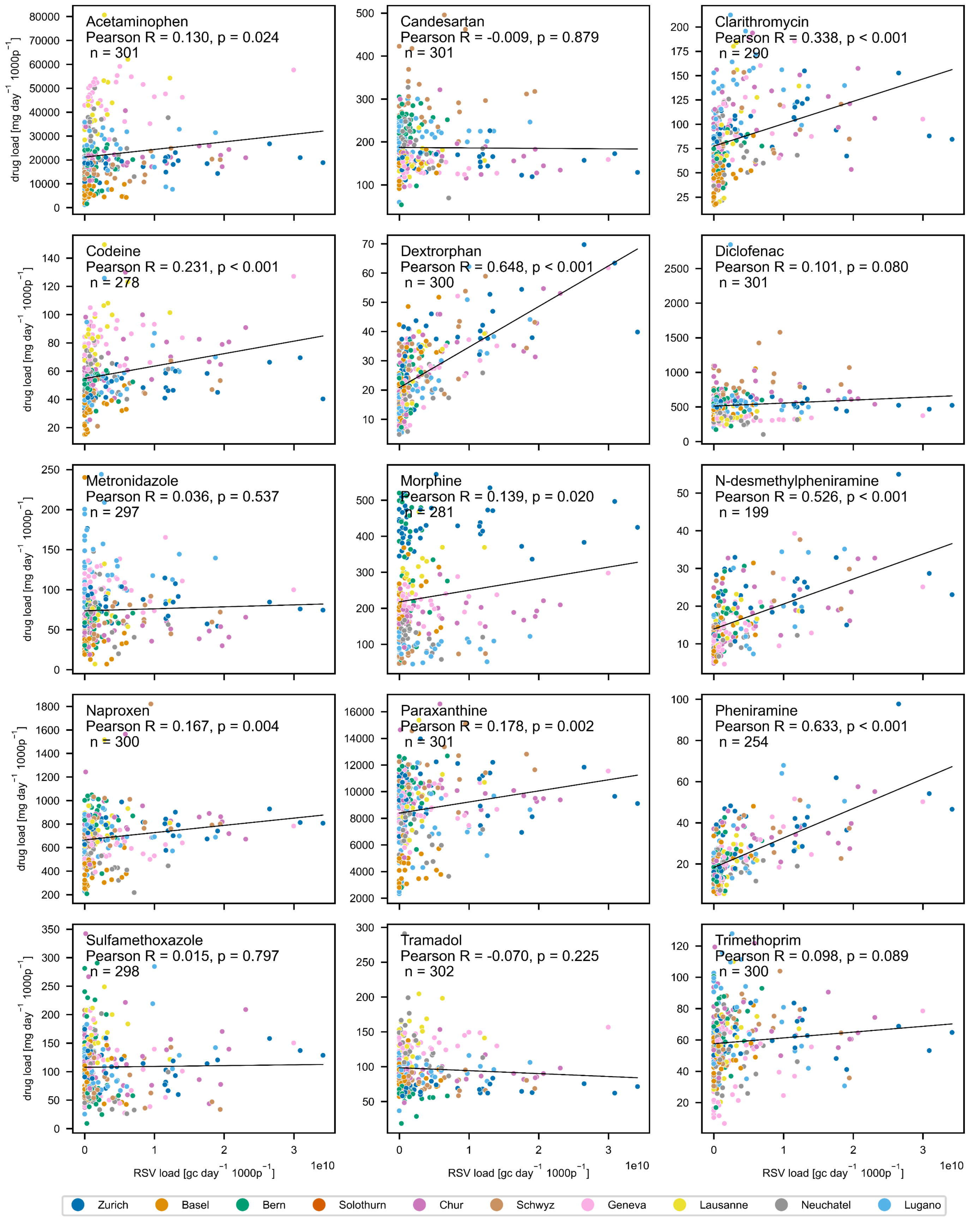


Figure SI 17: Scatterplot showing the correlation between RSV gene copy loads and chemical marker loads in wastewater, with data points colored by location. Pearson correlation coefficients (R), p-values, and the number of observations (n) are indicated for each marker.

**Correlation of Influenza A with Pharmaceutical Loads in Wastewater**

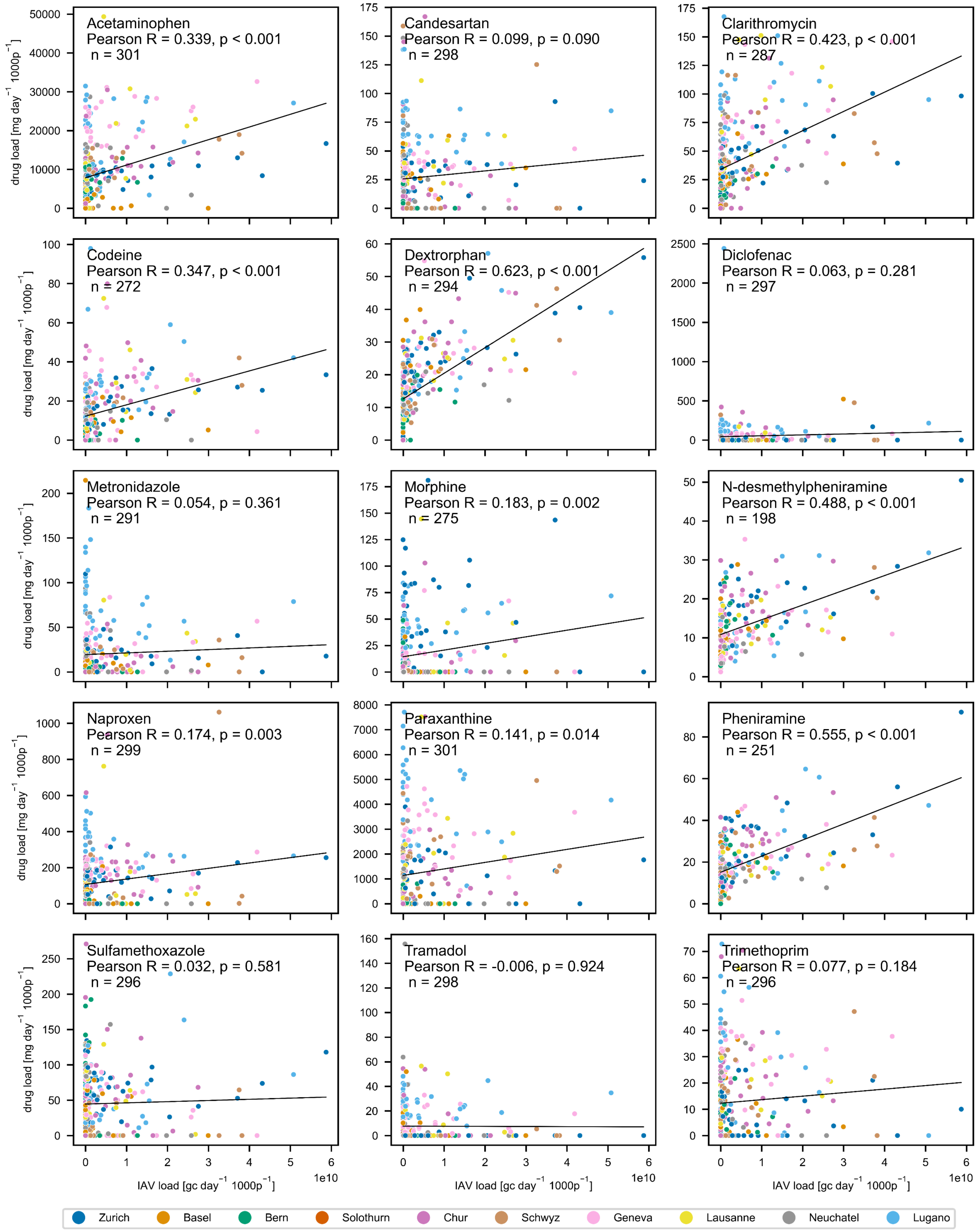


Figure SI 18: Scatterplot showing the correlation between Influenza A gene copy loads and chemical marker loads in wastewater, with data points colored by location. Pearson correlation coefficients (R), p-values, and the number of observations (n) are indicated for each marker.

**Correlation of Influenza B with Pharmaceutical Loads in Wastewater**

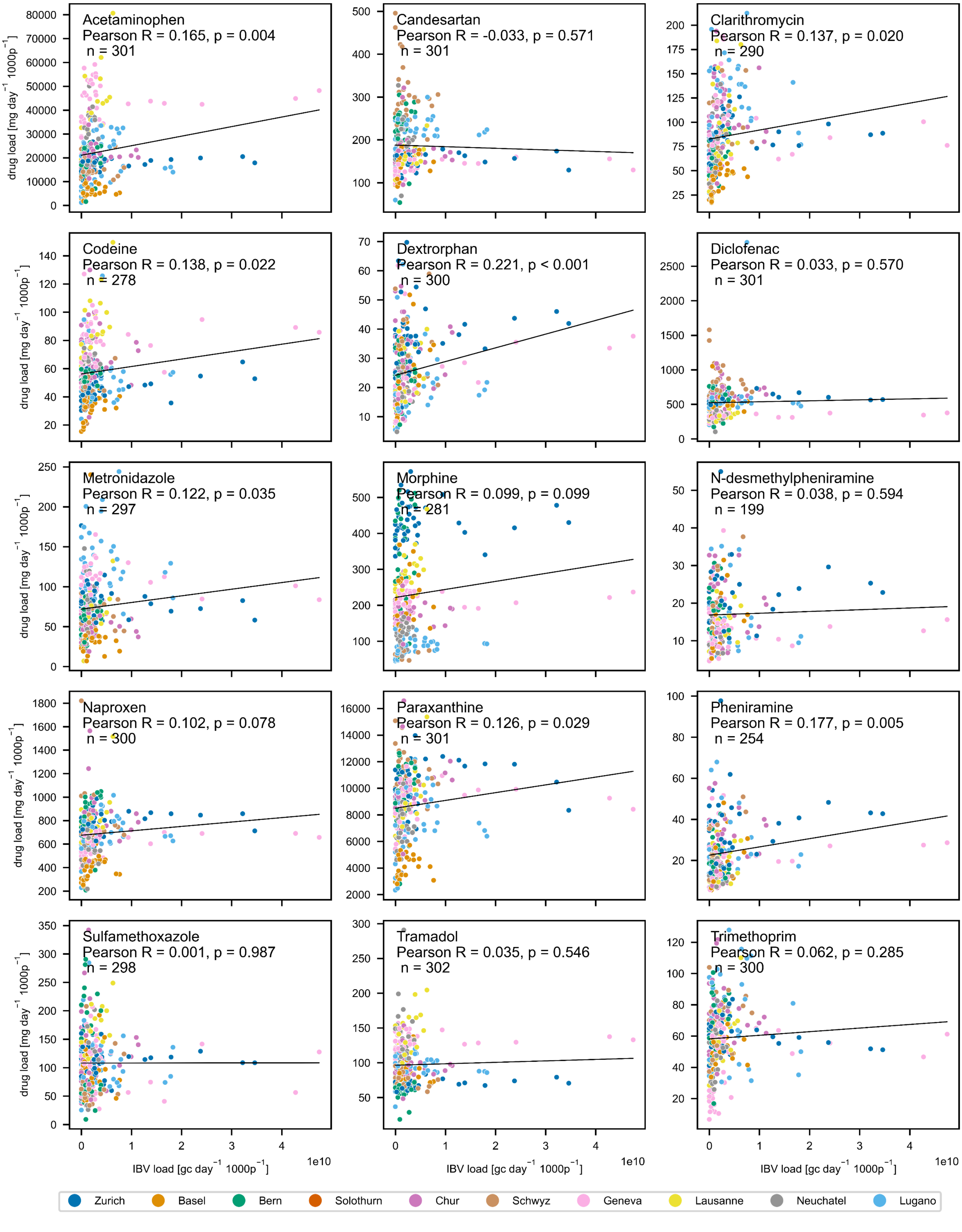


Figure SI 19: Scatterplot showing the correlation between Influenza B gene copy loads and chemical marker loads in wastewater, with data points colored by location. Pearson correlation coefficients (R), p-values, and the number of observations (n) are indicated for each marker.

#### SI 2.4 **Regression analyses of pharmaceutical loads in wastewater and viral exposure**

##### SI 2.4.1 **Ordinary least squares (OLS) analyses of pharmaceutical and viral loads in wastewater**

**OLS diagnostics**

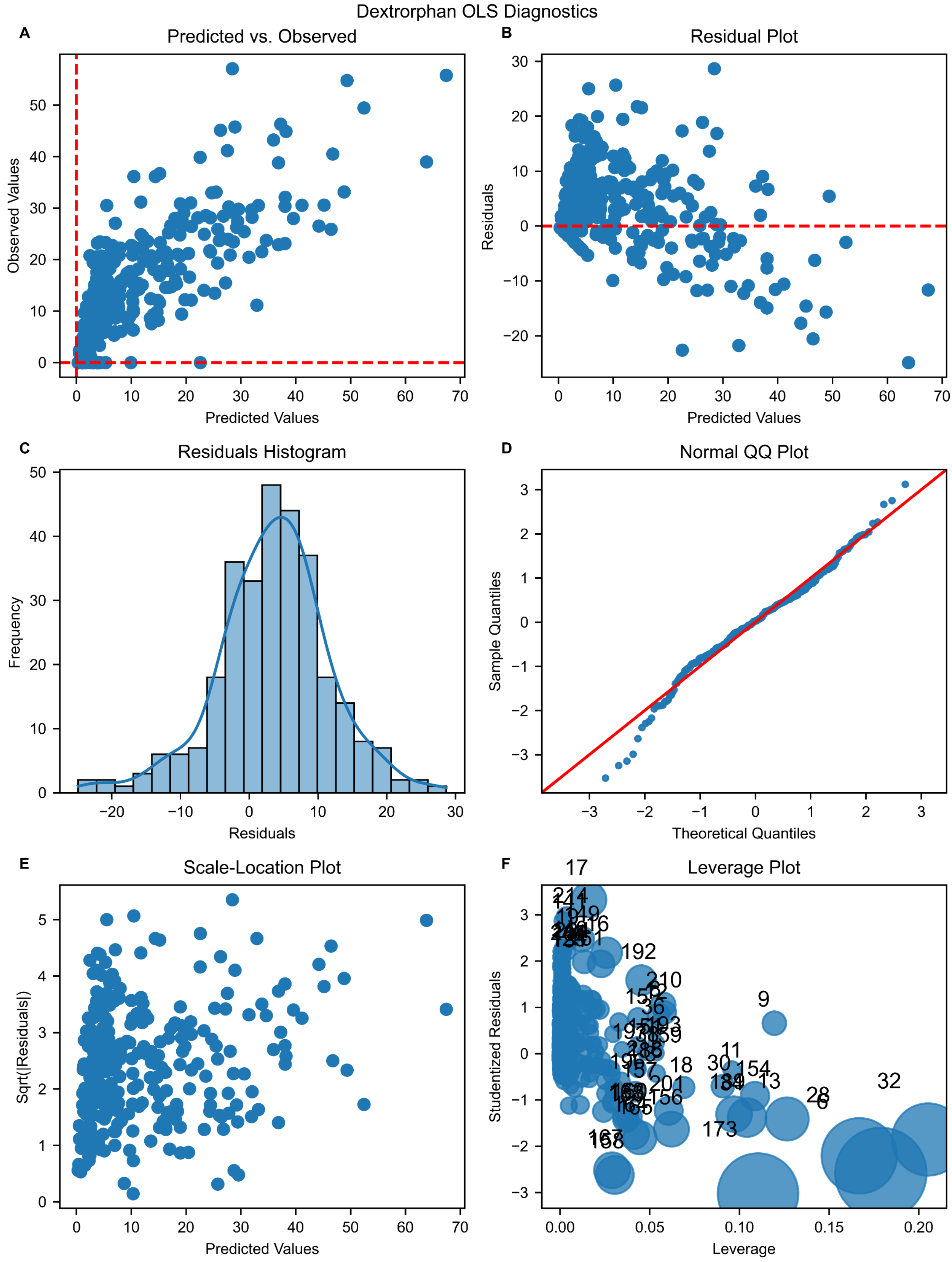


Figure SI 20: Diagnostic plots for the Ordinary Least Squares (OLS) regression analysis using 7-day centered median gene copy number loads of SARS-CoV-2, RSV, IAV, and IBV as predictors, with baseline-subtracted pharmaceutical loads as the response variable. The figure includes: (A) Predicted vs. Observed Values, depicting the correlation between predicted and observed pharmaceutical loads; (B) Residual Plot, illustrating residuals against predicted values to assess homoscedasticity; (C) Residuals Histogram, showing the residual distribution with an overlaid kernel density estimate; (D) Normal Q-Q Plot, evaluating the normality of residuals; (E) Scale-Location Plot, revealing the spread of residuals to detect non-constant variance; and (F) Leverage Plot, highlighting influential data points using Cook's distance.

Table SI 5: Ordinary Least Squares (OLS) regression results using gene copy loads of SARS-COV-2, RSV, IAV and IBV as input as predicting variable and baseline-subtracted dextrorphan loads as the response variable.

```
                                 OLS Regression Results
=======================================================================================
Dep. Variable:            Dextrorphan   R-squared (uncentered):                   0.796
Model:                            OLS   Adj. R-squared (uncentered):              0.793
Method:                 Least Squares   F-statistic:                              283.8
Date:                Sat, 12 Oct 2024   Prob (F-statistic):                    4.31e-99
Time:                        20:05:14   Log-Likelihood:                         -1059.5
No. Observations:                 295   AIC:                                      2127.
Df Residuals:                     291   BIC:                                      2142.
Df Model:                           4
Covariance Type:            nonrobust
==============================================================================
                 coef    std err          t      P>|t|      [0.025      0.975]
------------------------------------------------------------------------------
SARSCoV2    4.716e-11   4.73e-12      9.971      0.000    3.79e-11    5.65e-11
RSV         1.016e-09   1.22e-10      8.303      0.000    7.75e-10    1.26e-09
IAV         4.555e-10   7.47e-11      6.098      0.000    3.09e-10    6.03e-10
IBV         7.838e-10   8.71e-11      8.997      0.000    6.12e-10    9.55e-10
==============================================================================
Omnibus:                       13.243   Durbin-Watson:                   1.083
Prob(Omnibus):                  0.001   Jarque-Bera (JB):               20.882
Skew:                          -0.291   Prob(JB):                     2.92e-05
Kurtosis:                       4.166   Cond. No.                         35.9
==============================================================================

Notes:
[1] R² is computed without centering (uncentered) since the model does not contain a constant.
[2] Standard Errors assume that the covariance matrix of the errors is correctly specified.
```

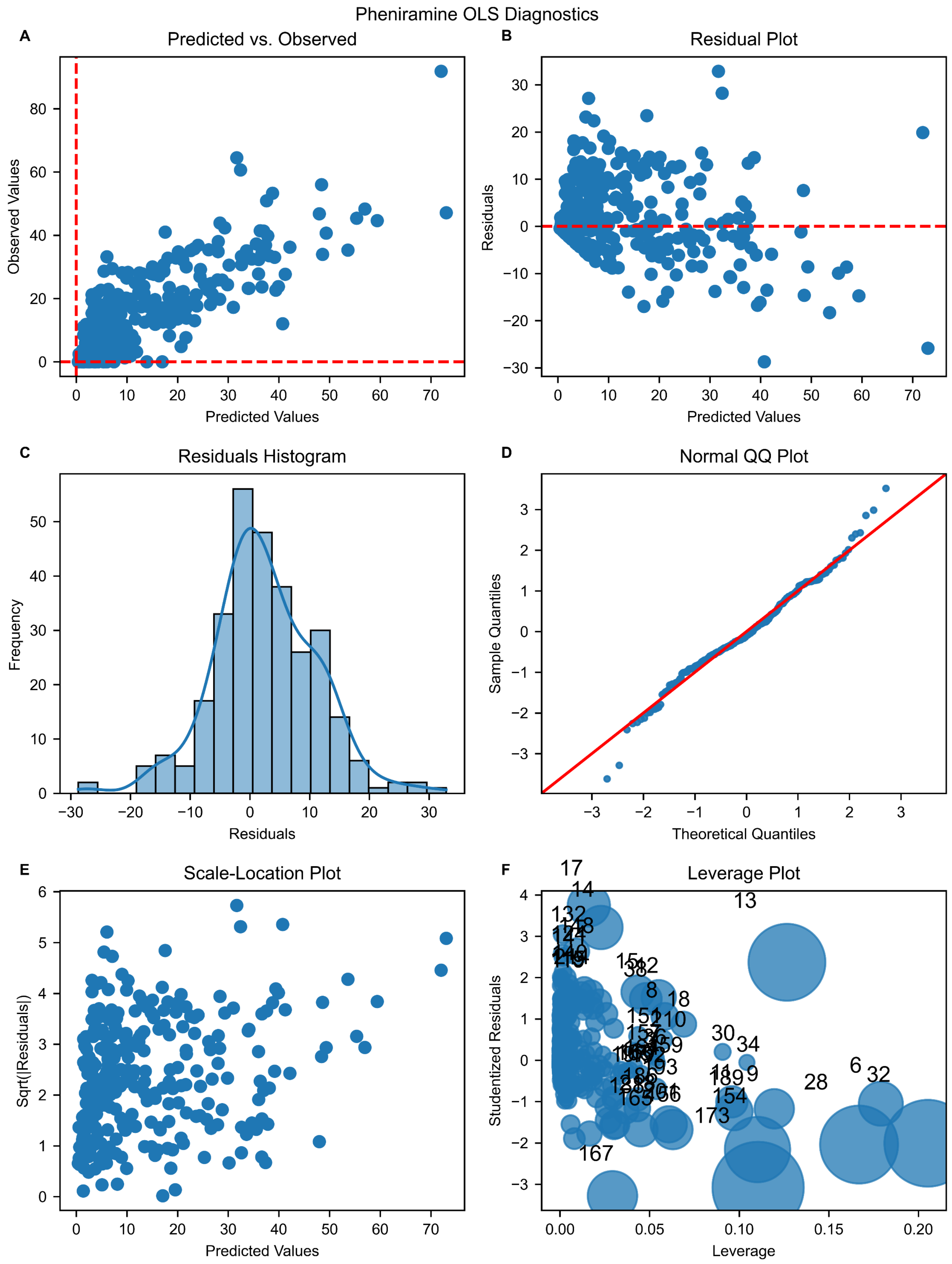


Figure SI 21: Diagnostic plots for the Ordinary Least Squares (OLS) regression analysis using 7-day centered median gene copy number loads of SARS-CoV-2, RSV, IAV, and IBV as predictors, with baseline-subtracted pharmaceutical loads as the response variable. The figure includes: (A) Predicted vs. Observed Values, depicting the correlation between predicted and observed pharmaceutical loads; (B) Residual Plot, illustrating residuals against predicted values to assess homoscedasticity; (C) Residuals Histogram, showing the residual distribution with an overlaid kernel density estimate; (D) Normal Q-Q Plot, evaluating the normality of residuals; (E) Scale-Location Plot, revealing the spread of residuals to detect non-constant variance; and (F) Leverage Plot, highlighting influential data points using Cook's distance.

Table SI 6: Ordinary Least Squares (OLS) regression results using gene copy loads of SARS-COV-2, RSV, IAV and IBV as input as predicting variable and baseline-subtracted pheniramine loads as the response variable.

```
                                 OLS Regression Results
=======================================================================================
Dep. Variable:            Pheniramine   R-squared (uncentered):                   0.823
Model:                            OLS   Adj. R-squared (uncentered):              0.821
Method:                 Least Squares   F-statistic:                              336.7
Date:                Sat, 12 Oct 2024   Prob (F-statistic):                   1.94e-107
Time:                        20:05:27   Log-Likelihood:                         -1058.8
No. Observations:                 293   AIC:                                      2126.
Df Residuals:                     289   BIC:                                      2140.
Df Model:                           4
Covariance Type:            nonrobust
==============================================================================
                 coef    std err          t      P>|t|      [0.025      0.975]
------------------------------------------------------------------------------
SARSCoV2     7.19e-11   4.87e-12     14.773      0.000    6.23e-11    8.15e-11
RSV         9.345e-10   1.25e-10      7.461      0.000    6.88e-10    1.18e-09
IAV          4.69e-10   7.64e-11      6.141      0.000    3.19e-10    6.19e-10
IBV         6.366e-10   8.91e-11      7.148      0.000    4.61e-10    8.12e-10
==============================================================================
Omnibus:                        7.410   Durbin-Watson:                   1.086
Prob(Omnibus):                  0.025   Jarque-Bera (JB):               12.107
Skew:                           0.052   Prob(JB):                      0.00235
Kurtosis:                       3.991   Cond. No.                         35.8
==============================================================================

Notes:
[1] R² is computed without centering (uncentered) since the model does not contain a constant.
[2] Standard Errors assume that the covariance matrix of the errors is correctly specified.
```

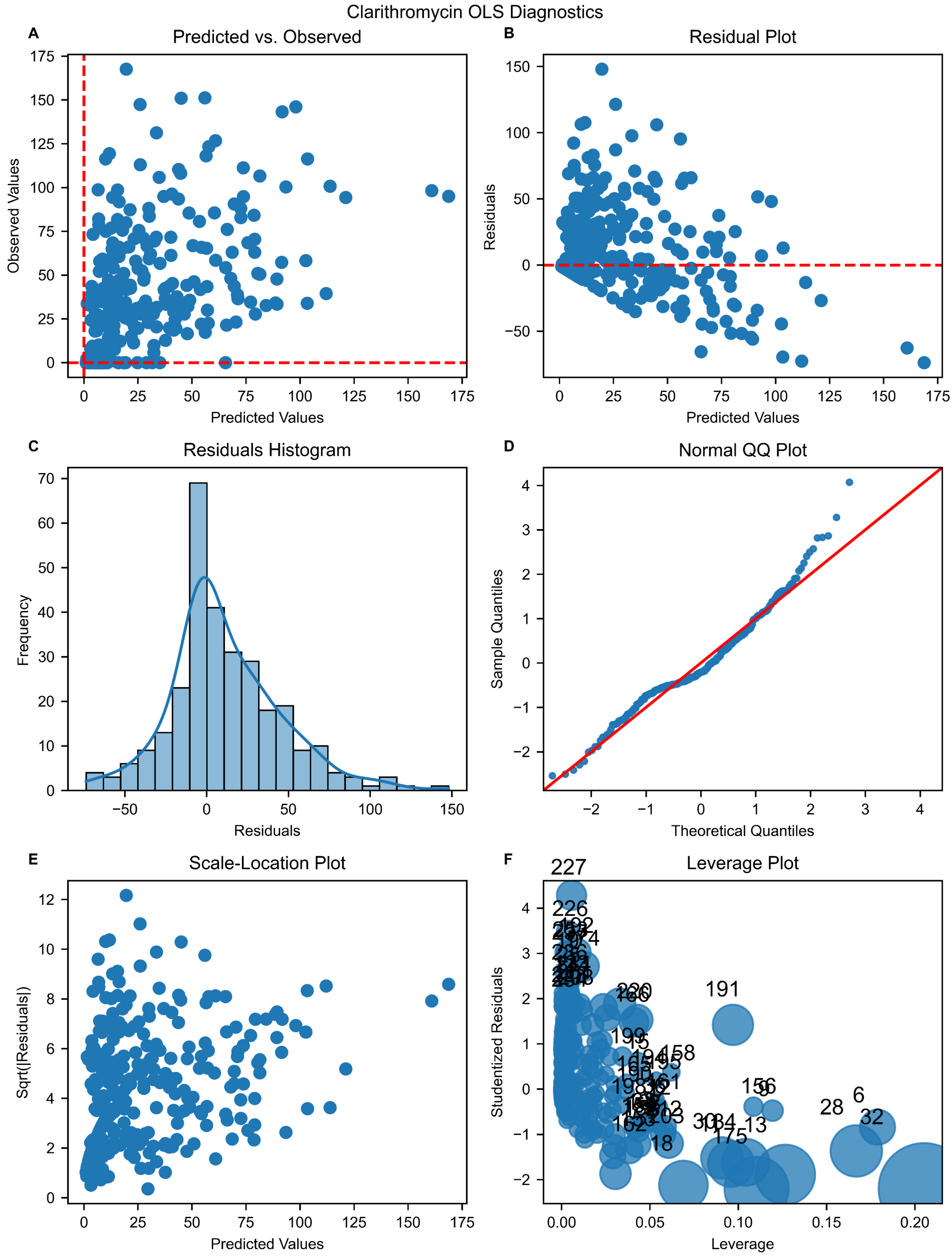


Figure SI 22: Diagnostic plots for the Ordinary Least Squares (OLS) regression analysis using 7-day centered median gene copy number loads of SARS-CoV-2, RSV, IAV, and IBV as predictors, with baseline-subtracted pharmaceutical loads as the response variable. The figure includes: (A) Predicted vs. Observed Values, depicting the correlation between predicted and observed pharmaceutical loads; (B) Residual Plot, illustrating residuals against predicted values to assess homoscedasticity; (C) Residuals Histogram, showing the residual distribution with an overlaid kernel density estimate; (D) Normal Q-Q Plot, evaluating the normality of residuals; (E) Scale-Location Plot, revealing the spread of residuals to detect non-constant variance; and (F) Leverage Plot, highlighting influential data points using Cook's distance.

Table SI 7: Ordinary Least Squares (OLS) regression results using gene copy loads of SARS-COV-2, RSV, IAV and IBV as input as predicting variable and baseline-subtracted clarithromycine loads as the response variable.

```
                                 OLS Regression Results
=======================================================================================
Dep. Variable:          Clarithromycin   R-squared (uncentered):                   0.567
Model:                             OLS   Adj. R-squared (uncentered):              0.561
Method:                  Least Squares   F-statistic:                              95.75
Date:                 Sat, 12 Oct 2024   Prob (F-statistic):                    5.43e-52
Time:                         20:05:41   Log-Likelihood:                         -1481.0
No. Observations:                  297   AIC:                                      2970.
Df Residuals:                      293   BIC:                                      2985.
Df Model:                            4
Covariance Type:             nonrobust
==============================================================================
                 coef    std err          t      P>|t|      [0.025      0.975]
------------------------------------------------------------------------------
SARSCoV2    1.557e-10   1.91e-11      8.159      0.000    1.18e-10    1.93e-10
RSV         5.257e-10   4.94e-10      1.065      0.288   -4.46e-10     1.5e-09
IAV         1.766e-09   3.01e-10      5.861      0.000    1.17e-09    2.36e-09
IBV         1.647e-09   3.51e-10      4.687      0.000    9.56e-10    2.34e-09
==============================================================================
Omnibus:                       27.514   Durbin-Watson:                   1.152
Prob(Omnibus):                  0.000   Jarque-Bera (JB):               38.983
Skew:                           0.629   Prob(JB):                     3.43e-09
Kurtosis:                       4.252   Cond. No.                         35.9
==============================================================================

Notes:
[1] R² is computed without centering (uncentered) since the model does not contain a constant.
[2] Standard Errors assume that the covariance matrix of the errors is correctly specified.
```

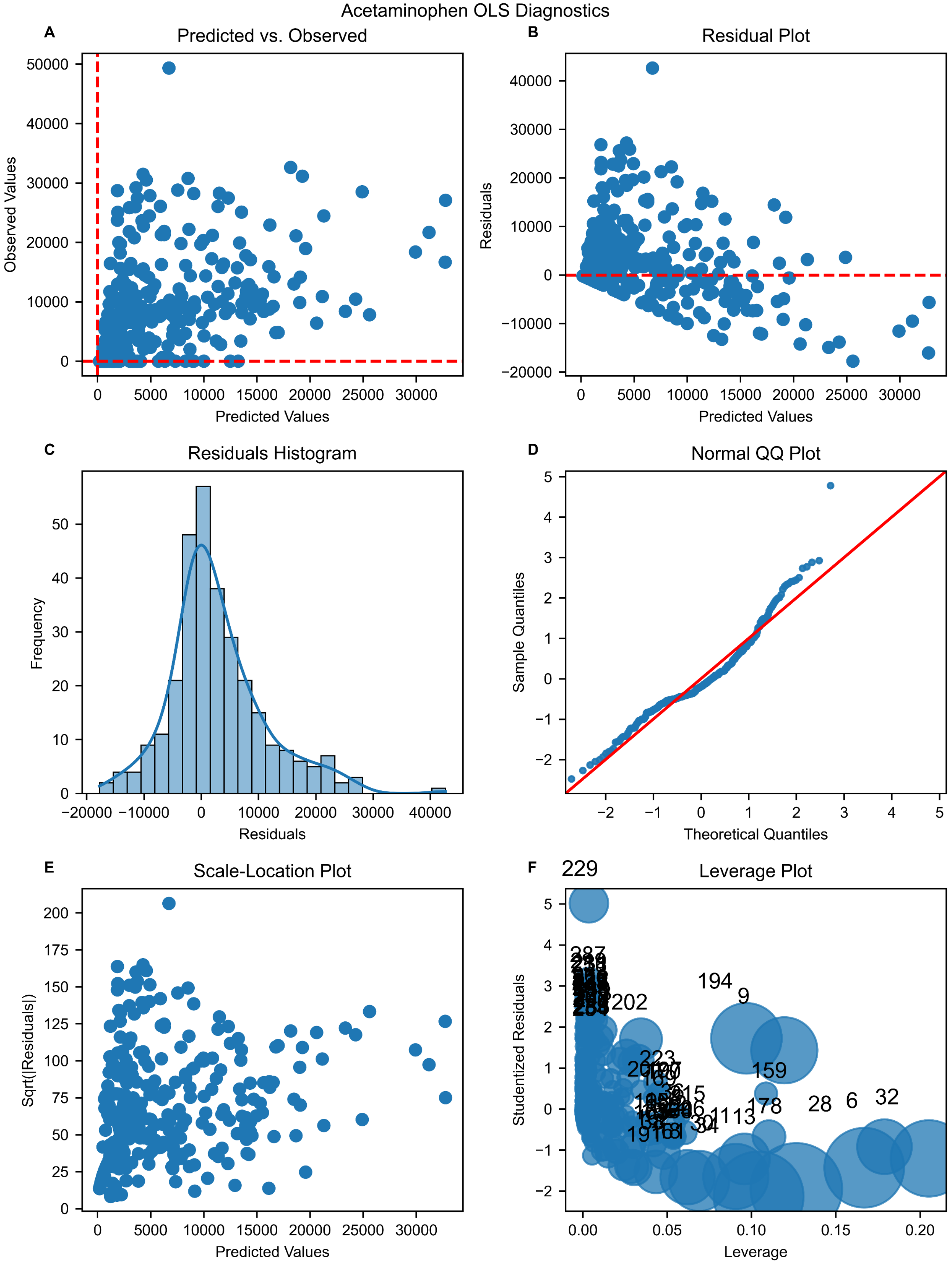


Figure SI 23: Diagnostic plots for the Ordinary Least Squares (OLS) regression analysis using 7-day centered median gene copy number loads of SARS-CoV-2, RSV, IAV, and IBV as predictors, with baseline-subtracted pharmaceutical loads as the response variable. The figure includes: (A) Predicted vs. Observed Values, depicting the correlation between predicted and observed pharmaceutical loads; (B) Residual Plot, illustrating residuals against predicted values to assess homoscedasticity; (C) Residuals Histogram, showing the residual distribution with an overlaid kernel density estimate; (D) Normal Q-Q Plot, evaluating the normality of residuals; (E) Scale-Location Plot, revealing the spread of residuals to detect non-constant variance; and (F) Leverage Plot, highlighting influential data points using Cook's distance.

Table SI 8: Ordinary Least Squares (OLS) regression results using gene copy loads of SARS-COV-2, RSV, IAV and IBV as input as predicting variable and baseline-subtracted acetaminophen loads as the response variable.

```
                                OLS Regression Results
=======================================================================================
Dep. Variable:          Acetaminophen   R-squared (uncentered):                  0.507
Model:                            OLS   Adj. R-squared (uncentered):             0.500
Method:                 Least Squares   F-statistic:                             76.05
Date:                Sat, 12 Oct 2024   Prob (F-statistic):                   2.78e-44
Time:                        20:05:34   Log-Likelihood:                        -3150.4
No. Observations:                 300   AIC:                                     6309.
Df Residuals:                     296   BIC:                                     6324.
Df Model:                           4
Covariance Type:            nonrobust
==============================================================================
                 coef    std err          t      P>|t|      [0.025      0.975]
------------------------------------------------------------------------------
SARSCoV2     2.68e-08   4.74e-09      5.657      0.000    1.75e-08    3.61e-08
RSV         2.759e-07   1.23e-07      2.250      0.025    3.46e-08    5.17e-07
IAV         2.948e-07   7.48e-08      3.939      0.000    1.48e-07    4.42e-07
IBV         5.629e-07   8.73e-08      6.451      0.000    3.91e-07    7.35e-07
==============================================================================
Omnibus:                       50.597   Durbin-Watson:                   0.881
Prob(Omnibus):                  0.000   Jarque-Bera (JB):               90.895
Skew:                           0.926   Prob(JB):                     1.83e-20
Kurtosis:                       4.960   Cond. No.                         35.9
==============================================================================

Notes:
[1] R² is computed without centering (uncentered) since the model does not contain a constant.
[2] Standard Errors assume that the covariance matrix of the errors is correctly specified.
```

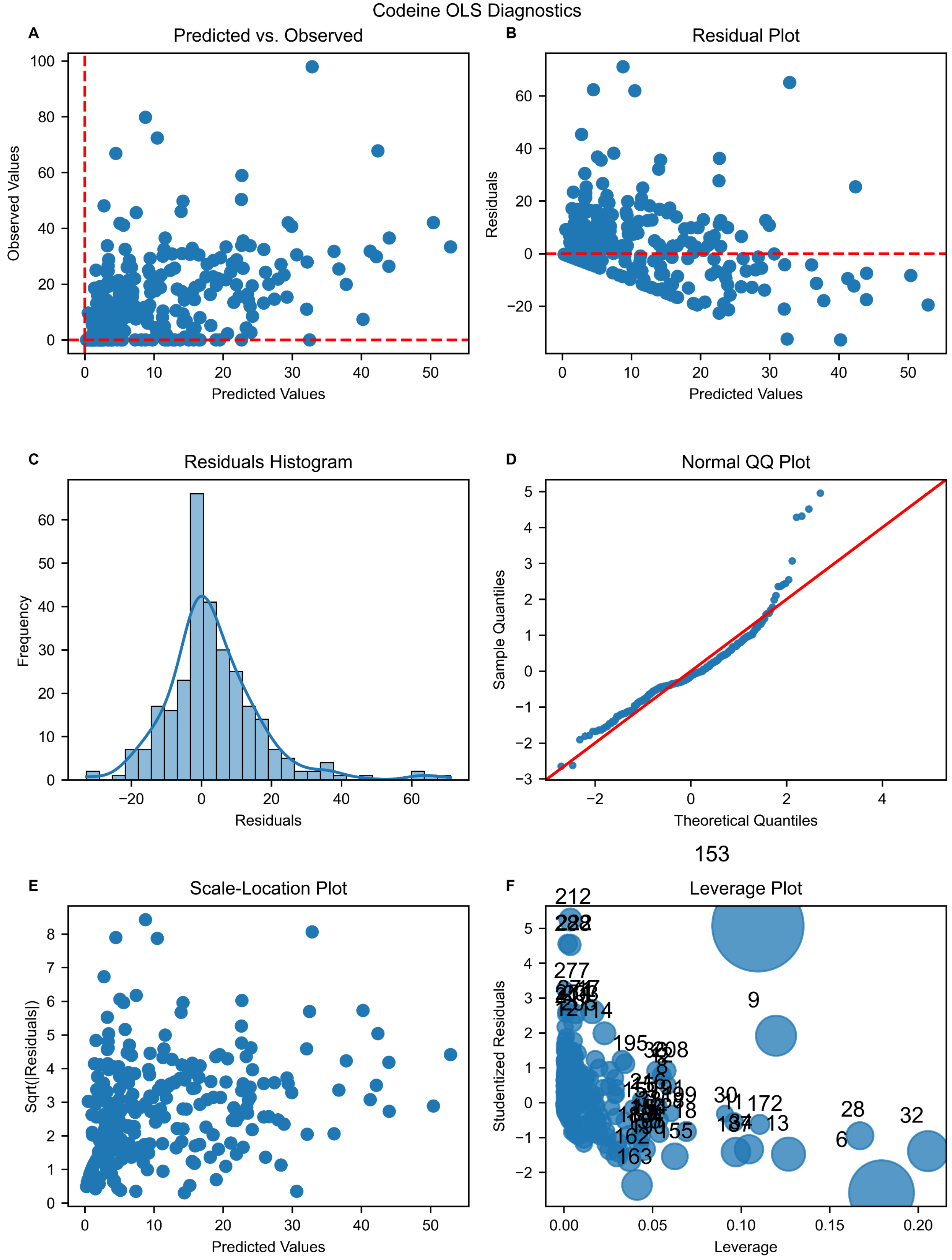


Figure SI 24: Diagnostic plots for the Ordinary Least Squares (OLS) regression analysis using 7-day centered median gene copy number loads of SARS-CoV-2, RSV, IAV, and IBV as predictors, with baseline-subtracted pharmaceutical loads as the response variable. The figure includes: (A) Predicted vs. Observed Values, depicting the correlation between predicted and observed pharmaceutical loads; (B) Residual Plot, illustrating residuals against predicted values to assess homoscedasticity; (C) Residuals Histogram, showing the residual distribution with an overlaid kernel density estimate; (D) Normal Q-Q Plot, evaluating the normality of residuals; (E) Scale-Location Plot, revealing the spread of residuals to detect non-constant variance; and (F) Leverage Plot, highlighting influential data points using Cook's distance.

Table SI 9: Ordinary Least Squares (OLS) regression results using gene copy loads of SARS-COV-2, RSV, IAV and IBV as input as predicting variable and baseline-subtracted acetaminophen loads as the response variable.

```
                                 OLS Regression Results                                
=======================================================================================
Dep. Variable:                Codeine   R-squared (uncentered):                   0.524
Model:                            OLS   Adj. R-squared (uncentered):              0.518
Method:                 Least Squares   F-statistic:                              79.35
Date:                Sat, 12 Oct 2024   Prob (F-statistic):                    2.64e-45
Time:                        20:05:21   Log-Likelihood:                         -1186.5
No. Observations:                 292   AIC:                                      2381.
Df Residuals:                     288   BIC:                                      2396.
Df Model:                           4                                                  
Covariance Type:            nonrobust                                                  
==============================================================================
                 coef    std err          t      P>|t|      [0.025      0.975]
------------------------------------------------------------------------------
SARSCoV2    4.012e-11   7.61e-12      5.273      0.000    2.51e-11    5.51e-11
RSV         8.911e-10   1.96e-10      4.536      0.000    5.04e-10    1.28e-09
IAV         2.939e-10    1.2e-10      2.454      0.015    5.81e-11     5.3e-10
IBV         7.986e-10    1.4e-10      5.715      0.000    5.24e-10    1.07e-09
==============================================================================
Omnibus:                      100.429   Durbin-Watson:                   1.583
Prob(Omnibus):                  0.000   Jarque-Bera (JB):              398.586
Skew:                           1.417   Prob(JB):                     2.81e-87
Kurtosis:                       7.973   Cond. No.                         35.9
==============================================================================

Notes:
[1] R² is computed without centering (uncentered) since the model does not contain a constant.
[2] Standard Errors assume that the covariance matrix of the errors is correctly specified.
```

#### Ordinary least squares analyses of additional pharmaceuticals

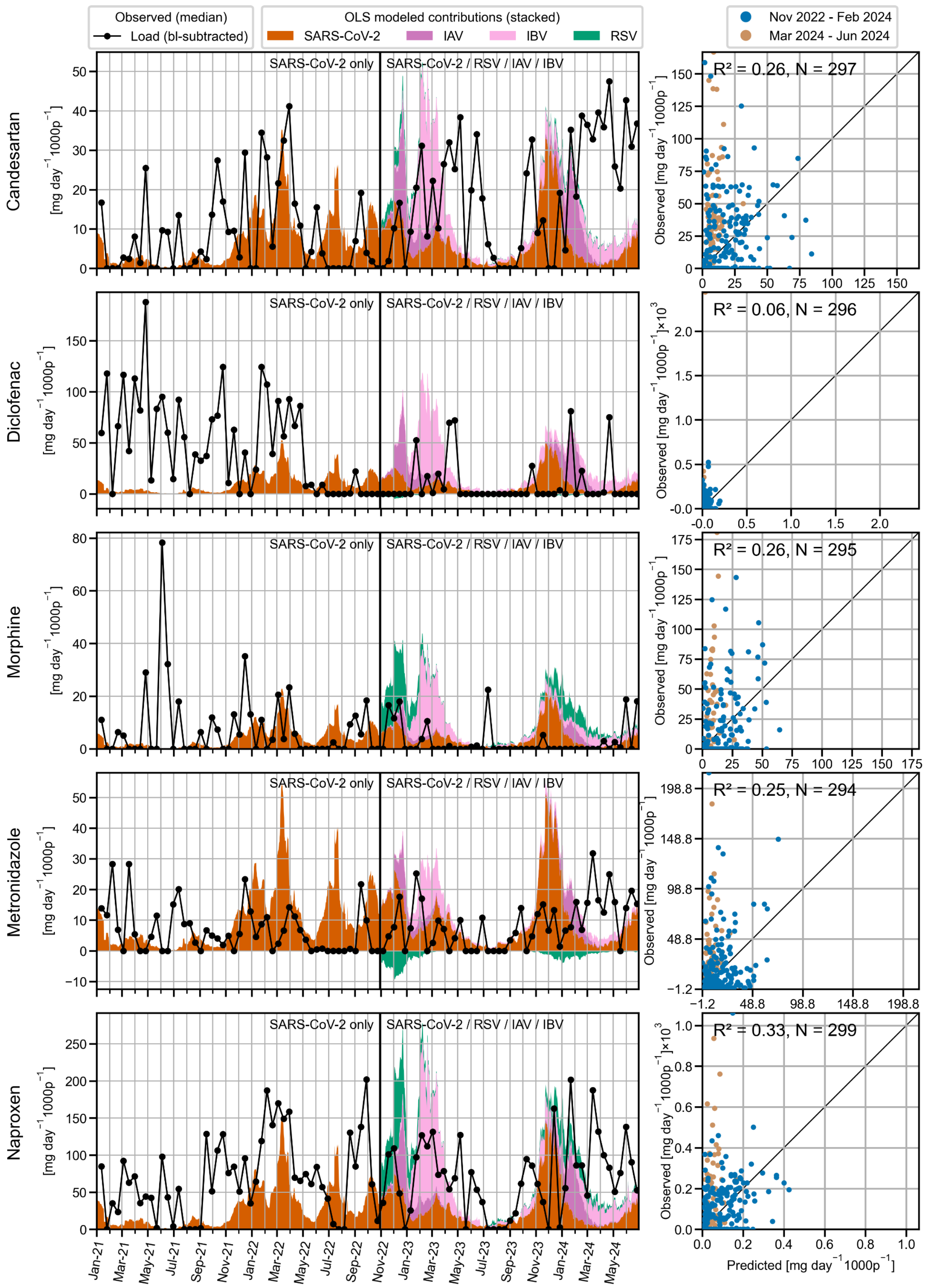


Figure SI 25: Modeled contributions of respiratory viruses to pharmaceutical loads. The left panel shows time series plots of observed baseline-subtracted pharmaceutical loads (black line) alongside the modeled contributions based on the median viral gene copy loads of SARS-CoV-2, RSV, IAV, and IBV, estimated using an ordinary least squares (OLS) model. The colored areas represent the stacked contributions of each virus to the overall baseline-subtracted pharmaceutical load. Baseline loads were determined from location-specific mean values during periods of low respiratory virus exposure (June 2021 and July to August 2023). For data before November 2022 (indicated by the vertical line), the model was retrospectively applied to SARS-CoV-2, as it was the only virus measured during those periods. The right panel presents scatter plots comparing observed versus predicted baseline-subtracted pharmaceutical loads for two distinct time periods: November 2022 to February 2024 (blue), March 2024 to June 2024 (orange). Each scatter plot includes the uncentered coefficient of determination ($R^2$) and the number of observations (N). The diagonal black line represents the 1:1 line, indicating perfect agreement between observed and predicted values.

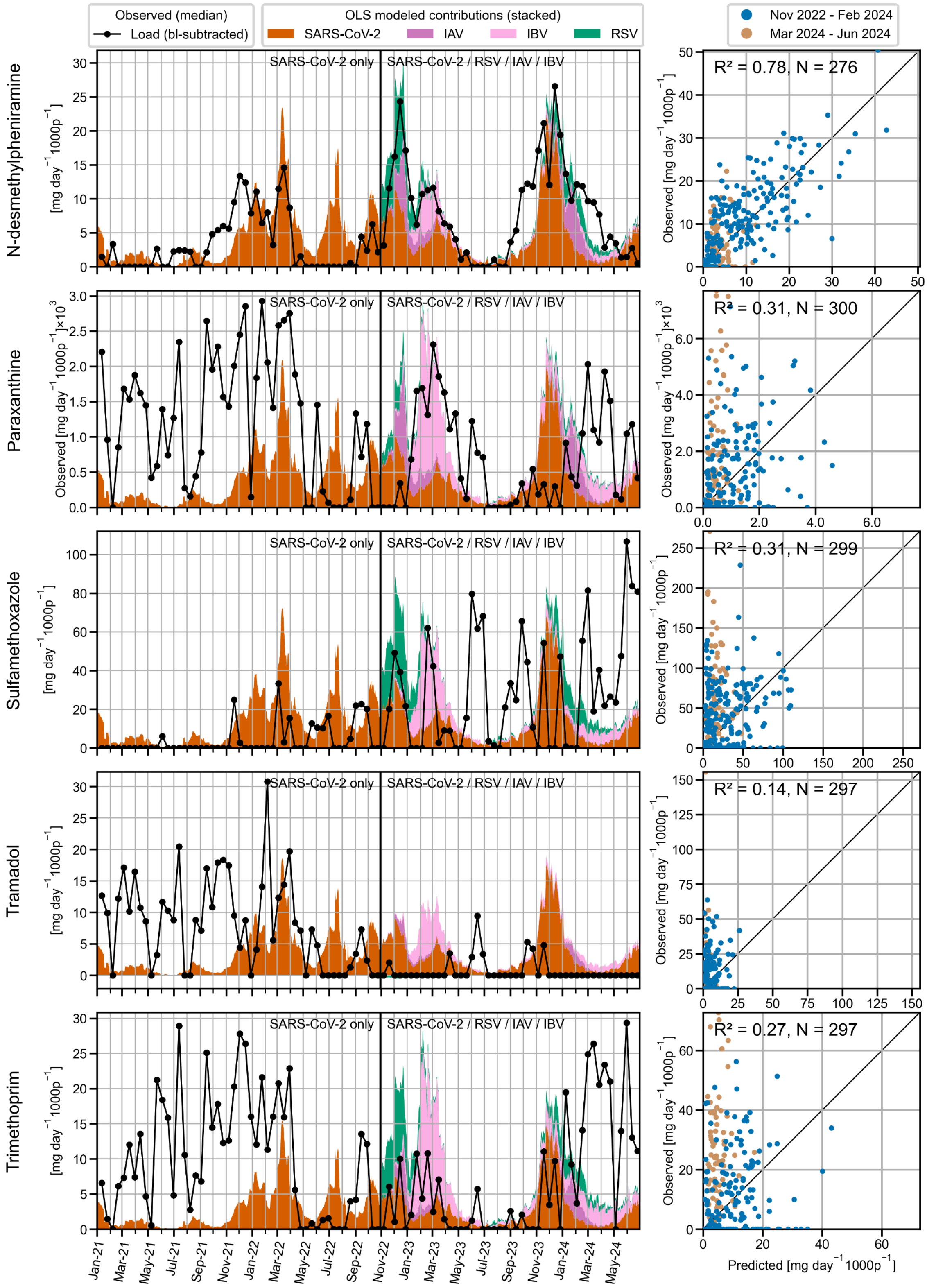


Figure SI 26: Modeled contributions of respiratory viruses to pharmaceutical loads. The left panel shows time series plots of observed baseline-subtracted pharmaceutical loads (black line) alongside the modeled contributions based on the median viral gene copy loads of SARS-CoV-2, RSV, IAV, and IBV, estimated using an ordinary least squares (OLS) model. The colored areas represent the stacked contributions of each virus to the overall baseline-subtracted pharmaceutical load. Baseline loads were determined from location-specific mean values during periods of low respiratory virus exposure (June 2021 and July to August 2023). For data before November 2022 (indicated by the vertical line), the model was retrospectively applied to SARS-CoV-2, as it was the only virus measured during those periods. The right panel presents scatter plots comparing observed versus predicted baseline-subtracted pharmaceutical loads for two distinct time periods: November 2022 to February 2024 (blue), March 2024 to June 2024 (orange). Each scatter plot includes the uncentered coefficient of determination ($R^2$) and the number of observations (N). The diagonal black line represents the 1:1 line, indicating perfect agreement between observed and predicted values.

**Ordinary least squares analyses with SARS-COV-2 wastewater loads as only predicting variable**

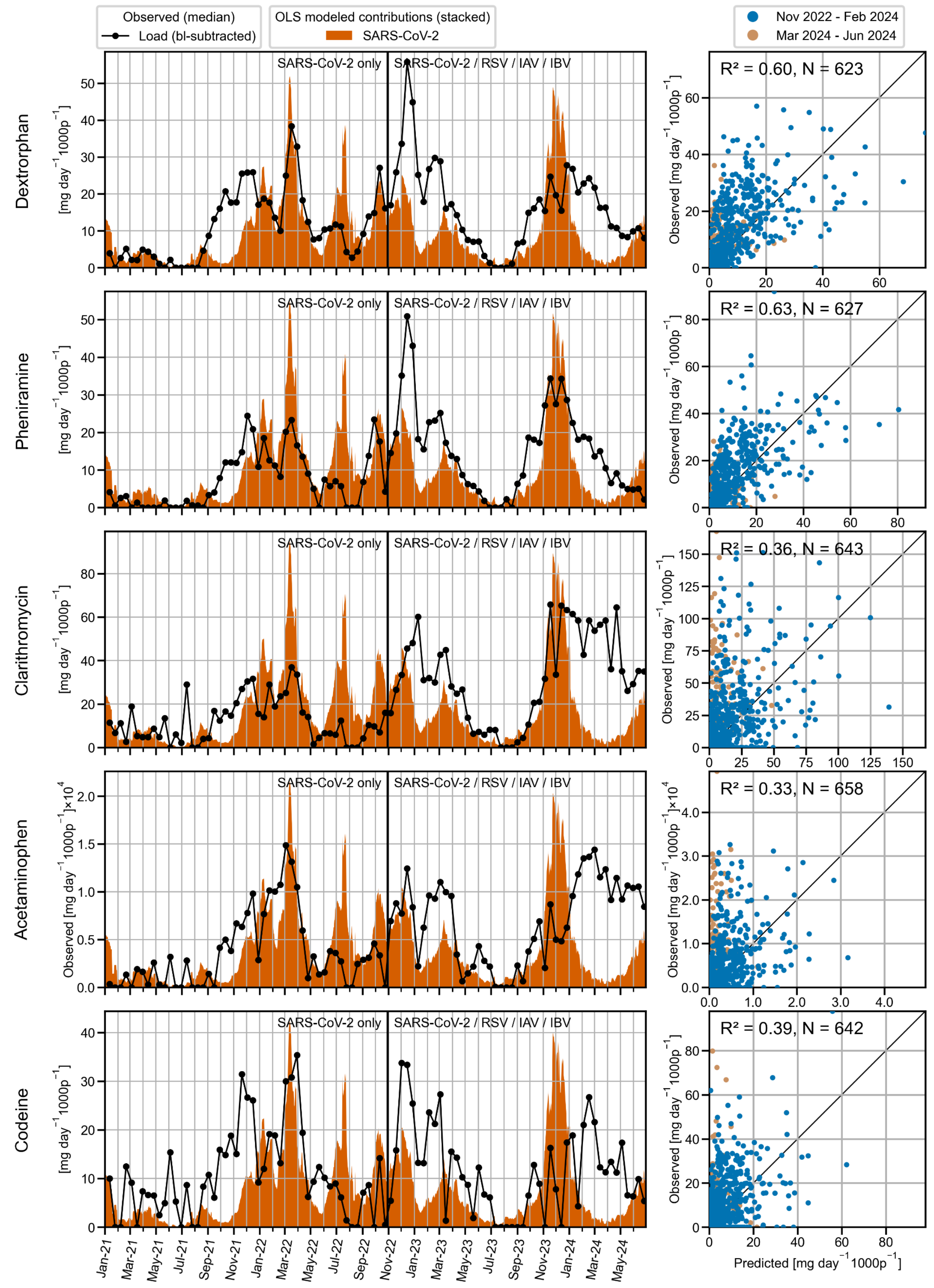


Figure SI 27: Modeled contributions of respiratory viruses to pharmaceutical loads. The left panel shows time series plots of observed baseline-subtracted pharmaceutical loads (black line) alongside the modeled contributions based on the median viral gene copy loads of SARS-CoV-2, estimated using an ordinary least squares (OLS) model. The colored areas represent the stacked contributions of each virus to the overall baseline-subtracted pharmaceutical load. Baseline loads were determined from location-specific mean values during periods of low respiratory virus exposure (June 2021 and July to August 2023). The right panel presents scatter plots comparing observed versus predicted baseline-subtracted pharmaceutical loads for two distinct time periods: November 2022 to February 2024 (blue), March 2024 to June 2024 (orange). Each scatter plot includes the uncentered coefficient of determination ($R^2$) and the number of observations (N). The diagonal black line represents the 1:1 line, indicating perfect agreement between observed and predicted values.

##### SI 2.4.2 **Ordinary least squares analyses with clinical cases as predicting variables**

To investigate whether the temporal occurrence of additional respiratory viruses recorded in the Swiss Sentinel Surveillance System (Sentinella) could explain unexplained gaps in pharmaceutical consumption, we conducted a series of separate OLS analyses. Initially, these analyses utilized Sentinel data for SARS-CoV-2, RSV, IAV, and IBV, which were also measured in wastewater. Subsequently, we expanded these analyses to include case data for either rhinovirus, an 'others' category (comprising seasonal human coronaviruses (HCoVs), bocavirus, human metapneumovirus (hMPV), and parainfluenza viruses (types 1-4)), or negative test results. The inclusion of negative test results aimed to identify potential triggers of acute respiratory infection (ARI) or influenza-like illness (ILI) symptoms in cases where no respiratory virus was detected. All analyses were based on median baseline-subtracted pharmaceutical loads, with Sentinella providing the national-level clinical case data.

#### OLS analysis with SARS-COV-2, RSV, IAV, IBV cases as predictors

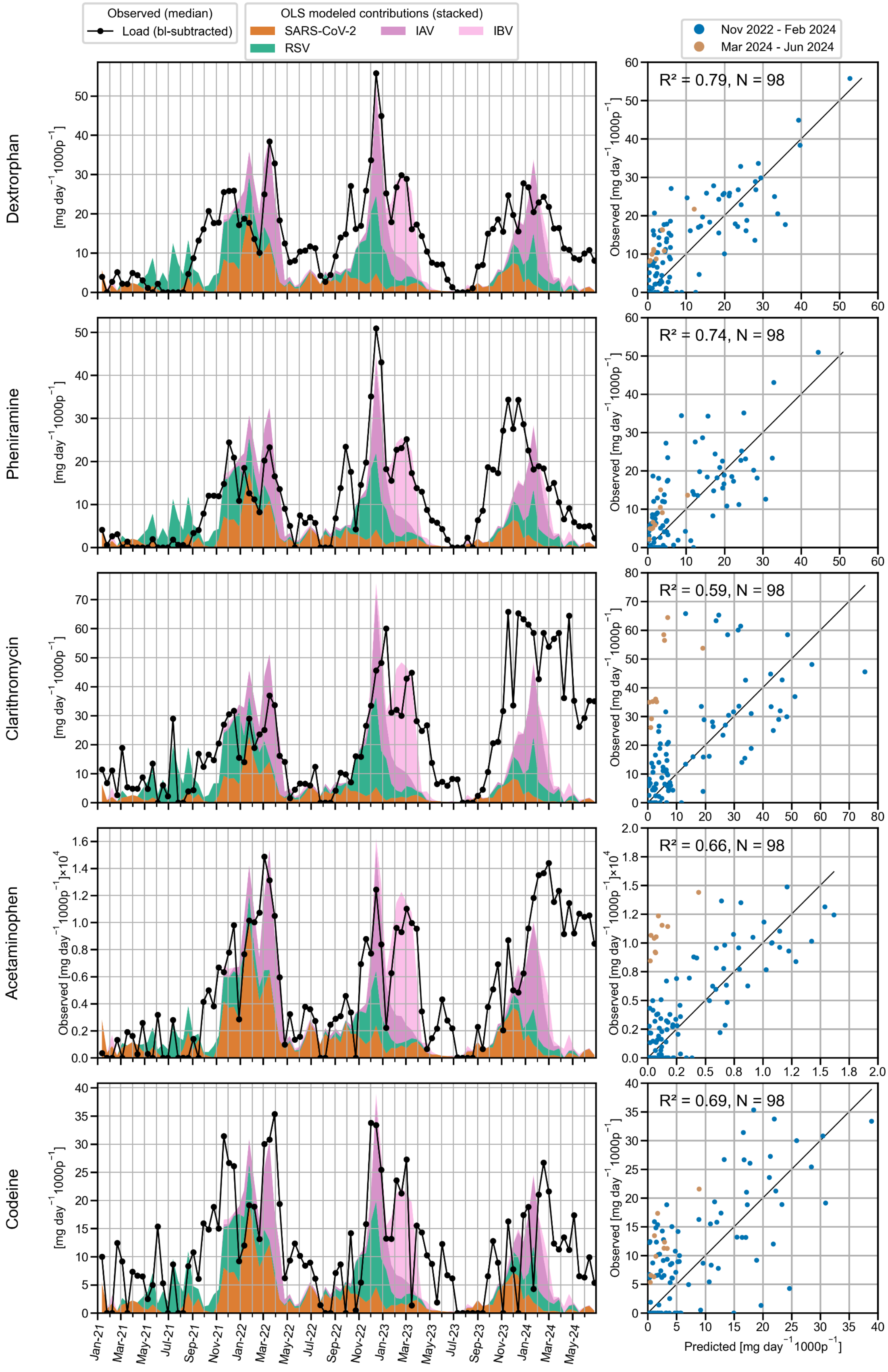


Figure SI 28: Modeled contributions of respiratory viruses to pharmaceutical loads. The left panel shows time series plots of observed median baseline-subtracted pharmaceutical loads (black line), alongside the modeled contributions of respiratory viruses, estimated using an ordinary least squares (OLS) model based on Sentinel system case data. The colored areas represent the stacked contributions of each virus to the overall baseline-subtracted pharmaceutical load. Baseline levels were determined using location-specific mean values during periods of minimal respiratory virus activity (June 2021 and July–August 2023). The right panel presents scatter plots comparing observed versus predicted baseline-subtracted pharmaceutical loads, with the uncentered coefficient of determination ($R^2$) and the number of observations (N). The diagonal black line represents the 1:1 line, denoting perfect agreement between observed and predicted values.

To investigate whether the temporal occurrence of additional respiratory viruses recorded in the Swiss Sentinel Surveillance System (Sentinella) could explain unexplained gaps in pharmaceutical consumption, we performed separate OLS analyses using case data for either rhinovirus, an "others" category (which included seasonal human coronaviruses (HCoVs), bocavirus, human metapneumovirus (hMPV), and parainfluenza viruses (types 1-4)), or negative test results. These were used in combination with case data for SARS-CoV-2, RSV, IAV, and IBV as predicting variables. Negative test results were included to capture potential triggers of acute respiratory infection or influenza-like illness symptoms where no respiratory virus was identified. Since Sentinella provides national-level clinical case data, the analyses were based on median baseline-subtracted pharmaceutical loads.

#### OLS Analysis with rhinovirus cases as an additional predictor

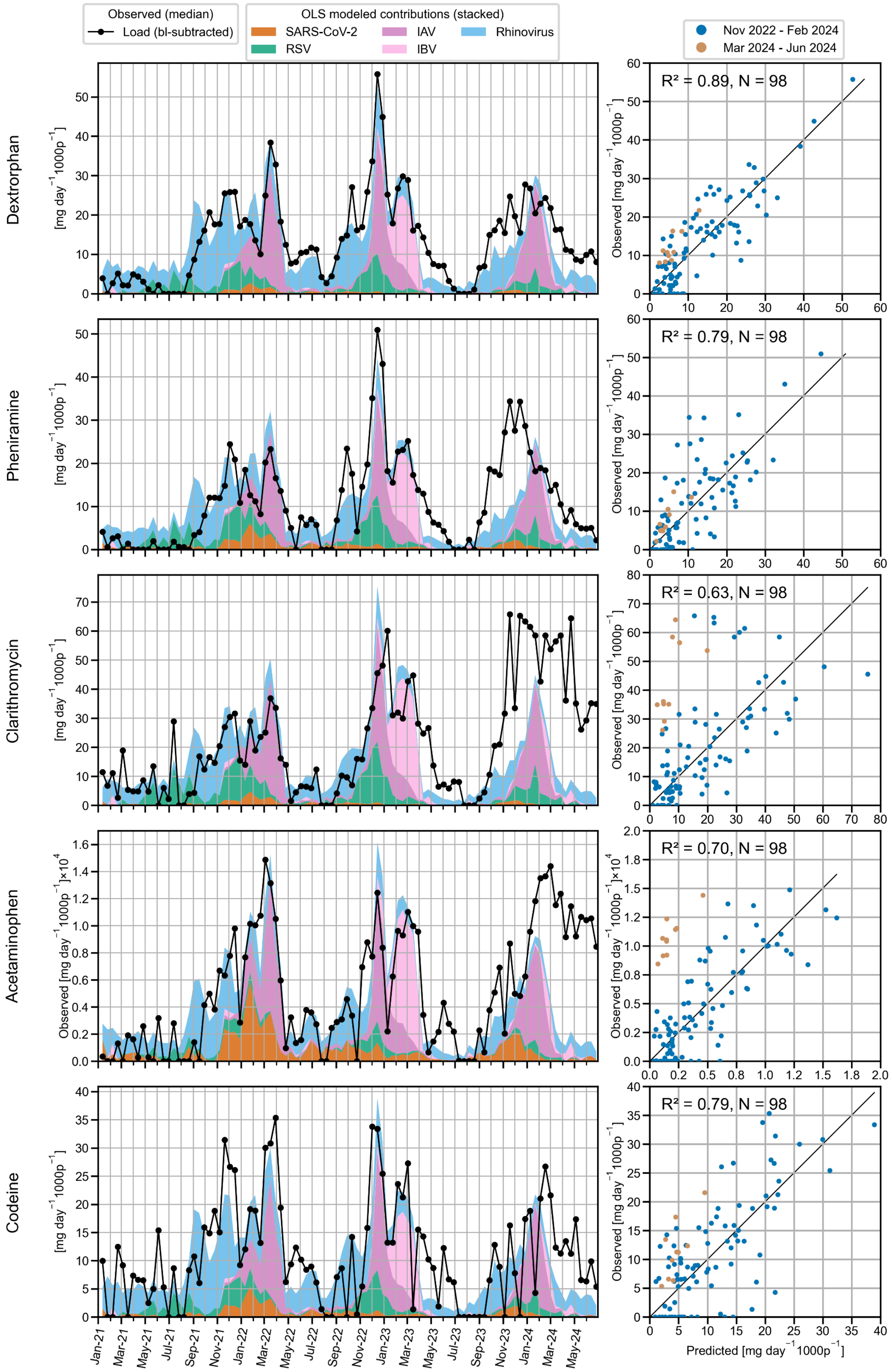


Figure SI 29: Modeled contributions of respiratory viruses to pharmaceutical loads. The left panel shows time series plots of observed median baseline-subtracted pharmaceutical loads (black line), alongside the modeled contributions of respiratory viruses, estimated using an ordinary least squares (OLS) model based on Sentinel system case data. The colored areas represent the stacked contributions of each virus to the overall baseline-subtracted pharmaceutical load. Baseline levels were determined using location-specific mean values during periods of minimal respiratory virus activity (June 2021 and July–August 2023). The right panel presents scatter plots comparing observed versus predicted baseline-subtracted pharmaceutical loads, with the uncentered coefficient of determination ($R^2$) and the number of observations (N). The diagonal black line represents the 1:1 line, denoting perfect agreement between observed and predicted values.

#### OLS Analysis with “Others” Category as an Additional Predictor

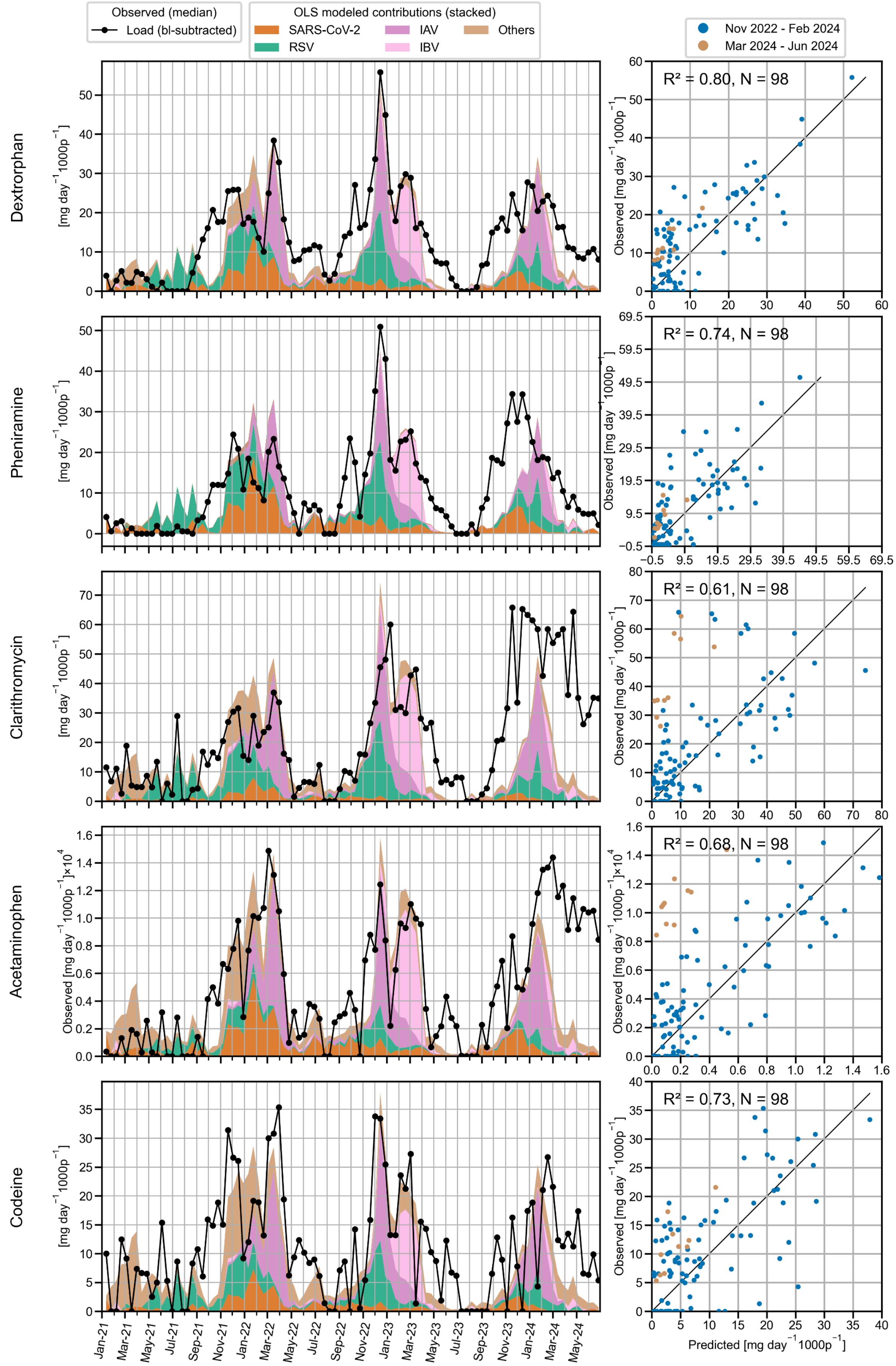


Figure SI 30: Modeled contributions of respiratory viruses to pharmaceutical loads. The left panel shows time series plots of observed median baseline-subtracted pharmaceutical loads (black line), alongside the modeled contributions of respiratory viruses, estimated using an ordinary least squares (OLS) model based on Sentinel system case data. The colored areas represent the stacked contributions of each virus to the overall baseline-subtracted pharmaceutical load. Baseline levels were determined using location-specific mean values during periods of minimal respiratory virus activity (June 2021 and July–August 2023). The right panel presents scatter plots comparing observed versus predicted baseline-subtracted pharmaceutical loads, with the uncentered coefficient of determination ($R^2$) and the number of observations (N). The diagonal black line represents the 1:1 line, denoting perfect agreement between observed and predicted values.

Including the "Others" category in the OLS modeling results in a negative coefficient for pheniramine, which is not physically meaningful and suggests a low explanatory power of this predictor.

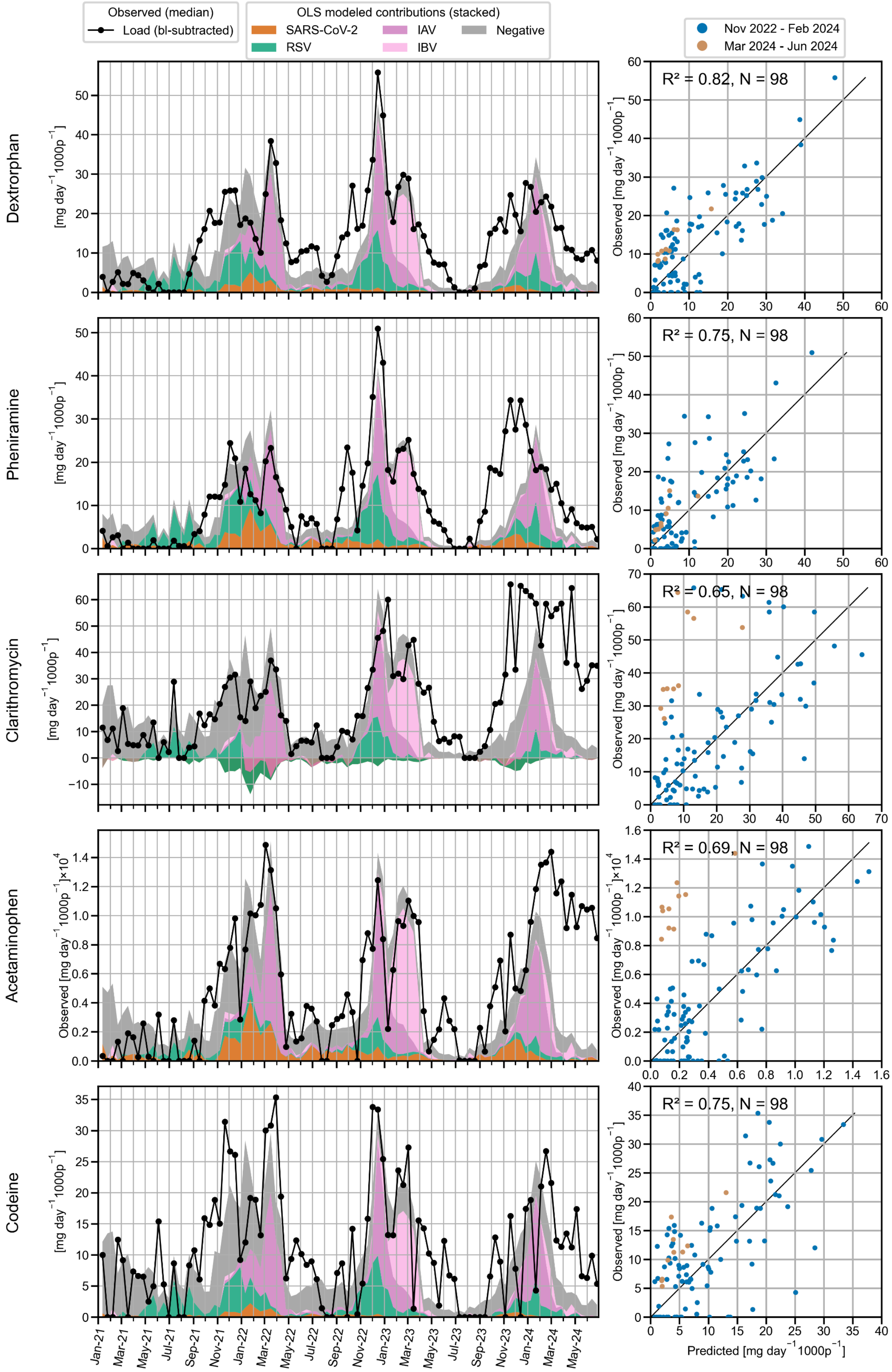


Figure SI 31: Modeled contributions of respiratory viruses to pharmaceutical loads. The left panel shows time series plots of observed median baseline-subtracted pharmaceutical loads (black line), alongside the modeled contributions of respiratory viruses, estimated using an ordinary least squares (OLS) model based on Sentinel system case data. The colored areas represent the stacked contributions of each virus to the overall baseline-subtracted pharmaceutical load. Baseline levels were determined using location-specific mean values during periods of minimal respiratory virus activity (June 2021 and July–August 2023). The right panel presents scatter plots comparing observed versus predicted baseline-subtracted pharmaceutical loads, with the uncentered coefficient of determination ($R^2$) and the number of observations (N). The diagonal black line represents the 1:1 line, denoting perfect agreement between observed and predicted values.

Including the "Negative" category in the OLS modeling results in a negative coefficient for clarithromycine, which is not physically meaningful and suggests a low explanatory power of this predictor.

### SI 3 In-sample stability of chemical markers

Stability experiments demonstrated that the degradation of chemical compounds within the samples under study conditions is negligible. The chemicals remain stable over a 24-hour sampling period at 4°C and maintain concentrations within 80% of their original levels even when stored for more than a year at -20°C (long-term storage).

#### SI 3.1 Sample preparation and analytical procedure

The stability of the target analytes was evaluated in both spiked wastewater and Evian mineral water across different storage temperatures (-20°C, 4°C, and 22°C).
A 24-hour composite influent sample from Zurich Werdhölzli WWTP, collected under dry weather conditions on 28.03.2022, was used to represent the wastewater matrix. This sample was stored in a muffled 1000 mL Schott DURAN borosilicate glass bottle (Kisker Biotech) at 4°C in the dark for four days before the experiment commenced. Prior to spiking, the bottle was gently shaken and allowed to sit for 1.5 hours to permit the larger particles to settle. 700 mL of the supernatant was then transferred to a muffled bulkhead bottle, where it was spiked with an ethanol-based mix (1.7 mL) to reach a final concentration of 1000 ng/L.
After shaking, the bottle was left to equilibrate for 2 hours. Following equilibration, 20 mL of the supernatant was withdrawn to measure the pH (SevenMulti, Mettler Toledo), and the remaining solution was aliquoted into muffled 1.5 mL MS glass vials (BGB Analytik). The same steps were followed to prepare the Evian mineral water samples.
At the start of the experiment, aliquots were incubated at their designated temperatures. Following incubation, samples were thawed for 1 minute under lukewarm water (for frozen samples), spiked with 20 µL of a 60 µg/L ethanol-based isotope-labeled internal standard solution, shaken and vortexed, centrifuged at room temperature for 10 minutes at 279.5 RCF (5427 R, Eppendorf centrifuge), and analyzed immediately. All samples were examined using large-volume direct injection reversed-phase LC-HRMS, as detailed in the methods section.
Replicates were analyzed at the initial measurement (t0) for samples from the same matrix. Although most time points involved only one measurement per sample condition, replicates were occasionally measured, as detailed in the accompanying table (Table SI 10).
The procedure closely follows the method used in [1] for determining in-sample stability.

Table SI 10: Experimental conditions of in-sample stability experiment and number of samples measured per condition and time point

| | Wastewater Influent | | | Evian mineral water | |
|---|---|---|---|---|---|
| Time point | 22°C | 4°C | -20°C | °4C | -20°C |
| 0h | 3 | 3 | 3 | 3 | 3 |
| 1d | 1 | 1 | 1 | 1 | 1 |
| 2d | 1 | 1 | 1 | 1 | 1 |
| 3d | 1 | 1 | 1 | 1 | 1 |
| 7d / 8d | 2 | 2 | 2 | 2 | 2 |
| 9d / 10d | 2 | 2 | 2 | 2 | 2 |
| 14d / 15d | 2 | 2 | 2 | 2 | 2 |
| 20d | 1 | 1 | 1 | 1 | 1 |
| up to 286d | - | - | 1 | - | 1 |

Table SI 11: Result of pH measurements of the stability experiment samples after spiking and at the end of experiment

| Matrix | Temperature [°C] | pH | |
|---|---|---|---|
| | | After spiking | End of experiment (t=20d) |
| Wastewater influent | 22 | 7.6 | 7.6 |
| | 4 | 7.6 | 5.9 |
| | -20 | 7.6 | 8.2 |
| Evian mineral water | 4 | 7.7 | 8.2 |
| | -20 | 7.7 | 9.5 |

#### SI 3.2 Data analysis

The change in relative concentration over time was assessed by comparing the peak area ratio of each analyte to its corresponding internal standard (ISTD) for short-term in-sample stability. For long-term stability assessments, relative concentrations were used to account for potential variations in ISTD concentrations between the different ISTD mixes prepared over the course of several years. For each substance and condition, the average response ratio or concentration of the triplicate at the initial time point (t0) was normalized to 100%. Changes in relative concentration were subsequently calculated as a percentage of this initial area ratio or concentration, enabling the evaluation of degradation or formation over time.
To determine the degradation rate constant for each analyte in the specific matrix and under particular conditions, a non-linear pseudo first-order kinetic model was fitted to the data. The degradation process is described by Equation SI 7, where $A_0$ represents the initial response ratio or concentration, $A$ is the response ratio or concentration at a given time point, $k$ denotes the degradation rate constant, and $t$ represents time (in days).

$$A = A_0 e^{-kt} \qquad (SI\ 7)$$

The parameters *A0* and *k* were optimized using Scipy in Python.

#### SI 3.3 Stability results

Key pharmaceutical markers dextrorphan, pheniramine, clarithromycin, acetaminophen, and codeine, exhibited consistent stability when stored at 22°C for 15 days and -20°C for 400 days in both Evian mineral water and wastewater (Figures SI32 - SI35). The relative concentrations of these markers remained within the 80% to 120% range, as described by pseudo-first-order kinetics models. This indicates that the conditions and durations used in the current study are unlikely to result in noticeable degradation, ensuring reliable data for these compounds. This observation applies to most other markers tested. In contrast, antibiotics metronidazole and sulfamethoxazole showed notable degradation, particularly at 22°C in wastewater matrix. Additionally, morphine concentrations increased significantly in spiked wastewater at both 22°C and 4°C, and in Evian water at 22°C (Figures SI 32), likely due to the hydrolysis of co-spiked heroin and 6-acetylmorphine [2]. However, given the generally low levels of heroin and 6-acetylmorphine in real wastewater, this effect has limited impact on real-world measurements.

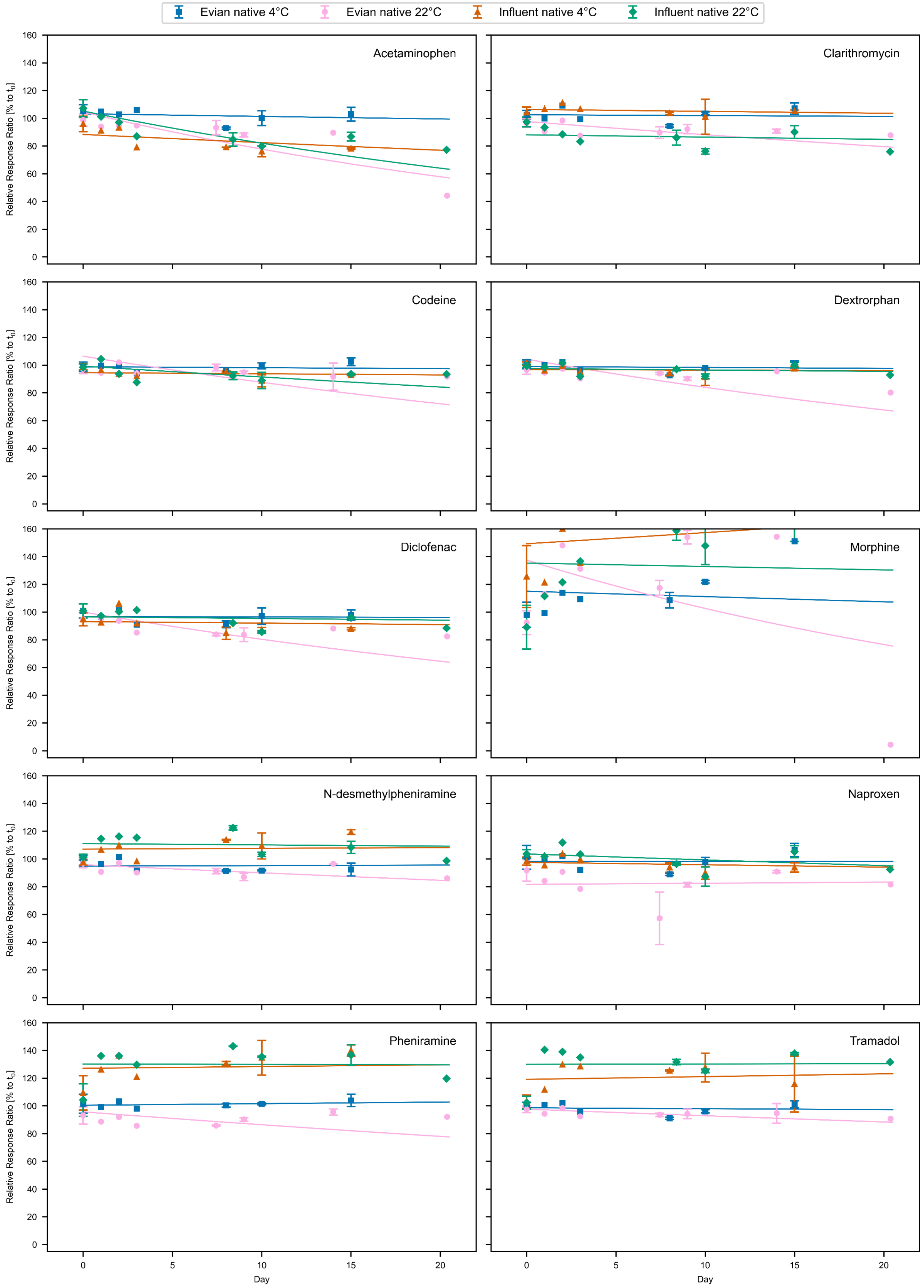


303

304 Figure SI 32: Stability of Chemicals in Wastewater Influent and Evian Mineral Water Incubated at 4°C and 22°C for 20 Days. The figure shows measured data points with
305 error bars representing standard deviations, alongside pseudo-first-order kinetic fits (lines).

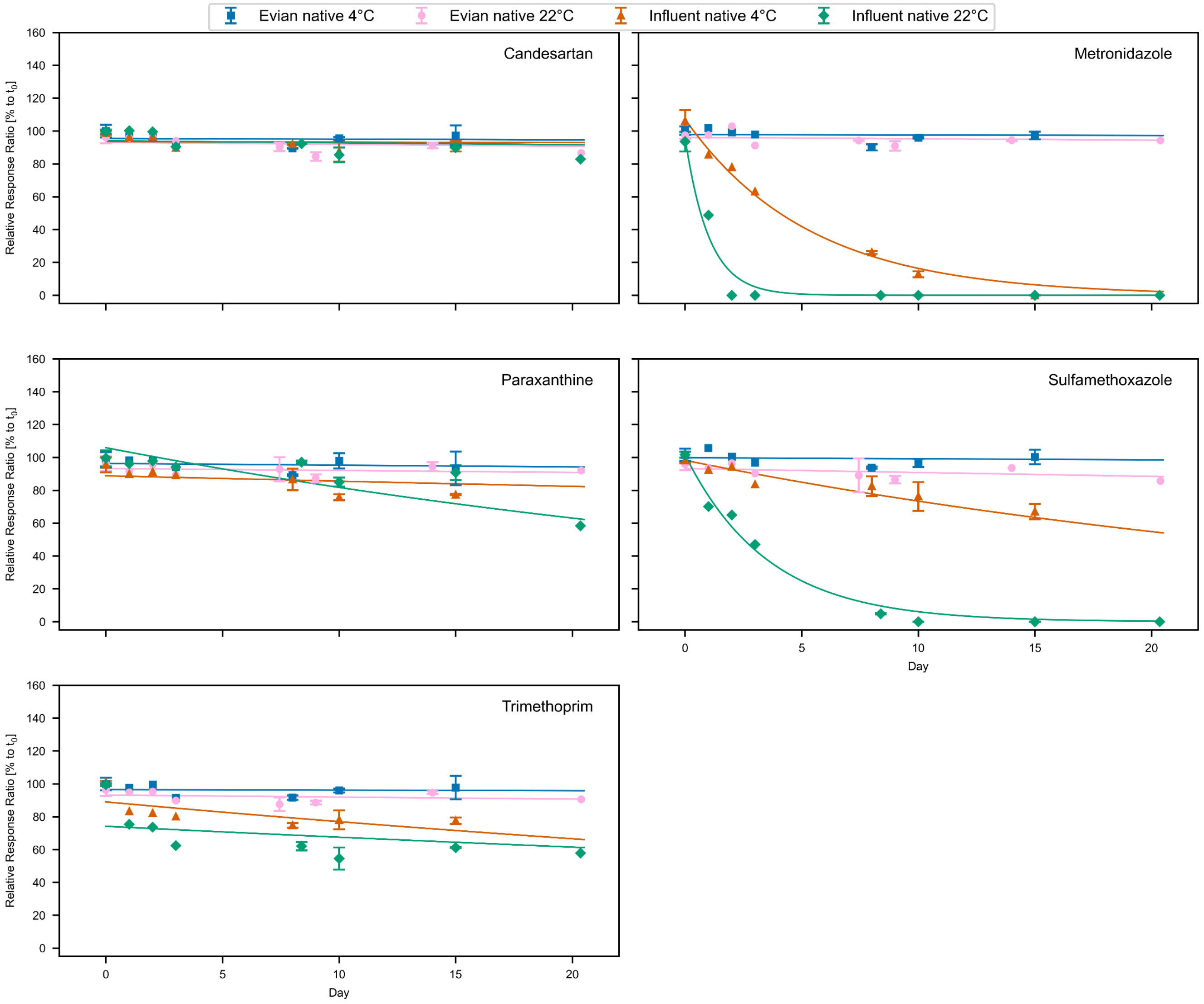


306

307 Figure SI 33: Stability of Chemicals in Wastewater Influent and Evian Mineral Water Incubated at 4°C and 22°C for 20 Days. The figure shows measured data points with
308 error bars representing standard deviations, alongside pseudo-first-order kinetic fits (lines).

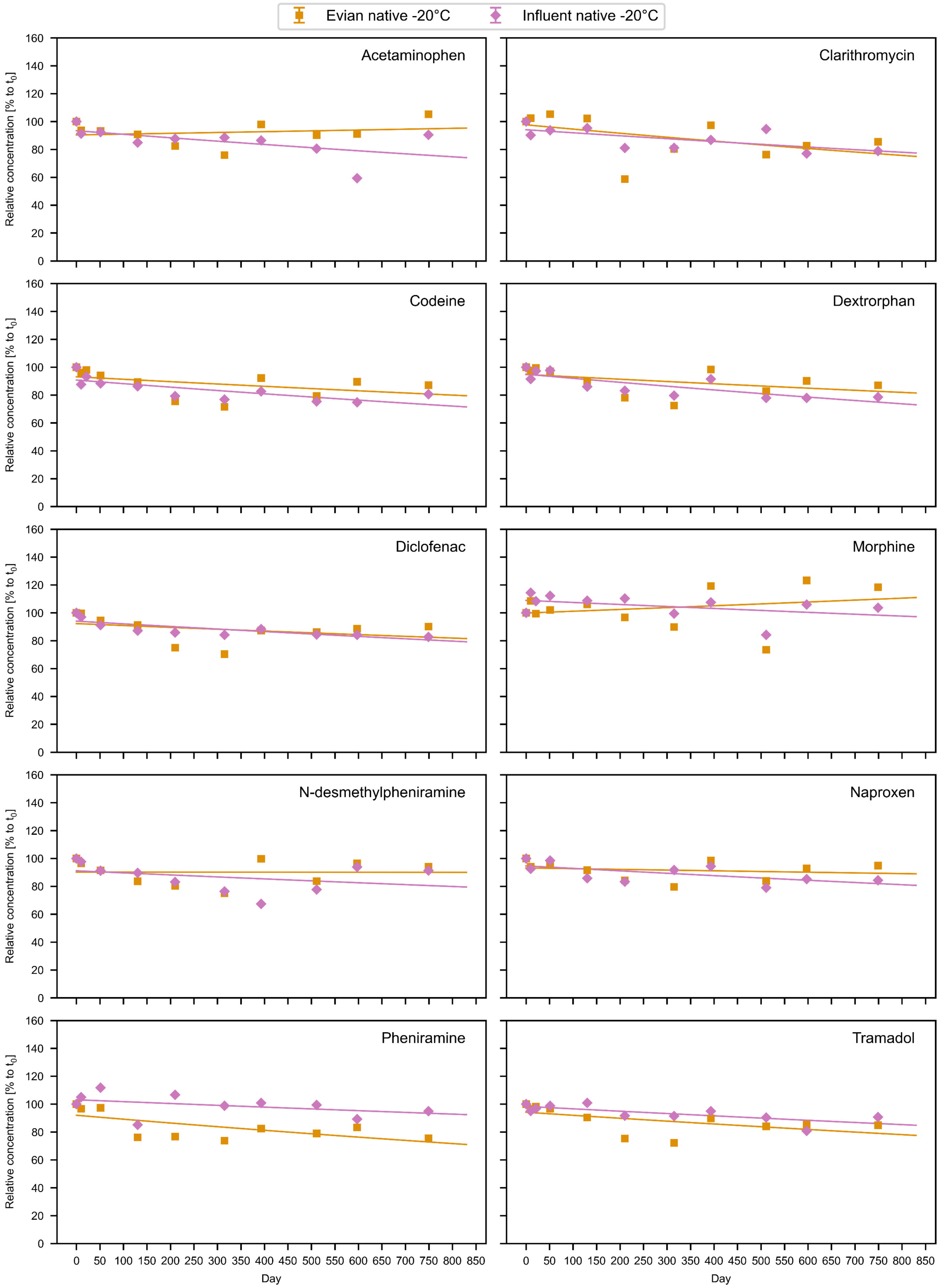


Figure SI 34: Long-term Stability of chemicals in Wastewater Influent and Evian Mineral Water Incubated at -20°C. The figure presents measured data points along with pseudo-first-order kinetic fits (lines).

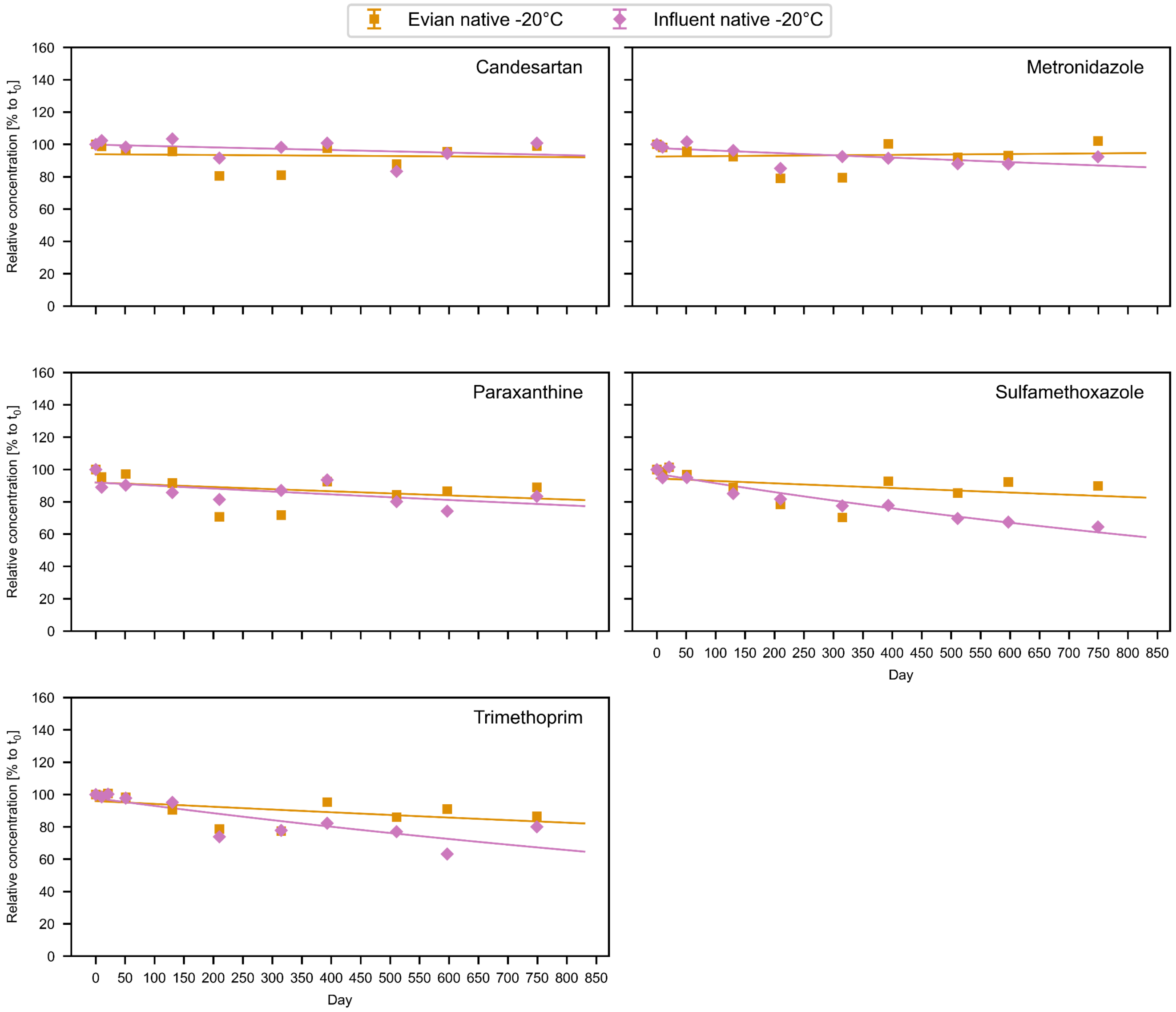


Figure SI 35: Long-term Stability of chemicals in Wastewater Influent and Evian Mineral Water Incubated at -20°C. The figure presents measured data points along with pseudo-first-order kinetic fits (lines).

#### SI 4 Licensed pharmaceutical products in Switzerland containing target chemicals

Table SI 12: Licensed Pharmaceutical Products in Switzerland Containing Dextromethorphan or Pheniramine, as Listed by the Swiss Agency for Therapeutic Products (Swissmedic)[3]. For products with multiple packaging sizes available, only a single option is listed.

| Compound | Product | Package Size | Unit | Composition | Dispensing Category Medicine | Application Area | License Holder |
|---|---|---|---|---|---|---|---|
| Dextromethorphan | GEM Antitussivum, Sirup | 200 | ml | dextromethorphanum 6.25 mg ut dextromethorphani hydrobromidum, …, excipiens ad solutionem pro 5 ml. | B | Husten | Tentan AG |
| Dextromethorphan | SUN STORE Dextromethorphan, Hustensirup | 200 | ml | dextromethorphanum 25 mg ut dextromethorphani hydrobromidum, … | B | Husten | Sun Store Health Care AG |
| Dextromethorphan | Amavita Dextromethorphan-N, Hustensirup | 200 | ml | dextromethorphanum 25 mg ut dextromethorphani hydrobromidum, … | B | Husten | Amavita Health Care AG |
| Dextromethorphan | Amavita Dextromethorphan, Hustentabletten | 16 | Tablette(n) | dextromethorphanum 25 mg ut dextromethorphani hydrobromidum,… | B | Husten | Amavita Health Care AG |
| Dextromethorphan | Coop Vitality Dextromethorphan-N, Hustensirup | 200 | ml | dextromethorphanum 25 mg ut dextromethorphani hydrobromidum, .. | B | Husten | Coop Vitality Health Care GmbH |
| Dextromethorphan | Coop Vitality Dextromethorphan, Hustentabletten | 16 | Tablette(n) | dextromethorphanum 25 mg ut dextromethorphani hydrobromidum, .. | B | Husten | Coop Vitality Health Care GmbH |
| Dextromethorphan | Bisolvon Dextromethorphan, Sirup | 200 | Flasche(n) | dextromethorphani hydrobromidum 10 mg corresp. dextromethorphanum 7.3 mg, … | B | trockener Reizhusten | Opella Healthcare Switzerland AG |
| Dextromethorphan | Irotussin Antitussivum, Sirup | 200 | ml | dextromethorphanum 6.25 mg ut dextromethorphani hydrobromidum, | B | Husten | Tentan AG |
| Dextromethorphan | Calmesin-Mepha, Sirup | 90 | ml | dextromethorphanum 11.5 mg ad resinam adsorbatum corresp., dextromethorphani hydrobromidum 15 mg, .. | B | Husten | Mepha Pharma AG |
| Dextromethorphan | Pulmofor, sirop | 200 | ml | dextromethorphani hydrobromidum 25 mg corresp. dextromethorphanum 18.3 mg, .. | B | Toux, particulièrement la toux sèche irritative | Verfora SA |
| Dextromethorphan | Pulmofor retard, capsules | 10 | Kapsel(n) | dextromethorphani hydrobromidum 50 mg corresp. dextromethorphanum 36.6 mg, .. | B | Toux, particulièrement la toux irritative | Verfora SA |
| Dextromethorphan | Pretuval Grippe & Erkältung, Filmtabletten | 20 | Tablette(n) | dextromethorphani hydrobromidum 20 mg corresp. dextromethorphanum 14.66 mg, .. | D | Erkältungskrankheiten mit Husten, Fieber und Schmerzen | Bayer (Schweiz) AG |
| Dextromethorphan | Bexin Hustentabletten, Filmtabletten | 24 | Tablette(n) | dextromethorphanum 25 mg ut dextromethorphani hydrobromidum, .. | B | Husten | Spirig HealthCare AG |
| Dextromethorphan | Pretuval Grippe & Erkältung C, Brausetabletten | 10 | Tablette(n) | dextromethorphani hydrobromidum 20 mg corresp. dextromethorphanum 14.66 mg, .. | D | Erkältungskrankheiten mit Husten, Fieber und Schmerzen | Bayer (Schweiz) AG |
| Dextromethorphan | Calmerphan-L, Sirup | 90 | ml | dextromethorphanum 11.5 mg ad resinam adsorbatum corresp., dextromethorphani hydrobromidum 15 mg, .. | B | Husten | Doetsch Grether AG |
| Dextromethorphan | Bexin, Hustentropfen | 20 | ml | dextromethorphanum 20.81 mg ut dextromethorphani hydrobromidum, .. | B | Husten | Spirig HealthCare AG |
| Dextromethorphan | Bexin, Hustensirup | 200 | ml | dextromethorphanum 25 mg ut dextromethorphani hydrobromidum, .. | B | Husten | Spirig HealthCare AG |
| Dextromethorphan | Emedrin N, Sirup | 150 | ml | dextromethorphanum 12.5 mg ut dextromethorphani hydrobromidum 17.075 mg, .. | B | Husten, insbesondere trockener Reizhusten | Streuli Pharma AG |
| Pheniramine | SUN STORE Flumol Grippe, Pulver zur Herstellung einer Lösung zum Einnehmen | 12 | Beutel | pheniramini maleas 20 mg, .. | D | Symptomatische Behandlung bei Erkältungskrankheiten | Sun Store Health Care AG |
| Pheniramine | NeoCitran Grippe / Erkältung für Erwachsene, Pulver zur Herstellung einer Lösung zum Einnehmen | 12 | Beutel | pheniramini maleas 20 mg.. | D | Symptomatische Behandlung bei Erkältungskrankheiten | Haleon Schweiz AG |
| Pheniramine | neotylol Grippe, Pulver zur Herstellung einer Lösung zum Einnehmen | 12 | Beutel | pheniramini maleas 20 mg, .. | D | Symptomatische Behandlung bei Erkältungskrankheiten | NOBEL Pharma Schweiz AG |
| Pheniramine | Dafalgan Grippal, Granulat zur Herstellung einer Lösung zum Einnehmen | 12 | Beutel | pheniramini maleas 25 mg, .. | D | Symptomatische Behandlung bei Erkältungskrankheiten | UPSA Switzerland AG |
| Pheniramine | Amavita Flumol Grippe, Pulver zur Herstellung einer | 12 | Beutel | pheniramini maleas 20 mg, .. | D | Symptomatische Behandlung bei | Amavita Health Care AG |

| Compound | Product | Package Size | Unit | Composition | Dispensing Category Medicine | Application Area | License Holder |
|---|---|---|---|---|---|---|---|
| | Lösung zum Einnehmen | | | | | Erkältungskrankheiten | |

Licensed products containing pheniramine are classified under dispensing category D, meaning they are available over-the-counter. Similarly, some dextromethorphan-containing products are also categorized as D, though the majority are classified under dispensing category B, which necessitates a medical prescription (Table SI 12).
Information on pharmaceutical products containing additional analyzed chemicals is included in the data package accompanying this publication.

### SI 5 References


1. Baumgartner S, Salvisberg M, Clot B, et al. Wastewater-Based Analysis of Antihistamines to Investigate Pollinosis Symptom Burden at Population-Scale. *medRxiv*. Published online June 21, 2024:2023.11.29.23299171. doi:10.1101/2023.11.29.23299171

2. Lin X, Choi PM, Thompson J, et al. Systematic Evaluation of the In-Sample Stability of Selected Pharmaceuticals, Illicit Drugs, and Their Metabolites in Wastewater. *Environ Sci Technol*. 2021;55(11):7418-7429. doi:10.1021/acs.est.1c00396

3. Swissmedic. Accessed August 20, 2024. https://www.swissmedic.ch/swissmedic/en/home/services/medicinal-product-information.html
